# A digital health approach for identifying polyendocrine metabolic ovarian syndrome using machine learning and body temperature

**DOI:** 10.64898/2026.07.23.26358666

**Authors:** Olalekan Awoniran, Deborah A Lawlor, Tom R Gaunt, Louise AC Millard

## Abstract

**Background:** Polyendocrine Metabolic Ovarian Syndrome (PMOS), formerly known as Polycystic Ovary Syndrome (PCOS), is a prevalent endocrine disorder with high rates of undiagnosed cases globally. Accessible screening tools are needed to facilitate appropriate management and earlier intervention. As PMOS is frequently characterised by oligo-anovulation, the absence of the characteristic rise in basal body temperature typically seen in ovulatory cycles may serve as a physiological marker for the condition.

**Objective:** This study aimed to assess the feasibility of using machine learning to identify individuals with PMOS from temperature data collected by a body-worn device.

**Methods:** We used data from 387 users of a vaginal temperature monitor (OvuSense^TM^) who responded to a questionnaire. The sample was restricted to individuals with at least three cycles with sufficient temperature data and whose PMOS case/control status could be determined from questions about prior clinical consultation for infertility and conditions for which they take medications. We randomly sampled three menstrual cycles for each participant and derived a set of cycle-level and user-level temperature features. Cycle-level features included cycle length and measures describing the temperature rise indicative of ovulation (e.g. temperature rise, cycle day of temperature rise start). We also constructed a reference cycle representing the typical bi-phasic cycle pattern (created using cycles from those without known fertility conditions) and used this to derive features describing how much a participant’s cycles differed from this reference. The cycle-level features were aggregated into user-level features by taking the minimum, maximum, median, and range of the cycle-level features across the three selected cycles for each participant. We used 5-fold nested cross-validation to evaluate the extent that PMOS could be predicted, at the cycle and user levels, using Logistic Regression (LR), Support Vector Machine (SVM), and Random Forest (RF).

**Results:** The average age of participants was 31.97 years (SD=4.58), with 49.6% having a self-reported PMOS diagnosis. The models demonstrated moderate discrimination, with cycle-level AUC-ROC scores ranging from 0.64 (SD=0.02) (LR) to 0.68 (SD=0.04) (RF), and user-level scores ranging from 0.65 (SD=0.07) (LR) to 0.70 (SD=0.04) (RF). All models were reasonably calibrated, though confidence intervals were wide (e.g. RF cycle-level: calibration slope = 0.83 (95% confidence interval [CI]: 0.68, 1.00), calibration intercept = 0.02 (95% CI: -0.11, 0.14); user-level: slope = 0.88 (95% CI: 0.69, 1.15), intercept = -0.01 (95% CI: -0.22, 0.16)).

**Conclusions:** This study demonstrates the potential of using body temperature from digital health devices to identify those with PMOS. Such a passive approach to identifying PMOS could help to identify undiagnosed PMOS in those who have not actively sought a diagnosis. Further research is needed to assess its predictive performance and acceptability in a general population using more widely used digital devices.

## Introduction

Polyendocrine metabolic ovarian syndrome (PMOS), formerly known as Polycystic Ovary Syndrome (PCOS)[1], is an endocrine disorder often characterised by features such as hyperandrogenism, hyperinsulinemia, ovulatory dysfunction, and infertility [2]. An estimated 8-13% of women of reproductive age are affected by PMOS, yet studies across various populations suggest that up to 70% of cases remain undiagnosed [3]. Evidence from large cohorts indicates that diagnosis is often significantly delayed, sometimes taking more than two years and visits to multiple health professionals before receiving a formal diagnosis [4,5]. This diagnostic gap is attributed largely to heterogeneity in symptoms, where individuals with PMOS may present with varying combinations and severities of reproductive, hyperandrogenic and metabolic symptoms [5]. Variations in diagnostic criteria across healthcare systems also contribute to this diagnostic gap [6,7]. The Rotterdam 2003 Consensus Workshop [8] provided the most widely used criteria for diagnosing the condition (including in Europe, UK and Australia) and requires the presence of at least two of the following three features: 1) hyperandrogenism, 2) oligo-anovulation, and 3) polycystic ovary morphology on ultrasound. Conversely, the AE-PCOS Society 2006 criteria are less widely used and require the presence of hyperandrogenism alongside ovarian dysfunction [6,9].

The high global rate of undiagnosed PMOS cases highlights the need for effective screening methods to identify those at high risk among those who are not currently seeking a diagnosis [10]. A key clinical feature of PCOS is ovulatory dysfunction, which is often manifested as oligo-anovulation. In a typical ovulatory cycle, progesterone’s thermal effect following ovulation raises basal body temperature (BBT), creating a characteristic biphasic pattern [11]. However, in anovulatory cycles, the absence of a luteal phase means this temperature rise does not occur [12]. The link between PMOS and oligo-anovulation suggests that longitudinal tracking of body temperature offers a plausible route to detecting PMOS in those who are using body-worn temperature-monitoring devices (e.g., smart watches or rings) but are not actively seeking a PMOS diagnosis.

BBT has been used for many years to identify ovulation and aid conception [13,14]. Traditionally, individuals would measure their temperature once daily after waking and record the data on a daily chart to track thermal shifts and determine when ovulation is likely to have occurred [15]. However, these point-in-time measurements are highly sensitive to external factors such as sleep disruptions, stress, or irregular measurement timing. More modern approaches now use wearable technology for automated prediction of ovulation [16,17] and fertile windows [18], as well as to forecast menstruation [19]. Devices available include those targeted at ovulation prediction [17], including skin-based [20,21] and vaginal sensors [22,23], and more general-purpose devices that include temperature tracking functionality [19]. These devices track temperature continuously during the night, in order to capture a better estimate of BBT from multiple readings ([24–27].

A number of previous studies have sought to develop predictive models for PMOS [28–31]. Several of these applied machine learning algorithms to a publicly available Kaggle dataset [32] containing hospital records of 541 individuals from Kerala, India [29–31]. Chauhan et al. (2021) collected data via questionnaire in 267 women in India, including general items such as age and lifestyle as well as questions specifically about PMOS features such as hair growth. They developed a phone app intended for initial screening among the general population to help individuals determine if they should seek further medical evaluation. We found only one study that has assessed the potential for body temperature to be used to predict PMOS [33]. They assessed a small number of pre-defined characteristics – the first day of temperature elevation, duration of the lower-temperature phase, and total daily amplitude – all of which were found to be predictive of PMOS. To our knowledge, no study has assessed the use of body temperature (or other data from digital devices) to develop and evaluate a predictive model for PMOS that integrates multiple features from body temperature data into a single model.

The global estimates of undiagnosed PMOS cases and the increasing prevalence of digital devices capable of capturing body temperature data present a possible opportunity for earlier detection of those with PMOS. The aim of this study was to evaluate whether PMOS may be identified using body temperature data collected from a device worn vaginally overnight. During the course of writing this manuscript, PCOS was renamed PMOS [1]. We use the term PMOS when referring to the condition but use PCOS for variable names and questionnaire wording to reflect their usage at the time of the research.

## Methodology

### Ethical approval

Ethical approval for this study was granted by the University of Bristol Faculty of Health Sciences Research Ethics Committee (Reference: 4527).

### Participant recruitment and informed consent

New, current, and previous OvuSense users were invited to participate in the study. Recruitment was conducted by posting invitations in the OvuSense Facebook group for current and previous users, inviting new users after device purchase, and inviting active users through occasional email invitations. Those invited were directed to a study website hosting a participant information sheet (provided in Multimedia appendix 5). The participant information sheet detailed the research objectives, the nature of data use, and protocols for data access. Participants were informed that they could withdraw their data at any point during the study by contacting viO HealthTech (who make OvuSense). Those wishing to participate were then directed to an online questionnaire (see below), and their informed consent was obtained as a mandatory step at the start of this questionnaire.

### Study data

We use data from (a) the OvuSense device, (b) the OvuSense smartphone app, and (c) a questionnaire deployed to OvuSense customers.

#### The OvuSense device and generated temperature data

The OvuSense device is worn overnight in the vagina and records temperature every five minutes [22]. Each morning, users synchronise the device with the OvuSense app on their phone, connecting it wirelessly via near-field communication. Users view their data on the associated smartphone app, including temperature charts and ovulation prediction. OvuSense users are advised not to wear the device during menstruation and are also informed that they do not need to wear it for the rest of the menstrual cycle once ovulation has been predicted. As a result, the data contain significant amounts of non-random missingness. The devices record temperature data with a precision of one thousandth of a degree Celsius.

#### Menstrual cycle start dates from the OvuSense app

As part of the standard OvuSense user process, users enter the start dates of each menstrual period into the OvuSense app, which marks the beginning of the cycle. This is so that the system can then identify the days pertaining to each cycle and predict ovulation within each of these cycles. As viO HealthTech Ltd store these derived cycles (i.e. the set of temperature data associated with a cycle ID, for each user), we use these directly in our prediction approach (see Cycle and validation section below).

#### Study questionnaire

Participants were asked for demographic details (e.g., education level and employment status), lifestyle factors (e.g., smoking status and alcohol intake), general health (e.g., existing health conditions and regular medications), and reproductive history (e.g., pregnancy history and fertility conditions). The full questionnaire is provided at https://github.com/MRCIEU/fertility-study (and Multimedia appendix 4).

Questionnaire deployment was managed by viO HealthTech Ltd, which linked the responses to OvuSense temperature data using the customer’s email address. To ensure participant privacy, all datasets were pseudonymised by viO HealthTech Ltd prior to transfer to the University of Bristol study team. The data received by the study team contained no personal identification fields, and all records were linked only through the pseudonymised, unique participant IDs. All data were transferred to the research team via a secure protocol.

### Overview of analytical pipeline

Our analytical pipeline is illustrated in Figure 1, and includes deriving a PCOS class variable, deriving predictors from the temperature data for each cycle, and then using these cycle-level features to derive predictors for each user. The specific steps are explained in the subsequent subsections.

**Figure 1:**
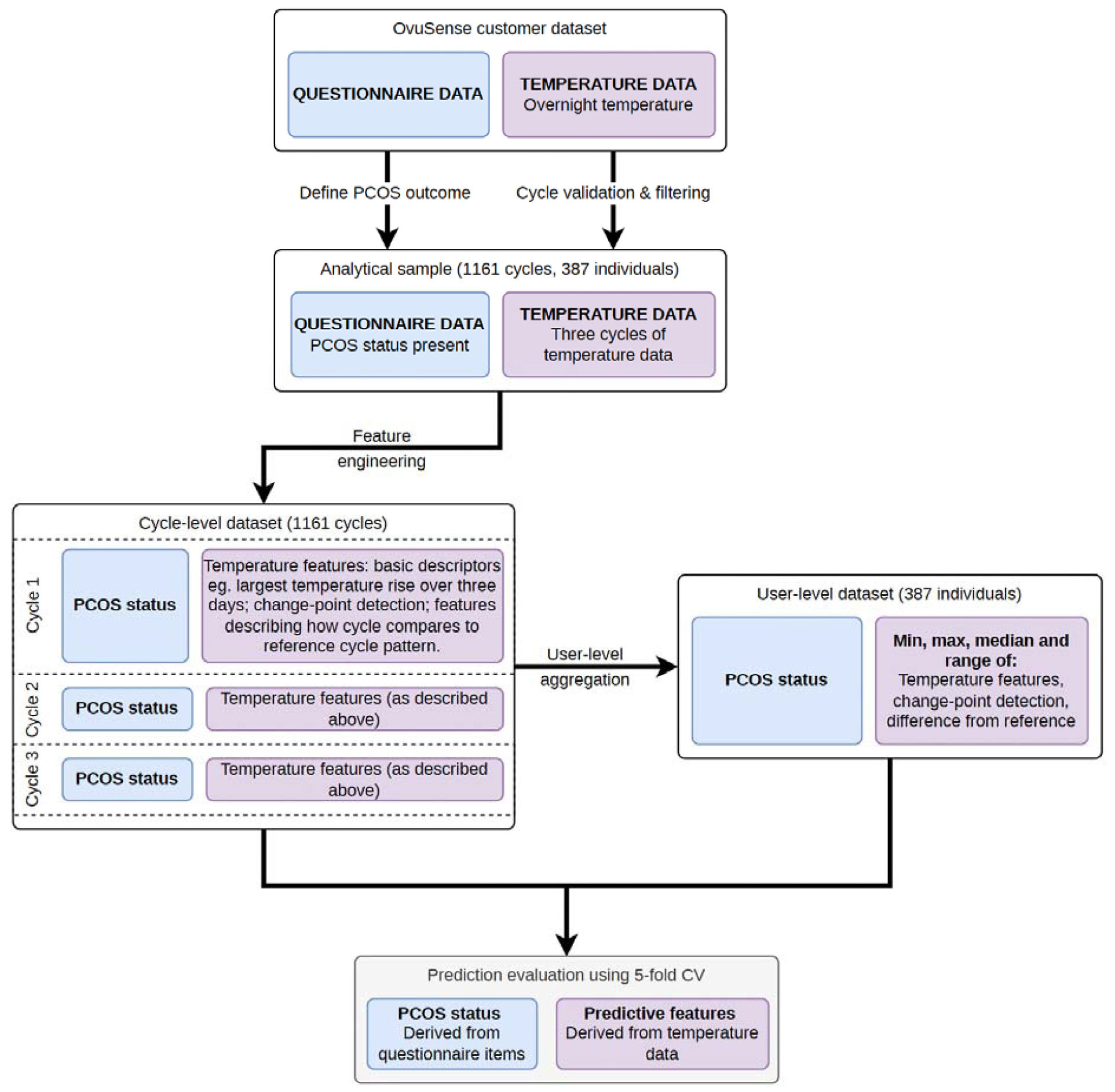
Data processing pipeline. The pipeline results in two feature sets, cycle-level and user-level, that are both evaluated for PCOS prediction. PCOS: Polycystic ovary syndrome; CV: cross-validation.

### PCOS class variable

We examined responses to a question about whether the women had ever visited a doctor due to infertility. Women who answered “Yes” were asked for the diagnosis of infertility provided by the doctor, with predefined categories of infertility (PCOS, other ovarian problem, endometriosis, problem with your cervix, partner low sperm count, overactive thyroid, underactive thyroid, pelvic inflammatory disease, diagnosis not made, other). Furthermore, women were asked, “Are there any problems for which you have regular treatment or medicine?” and asked to describe the problem and treatment or medication for each. Those who reported being diagnosed with PCOS by a doctor or stated that PCOS was a condition for which they took regular medication (or both) were classified as cases. Controls were defined as those who said they (a) had not visited a doctor for infertility and did not state PCOS as a condition they took medication for, or (b) had visited a doctor for infertility but did not receive a PCOS diagnosis and did not state PCOS as a condition they took medication for.

### Study participants

Between 21st August 2019 and 24th July 2024, OvuSense customers were invited to participate in this study through viO HealthTech customer emails. Previous or current customers were invited through being a member of the OvuSense Facebook group (approximately 5000 Facebook group members at the time of the recruitment post). The invitation included a link to the study participant information sheet (Multimedia Appendix 5). Of these, 683 (approximately 1.27%) provided informed consent and completed the questionnaire. From this group, 589 participants met our criteria for PCOS classification, and 575 of these individuals had temperature dataθ. In order to assess representativeness of our sample with the whole OvuSense customerbase we also used data from those who did not complete the questionnaire (and hence did not consent as part of this). Consent for these users was gained through OvuSense terms and conditions and ethics approval (described above) covers use of these wider OvuSense customer data.

#### Cycle validation and filtering

The 575 users with questionnaire and temperature data had 6,023 cycles recorded in the OvuSense data. We processed these cycles to remove those with insufficient temperature data. To do this, we first excluded the first five readings (25 minutes) of each night, as temperature measurements adjusted from ambient to intra-vaginal temperature during this period. Nightly records were considered valid if they had at least ten readings after excluding implausible BBT values less than 35.0°C or greater than 40.0°C). We removed 791 cycles with no valid nightly records, leaving 5,232 cycles from the 564 participants.eθ then excluded 68 cycles with incorrect start dates (null values or recorded after the first temperature reading) because they resulted in inaccurate cycle durations. Furthermore, we removed 558 cycles that lack a subsequent cycle start date, as we could not determine their end dates and cycle lengths from the provided cycles. From these, we removed θθcycles that were either very short (fewer than 10 days) or very long (more than 366 days) to increase the likelihood that the defined cycle periods menstrual cycles. θWe also dropped 35 cycles that had fewer than 10 valid nightly before 10 consecutive missing readings. We then θremoved 577 cycles that had less than 40% validnightly recordsθ. Following this validation process, 3906 cycles from 499 individuals, and these are referred to as valid cycles θThe cycle validation phases θare described further in Section A of the Multimedia Appendix 1.

#### Analytical sample

Out of the 499 participants who had at least one valid cycle, three were excluded because their data were used to establish a reference cycle (see ‘Feature construction from temperature data’ section below). Additionally, we excluded 108 individuals who had fewer than three valid cycles. One more participant was removed because they reported never having experienced menstrual bleeding, resulting in a final analytical sample of 387 women (including 192 participants assigned as PCOS cases and 195 assigned as PCOS controls). From these 387 participants, a total of 3,719 valid cycles were available (1,962 from those assigned as a PCOS case and 1,757 from those assigned as a PCOS control). To ensure a balanced contribution and equal weighting across all participants in our PCOS prediction evaluation, three cycles were randomly selected per participant, yielding a final set of 1,161 cycles. The participant flow diagram is illustrated in Figure 2, and Section B of Multimedia Appendix 1 details the PCOS outcome grouping of the analytical sample.

**Figure 2:**
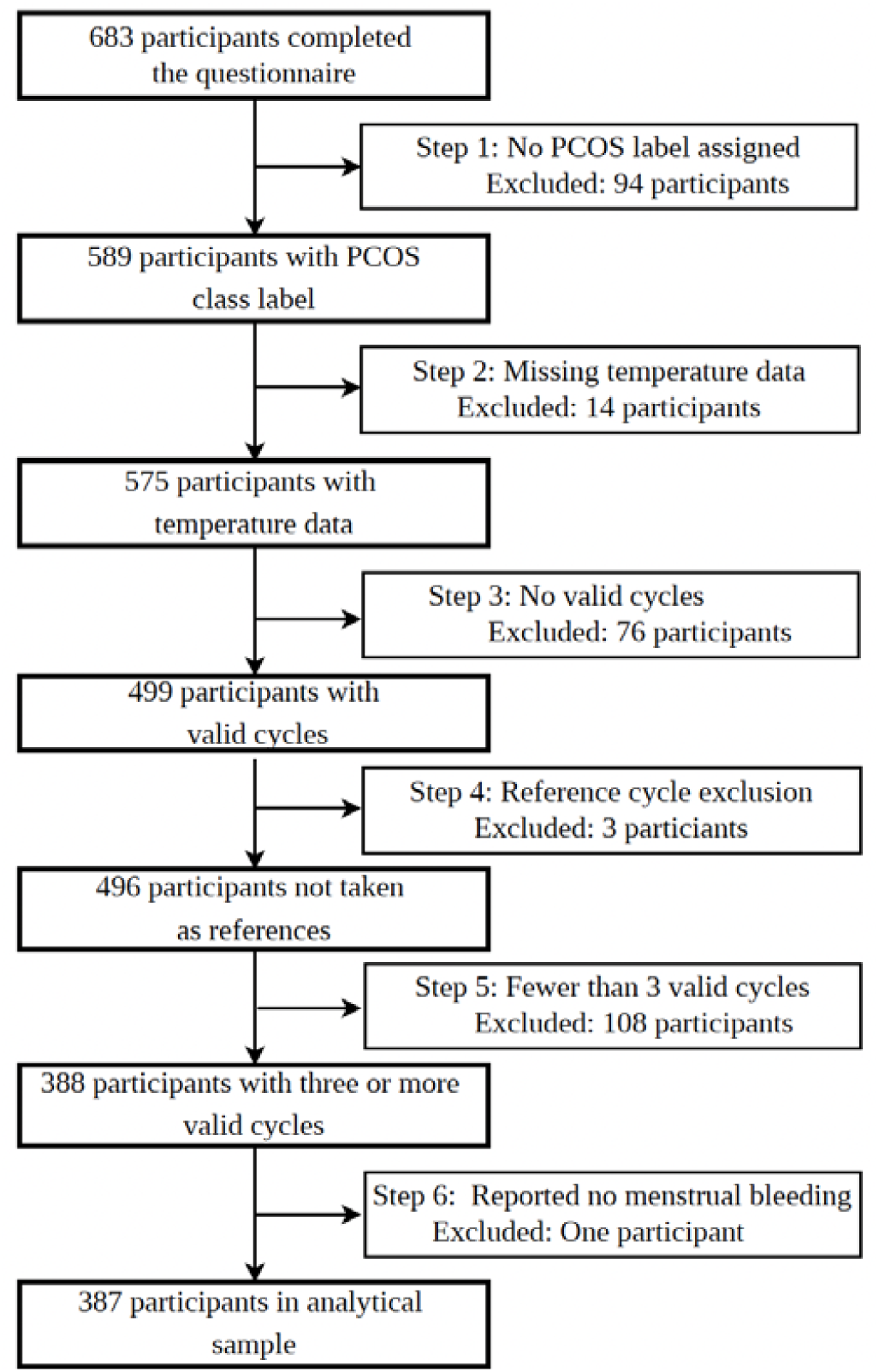
Participant flow and analytical sample selection. The diagram illustrates the stepwise exclusion criteria applied to the initial set of participants to reach the final analytical sample (n=387). Steps 1–3 focus on labelling and data availability, while Steps 4–6 ensure robustness of our approach evaluation.

### Temperature data pre-processing

The mean of each nightly reading was calculated to establish a single daily temperature value for each calendar day. To address data gaps, we imputed missing daily records within each valid cycle using linear interpolation [34]. Subsequently, the cycle temperature curves were smoothed using Savitzky-Golay filtering with a window length of 11 and a polynomial order of 2 [35,36]. This smoothing process reduces noise in the temperature data and highlights the underlying physiological patterns across the cycle [37]. These initial steps are shown in Figure 3 (Steps 1 and 2), with additional technical details provided in Section C of Multimedia Appendix 1. This intermediate output is hereafter referred to as “processed-unstandardised” temperature data.

**Figure 3:**
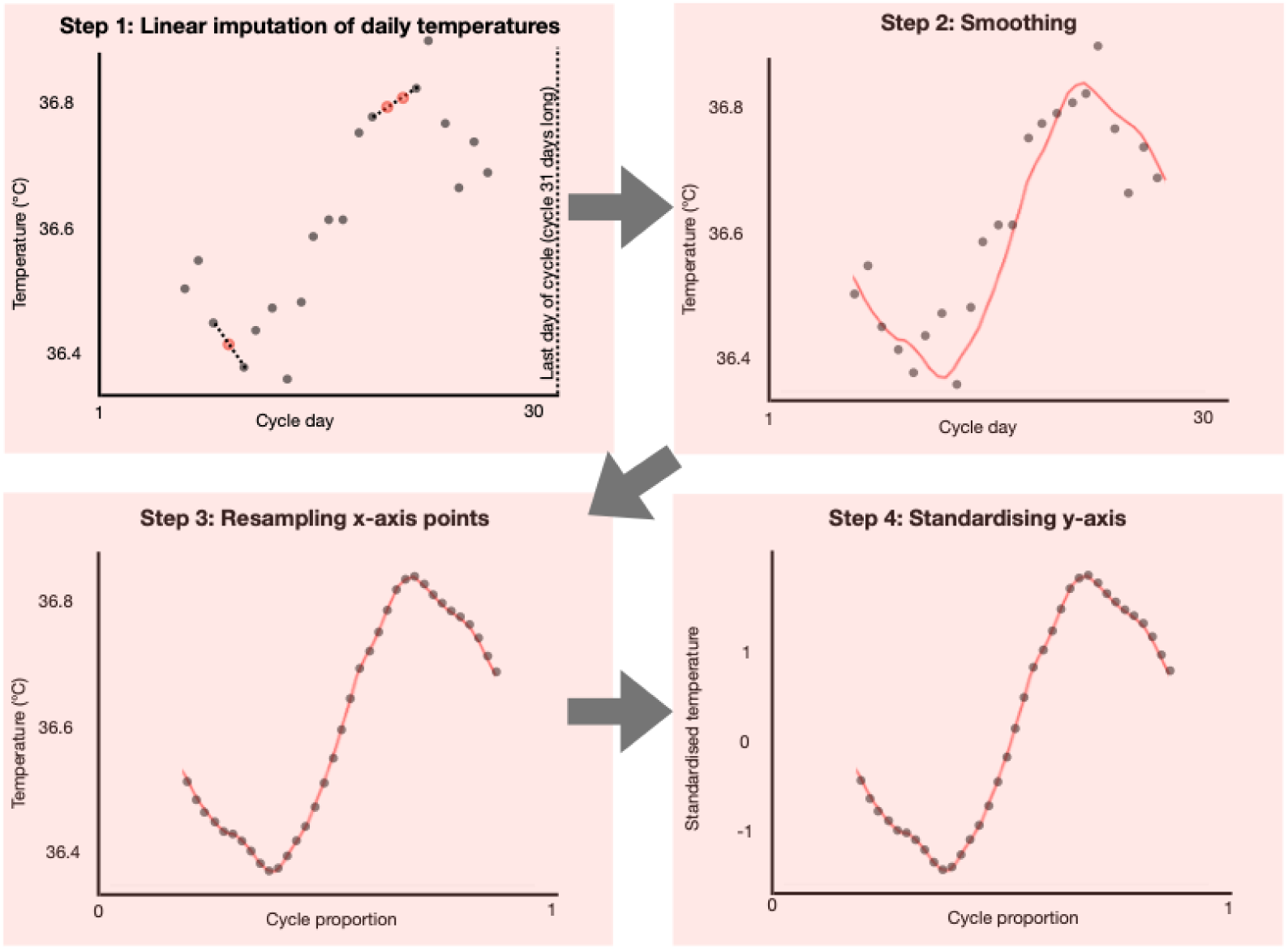
Illustration of cycle temperature data processing steps. Step 1: Nights with no mean temperature value were imputed with linear interpolation. Step 2: Savitzky-Golay smoothing (window=11, polynomial=2) was applied to reduce noise, resulting in the “processed-unstandardised” temperature data. Step 3: Conversion to a uniform timescale by resampling the cycle to have 51 data points. Step 4: Z-score standardisation (mean=0, variance=1) on the temperature values, resulting in the “processed-standardised” temperature data.

To account for the natural variability in menstrual cycle duration, we resampled each cycle to a fixed length of 51 data points, evenly spaced from the first to the last day of the cycle. This fixed-length representation provides a uniform time axis, ensuring all cycles are compared on a consistent relative timescale, both to one another and to the reference cycle, regardless of their original duration. Finally, temperature values for each cycle were standardised by subtracting the mean and dividing by the standard deviation [38,39]. The results of these transformations are illustrated in Figure 3 (Steps 3 and 4). This final output is hereafter referred to as ‘processed-standardised’ temperature data. As described in the following section, specific features are derived from either the processed-unstandardised or processed-standardised datasets, depending on the physiological characteristic being measured.

### Feature construction from temperature data

We developed two types of predictive models using features derived from the temperature data, using (a) cycles as examples (i.e. cycle-level), and (b) individuals as examples (i.e. user-level). To achieve this, we first extracted cycle-level features and then aggregated these to construct user-level features for each participant.

#### Cycle-level feature construction

We derived cycle-level features for three main categories: (a) features that directly describe cycle characteristics, (b) features derived using change-point detection that capture temperature changes in each cycle and their timing, and (c) features that compare each cycle to a typical cycle pattern. These are summarised in Table 1, which provides a full list of the cycle-level features.

**Table 1:** Feature set derived from the temperature data of each cycle, and used to predict PCOS.

| Feature | Description |
| --- | --- |
| **Features derived directly from processed-unstandardised cycle data** |  |
| Duration of menstrual phase | The number of days between the reported start of a cycle and the first temperature reading. This serves as a proxy for the length of the menstruation bleed days, as users are advised not to wear the device during this phase. |
| Length of cycle recorded period | The number of days between the first day of temperature recording and last day of temperature recording in a cycle. |
| Cycle length | The number of days between the first day of menstrual cycle (inclusive) at the beginning of a cycle, and the first day of next menstrual cycle (exclusive). |
| Proportion of recorded cycle period | The length of the cycle's recorded period divided by the cycle length |
| Length of the line of the cycle curve | The sum of the Euclidean distance between each consecutive pair of temperature points, divided by the number of days in the recorded period. This represents complexity of the curve, i.e. the extent that it varies from one day to the next. |
| Start day of three-day period with the largest rise in temperature | The three-day period within a cycle with the largest increase in temperature is identified, and the first day of this three-day period is used as a feature. |
| Temperature rise of three-day period with the largest rise | As above, with the difference in temperature between the first and third day included as a feature. |
| Start day of four-day period with the largest rise in temperature | The four-day period within a cycle with the largest increase in temperature is identified, and the first day of this four-day period is used as a feature. |
| Temperature rise of four-day period with the largest rise | As above, with the difference in temperature between the first and fourth day included as a feature. |
| **Features derived with change-point detection (applied to the processed-unstandardised cycle data)** |  |
| Day of temperature change | Day identified using change-point detection that splits the cycle into two sections that are most different. |
| Difference in temperature before and after detected temperature change | Mean temperature after day of temperature change, minus mean temperature before day of temperature change. |
| **Features comparing cycles with typical cycle pattern, derived with DTW (applied to the processed-standardised cycle data)** |  |
| DTW distance | Dynamic time warping distance between the cycle and the reference pattern, i.e. the extent to which DTW was able to warp the cycle to the reference pattern divided by the length of the data period in the cycle. |
| Length of optimal warping path | The amount of warping that DTW has performed to create the optimal alignment between the reference pattern and a given cycle. This is calculated as the number of cells in the DTW matrix, discounting the 1-to-many matches at either end, divided by the length of the data period in the cycle (see Section E of Multimedia Appendix 1). |
| Nadir Day | The day on the normalised cycle (from the processed-standardised cycle data) that is warped to the lowest temperature of the reference. |
| Peak Day | The day on the normalised cycle (from the processed-standardised cycle data) that is warped to the highest temperature of the reference. |
| Nadir Temperature | Cycle temperature (from the processed-standardised cycle data) on DTW nadir day. |
| Peak Temperature | Cycle temperature (from the processed-standardised cycle data) on DTW peak day. |
| Time to peak | The difference (number of days) between the nadir day and peak day on the on the normalised cycle |
| Difference between the nadir and peak temperatures | The difference between the standardised nadir and peak temperatures. |

PCOS: Polycystic ovary syndrome; DTW: Dynamic time warping

##### Features describing cycle characteristics derived using the processed, unstandardised cycle data

We included cycle length and derived features that describe the temperature data within the cycle [24,40]. These features encompass the number of days with recorded temperatures and the proportion of those days relative to the cycle length. Additionally, we derived features that aim to describe the timing and magnitude of the temperature surge accompanying ovulation, such as the largest temperature rise over a three-day period and the day index at which this rise began.

##### Features derived using change-point detection applied to the processed, unstandardised cycle data

We used change-point detection to mathematically partition each cycle into two distinct phases [41,42]. We then derived the day index of the detected change point, along with the difference in mean temperature before and after the change.

##### Features from comparing cycles with a typical temperature pattern, derived using the processed, standardised temperature data

Given the established physiological temperature changes across the menstrual cycle, we aimed to establish a standardised reference pattern derived from the cycles of participants who reported no known fertility issues and then extract features (for use in prediction models) that describe how each cycle deviates from this reference. We identified 25 individuals who stated they were using OvuSense for cycle monitoring and did not report having a fertility condition. Of these, we identified 3 individuals with suitable temperature data (including having cycles with sufficient data and typical cycle length) for generating a reference cycle. We used Dynamic Time-Warping Barycentre Averaging [43] to first derive person-specific reference patterns for each individual, and then to derive a single reference cycle from these. Full details of this process are provided in Section D of Multimedia Appendix 1.

Individual cycles were compared to the reference pattern using Dynamic Time Warping (DTW). The DTW algorithm computes an optimal alignment between sequence elements by warping (compressing or stretching) segments of the user cycle to match the reference. This method is particularly effective for menstrual cycle data, where physiological trajectories may follow similar patterns but occur with varying timing or durations [44,45]

After performing DTW alignment, we calculated the DTW distance to quantify the similarity between the reference and user cycles. We also measured the length of the optimal warping path, which indicates the total phase adjustment required by the algorithm to achieve the closest possible alignment. Using the warped indices, we identified the cycle nadir and peak (corresponding to the reference’s lowest and highest temperature points) and extracted their respective standardised temperatures. Finally, we computed the phase index difference between the nadir and peak points, and the magnitude of the temperature shift between them [14,46,47]. A detailed technical description of the nadir and peak identification process is provided in section E of the Multimedia Appendix 1.

#### User-level feature construction

We derived user-level features by calculating the minimum, maximum, median, and range of the cycle-level features across the three selected cycles for each participant in our sample. This approach captures the distribution and intra-individual variability of temperature-based physiological markers over time. To further account for heterogeneity between a participant’s cycles, we calculated the DTW distance and the length of the optimal warping path (as described above for the cycle-level features) for each unique pair of an individual’s three cycles. This results in three pairwise comparisons per individual. We then calculated the minimum, maximum, median, and range of these pairwise DTW distances and warping path lengths, incorporating them as additional features. The complete set of aggregated features for this stage is provided in Section F of Multimedia Appendix 1.

### Machine Learning for PCOS Identification

We stratified the participants in our dataset into 5 cross-validation (CV) folds, maintaining a consistent proportion of PCOS cases in each fold. This fold assignment was applied to both cycle-level and user-level predictions. This ensured that (a) all cycles for a given user were in the same fold for cycle-level prediction to reduce the risk of data leakage, and (b) users were in the same fold for both the cycle-level and user-level predictions, allowing for direct comparison of performance across folds.

We evaluated Logistic Regression (LR), Support Vector Machines (SVM), and Random Forest (RF) machine learning algorithms for predicting PCOS. We implemented a nested CV strategy on the stratified data to ensure robust evaluation. This approach involved two layers of cross-validation. The outer CV was used for performance estimation and consisted of 5-fold CV using the 5 folds described above. The inner CV (within the training set of each outer loop iteration) also used 5 folds and was used for hyperparameter optimisation using a grid search (where hyperparameters with the highest mean F1-score across this inner CV were selected). The hyperparameter search for LR included identifying the optimal regularisation strategy by testing L1, L2, and elastic net penalties. Details of other hyperparameters considered are provided in Section G of Multimedia Appendix 1. For each outer CV iteration, the model was trained using the locally optimal hyperparameters identified by the inner CV. The resulting models were then evaluated solely on the corresponding test fold of the outer CV, thereby ensuring the test data was never used in any part of the training process.

Model performance was assessed using the Area Under the Receiver Operating Characteristic Curve (AUC-ROC). To evaluate the reliability of the predicted probabilities, we assessed model calibration on the outer test sets using the Brier Score and calibration (reliability) curves. Calibration slope and intercept were calculated by fitting a logistic regression model to the predicted probabilities against the true labels, providing quantitative measures of miscalibration. Calibration curves were generated using 12 bins, and 100 bootstrap resamples were used to derive the 95% confidence intervals.

#### Assessing feature importance

We used SHAP (Shapley Additive exPlanations) values to evaluate feature importance. This method assesses each feature’s contribution to a model’s prediction by evaluating the average marginal contribution of that feature across all possible feature coalitions. We visualise feature importance using beeswarm plots, aggregating SHAP values from the held-out test sets of the five outer cross-validation folds to compare model interpretability across the different classifier types.

#### Assessment of PCOS prediction using available questionnaire data

In addition to predicting PCOS using temperature data, we also explored developing a prediction model using relevant available questionnaire data to serve as a comparison. However, due to the lack of important PCOS features, such as the presence of excess body hair in the questionnaire data, we relegate these analyses to the Multimedia Appendix (Section H).

All modelling and analyses were performed using Python 3.11.0, and all code is available at https://github.com/MRCIEU/PMOS-prediction-from-body-temperature/. For commercial enquiries, please contact the University of Bristol.

## Results

### Summary of the participant sample

Table 2 summarises the basic demographics of our participant sample.

**Table 2:**
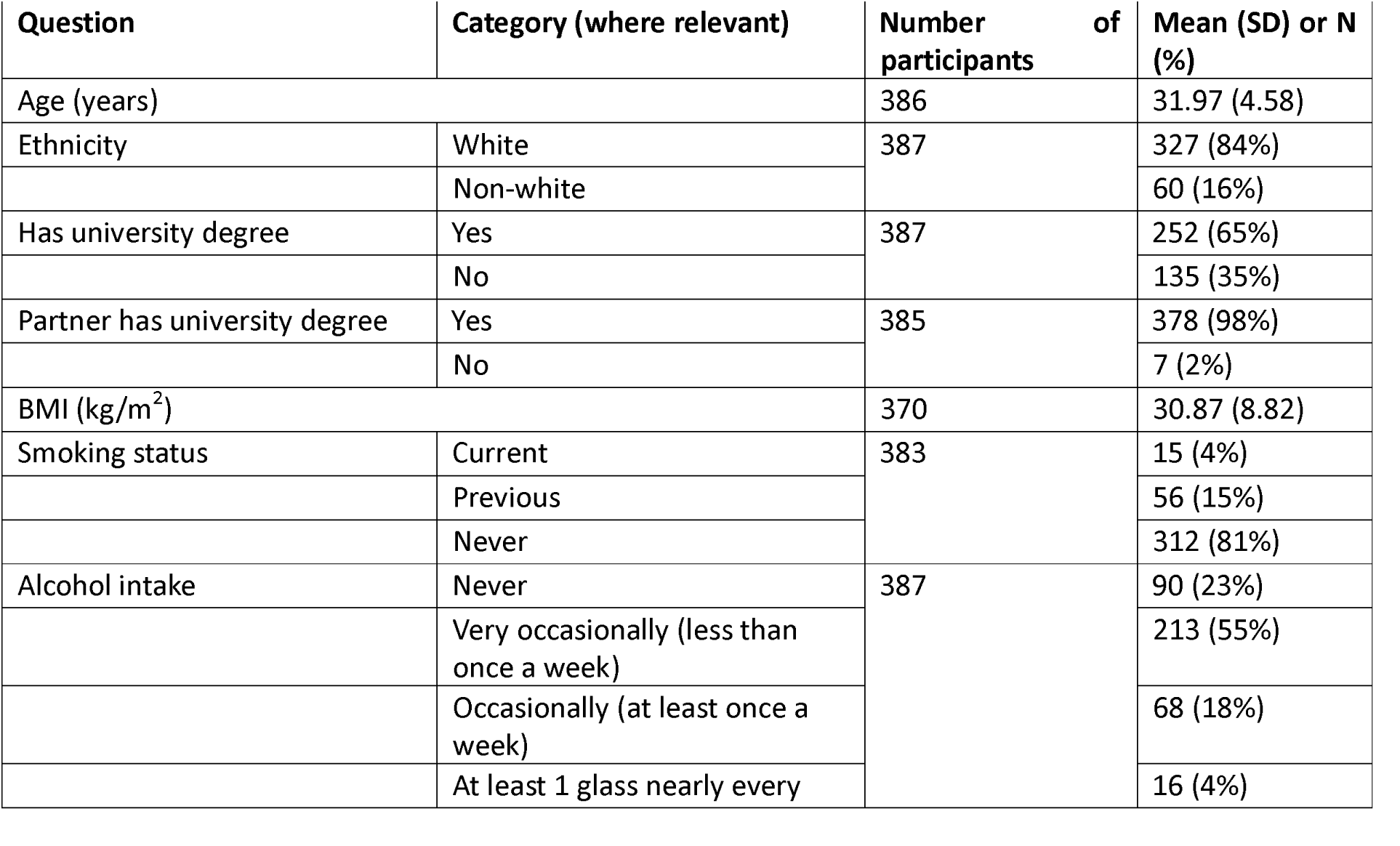
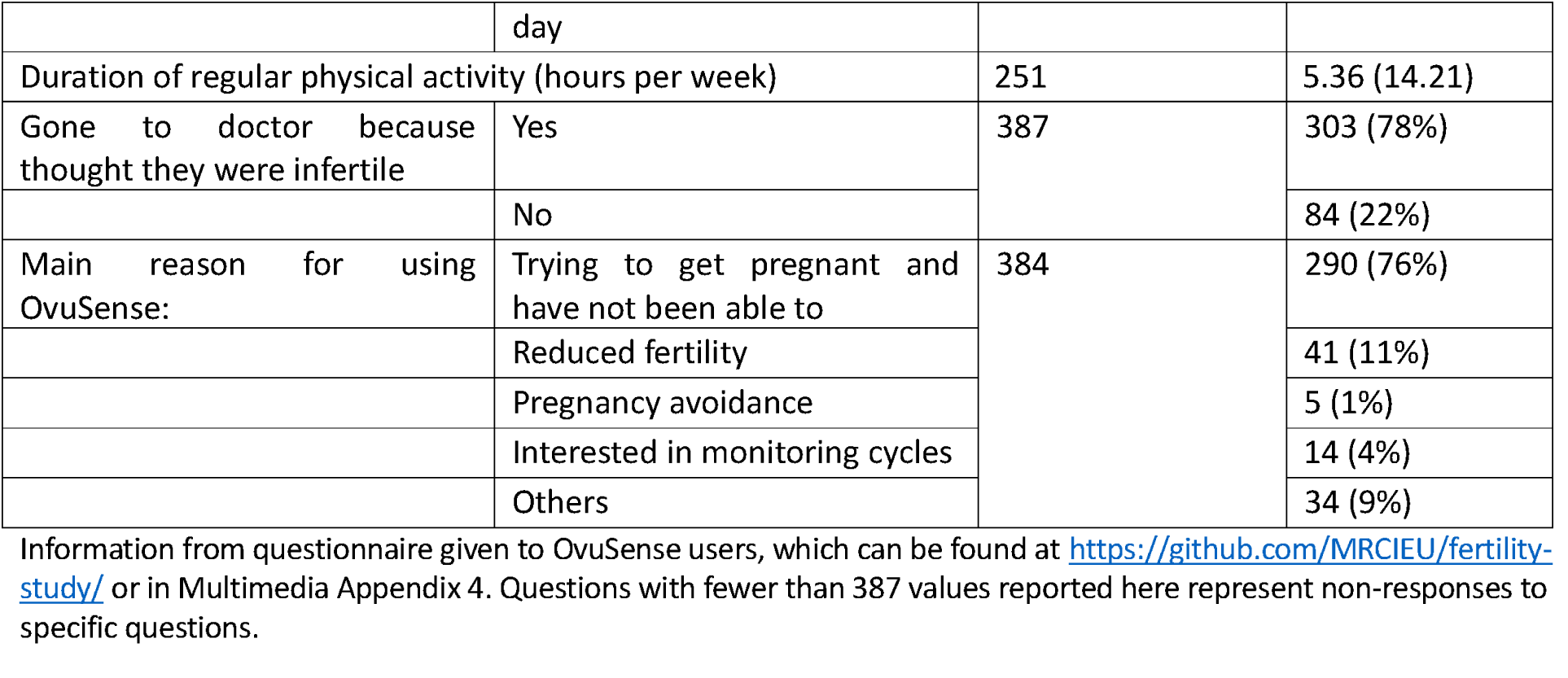
Summary of the participant sample.

| Question | Category (where relevant) | Number of participants | Mean (SD) or N (%) |
| --- | --- | --- | --- |
| Age (years) |  | 386 | 31.97 (4.58) |
| Ethnicity | White | 387 | 327 (84%) |
|  | Non-white |  | 60 (16%) |
| Has university degree | Yes | 387 | 252 (65%) |
|  | No |  | 135 (35%) |
| Partner has university degree | Yes | 385 | 378 (98%) |
|  | No |  | 7 (2%) |
| BMI (kg/m<sup>2</sup>) |  | 370 | 30.87 (8.82) |
| Smoking status | Current | 383 | 15 (4%) |
|  | Previous |  | 56 (15%) |
|  | Never |  | 312 (81%) |
| Alcohol intake | Never | 387 | 90 (23%) |
|  | Very occasionally (less than once a week) |  | 213 (55%) |
|  | Occasionally (at least once a week) |  | 68 (18%) |
|  | At least 1 glass nearly every day |  | 16 (4%) |
| Duration of regular physical activity (hours per week) |  | 251 | 5.36 (14.21) |
| Gone to doctor because thought they were infertile | Yes | 387 | 303 (78%) |
|  | No |  | 84 (22%) |
| Main reason for using Ovusense: | Trying to get pregnant and have not been able to | 384 | 290 (76%) |
|  | Reduced fertility |  | 41 (11%) |
|  | Pregnancy avoidance |  | 5 (1%) |
|  | Interested in monitoring cycles |  | 14 (4%) |
|  | Others |  | 34 (9%) |

Information from questionnaire given to Ovusense users, which can be found at https://github.com/MRCIEU/fertility-study/ or in Multimedia Appendix 4. Questions with fewer than 387 values reported here represent non-responses to specific questions.

Given our sample is a small subset of the 53,928 previous and existing OvuSense users, we sought to evaluate whether it is a representative subsample. The mean age at first temperature record for OvuSense users in our sample was 31.97 (SD = 4.58) years, compared with 31.97 (SD = 5.12) for the 27,595 customers who provided their date of birth and were not in our sample. Our sample used OvuSense for an average of 9.60 cycles (SD = 8.02), compared with 9.45 cycles (SD = 8.02) for the 32,593 users who had at least one cycle but were not in our sample. Also, the average cycle length in our sample was 36.89 (SD = 19.19), compared with 50.20 (SD = 87.75) for those not in our sample.

### Predicting PMOS using cycle- and user-level temperature features

Our models used 19 and 84 features for the cycle-level and user-level prediction, respectively. Correlations of the cycle-based features are provided in Figure S6 of Multimedia Appendix 2, and their distributions are shown in Figure S7 of Multimedia Appendix 2.

Figure 4 shows the ROC curves for the cycle- and user-level machine learning models, and Table 3 presents our prediction evaluation results (see Figures S9 and S10 of Multimedia Appendix 2 for results for each CV fold). For models using cycle-level features, the mean AUC across folds ranged from 0.64 (SD = 0.02) for logistic regression to 0.68 (SD = 0.04) for random forests. For the models with user-level features, the mean AUC ranged from 0.65 (SD = 0.07) for logistic regression to 0.70 (SD = 0.04) for random forests.

**Figure 4:**
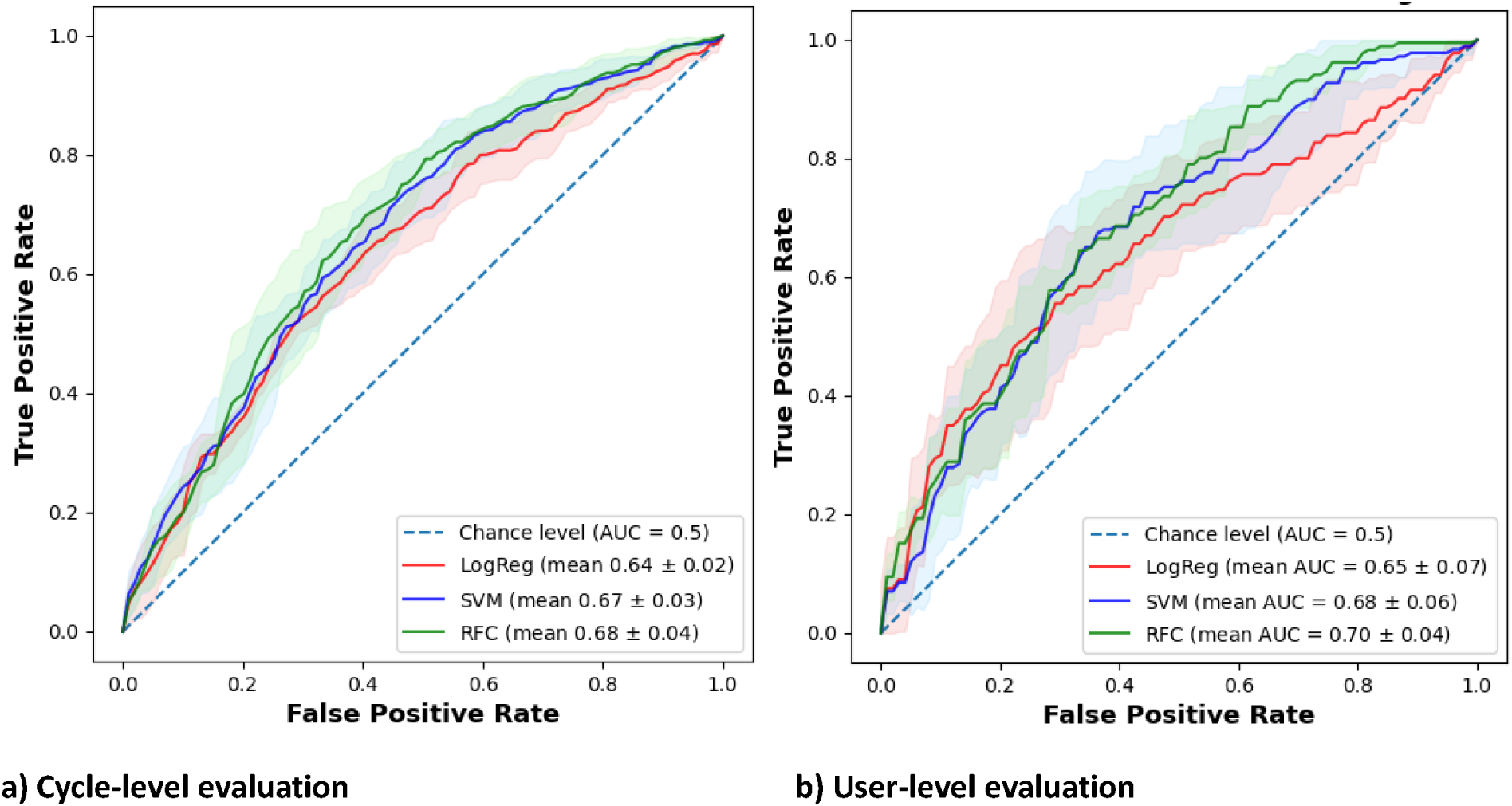
Mean ROC curves and confidence intervals from 5-fold cross-validation. ROC curves averaged vertically

**Table 3:** Results of PMOS prediction using cycle-level and user-level features.

|  | Logistic<br>Regression | Support Vector<br>Machine | Random Forest |
| --- | --- | --- | --- |
| **Cycle-level prediction** |  |  |  |
| Mean (SD) AUC across 5-fold CV | 0.64 (0.02) | 0.67 (0.03) | 0.68 (0.04) |
| Mean (SD) Brier Score across 5-fold CV | 0.241 (0.008) | 0.229 (0.006) | 0.226 (0.012) |
| Calibration slope (95% CI) <sup>1</sup> | 1.05 (0.69, 1.40) | 0.87 (0.70, 1.05) | 0.83 (0.68, 1.00) |
| Calibration intercept (95% CI) <sup>1</sup> | -0.02 (-0.14, 0.10) | 0.01 (-0.11, 0.13) | 0.02 (-0.11, 0.14) |
| **User-level prediction** |  |  |  |
| Mean (SD) AUC across 5-fold CV | 0.65 (0.07) | 0.68 (0.06) | 0.70 (0.04) |
| Mean (SD) Brier Score across 5-fold CV | 0.242 (0.024) | 0.228 (0.023) | 0.219 (0.012) |
| Calibration slope (95% CI) <sup>1</sup> | 0.47 (0.22, 0.79) | 0.81 (0.64, 1.09) | 0.88 (0.69, 1.15) |
| Calibration intercept (95% CI) <sup>1</sup> | -0.01 (-0.22, 0.15) | -0.04 (-0.25, 0.12) | -0.01 (-0.22, 0.16) |

AUC: Area under the ROC curve; SD: standard deviation; CV: cross-validation; CI: confidence interval. <sup>1</sup> Calibration slope and intercept are derived by concatenating predictions across the 5 cross-validation folds, and CIs are generated with bootstrapping. Calibration curves are provided in Figures S13-S18 in Multimedia Appendix 2.

### Assessment of calibration

The mean Brier scores for the cycle-level models ranged from 0.226 (SD=0.012) for RF to 0.241 (SD=0.008) for LR. For the user-level models, the Brier scores ranged from 0.219 (SD=0.012) for RF to 0.242 (SD=0.024 for LR. Overall, the models were reasonably well calibrated, though confidence intervals were wide. For example, the user-level RF model (that had the highest mean AUC overall) had a calibration slope of 0.88 (95% CI: 0.69, 1.15) and an intercept of -0.01 (-0.22, 0.16). Only the calibration slope confidence interval for the LR user-model did not include one, and all calibration intercept confidence intervals included zero.

### Assessment of Feature Importance

At the cycle-level, cycle length consistently emerged as the most important predictor, ranking among the top three most impactful features across all three model types. The direction of SHAP values indicates that longer cycle lengths are predictive of having a PCOS diagnosis. Furthermore, the length of the cycle recorded period was recognised as an important feature by the SVM and RF models, ranking among the top three features for both models. The importance of other features varied across the different models. At the user level, feature importance was more model-dependent, resulting in less consensus across algorithms. Features derived from DTW distance (comparing the cycles with the reference pattern) had high importance for the LR and SVM models, with the range and maximum DTW distance ranking among the top three predictors for both. For the RF model, features based on cycle length were the main contributors, with the maximum, medium and range of cycle length all ranked in the top three. Beeswarm plots for both the cycle and user-level are provided in Figures S25-S30 of Multimedia Appendix 2.

## Discussion

### Principal findings

In this study, we sought to evaluate whether body temperature captured overnight could be used to predict PMOS (referred to as PCOS at the time this study was conducted) using machine learning algorithms. For this early-stage evaluation, we used a sample of users of a device that measures temperature vaginally, who self-reported their PMOS diagnosis via a questionnaire. The primary finding of this study is that body temperature does have predictive potential for identifying individuals with PMOS, with mean AUC-ROCs across CV-folds of 0.64 or higher for all algorithms and temperature feature sets. We assessed prediction on both the cycle- and user-level to assess the added value of capturing heterogeneity across cycles (through aggregating cycle-level features into user-level features). However, discrimination performance was similar for both these feature sets, suggesting little added value of capturing heterogeneity across cycles. Our analysis of feature importance found cycle length was a key predictor in both the cycle- and user-level analyses, consistent with the established clinical symptoms of PMOS [5]. We used a novel approach to derive features comparing individuals’ cycles to a reference cycle (derived using the DTW time-series approach) that represented a typical cycle pattern in those without a fertility condition, and found these were also predictive across a number of algorithms/feature-sets.

### Related work

To our knowledge, only one previous study has examined whether it may be possible to predict PMOS using body temperature [33]. In that study, 51 participants were recruited and asked to wear the OvulaRing device (which is worn vaginally, both day and night) for one menstrual cycle. The authors assessed the predictive performance of three temperature-based measures – total daily amplitude over the whole cycle, first day of temperature rise, and duration of lower temperature phase – individually with PMOS. In contrast, our study, in a much larger sample, looks at predicting PMOS using real-world data, and assesses the extent that a model could be developed to predict PMOS passively i.e. among those who are not using a device explicitly for this purpose (in our sample the majority of our sample are using the OvuSense device to aid conception). While Goeckenjan et al. assessed three characteristics, we applied time-series analysis approaches, including DTW and change-point detection, to derive features capturing a range of characteristics of the temperature curve’s shape and dynamics, that then contributed to differing extents to the predictive performance of our models.

### Strengths and limitations

A key strength of our study is the assessment of PMOS prediction using real-world data, as this would be the intended deployment context of such an approach where consumer wearable sensors could potentially enable earlier detection by suggesting that those deemed to be high risk consult with a medical professional. Our evaluation uses data with substantial amounts of missing data that would also be commonplace if such a model was deployed, though the amount and specific patterns of missing data would depend on the device and its userbase. In our case, missing data occurred both due to the intended use of the device (e.g. OvuSense users are instructed to not wear the device whilst menstruating) and also due to user-driven factors where the device is not worn on some nights or cycles start dates are not indicated in the smartphone app (needed to segment the temperature data into cycles). In other devices, such as those worn on the wrist or a finger, missing data may be due primarily to user-driven factors as these devices are intended to be worn continuously. Devices worn across the whole cycle (e.g. smart rings) may help improve data completeness but may also provide a less accurate proxy of BBT due to being worn peripherally rather than internally [24].

To deal with the challenges of the real-world data used in this study, we developed a rigorous data filtering pipeline. This pipeline removed erroneous data (e.g. implausible temperatures values), cycles with insufficient amounts of data, and data with inconsistencies (e.g. records predating reported cycle start dates). Our study used several time-series analysis approaches to derive novel features that may be predictive of PMOS. The approaches used are informed by the known menstrual cycle temperature patterns and specifically designed to capture deviations from this. We used a reasonable sample size (N = 387), much larger than the only previous study that has evaluated the use of temperature data to predict PMOS (described above) [33].

A key limitation of this study is the use of a self-reported PMOS outcome variable that is defined retrospectively such that it is likely that most participants reporting PMOS will have been diagnosed with this condition prior to or during the period when they used the OvuSense device. This assumes that the temperature patterns before a PMOS diagnosis are the same as after diagnosis, which may not be the case as treatments for PMOS may then impact a participant’s menstrual cycle patterns captured through body temperature. Future work should use a sample who do not have PMOS at the time of device wear and use the recorded temperature data to predict a subsequent PMOS diagnosis. Our PMOS variable may also suffer from misclassification bias for a number of reasons. First, we defined PMOS case status using two questions – one asking whether participants had received a diagnosis from a doctor, and one asking for the conditions they took medication for. We did this as the medication question was much earlier in the questionnaire, and it is possible that some may have not answered the later questions as thoroughly or may have skipped them. This may mean that some participants were classified as a PMOS case who did not have a doctor diagnosis, although the numbers were small (7 of the 192 cases stated PMOS as a condition for which they took regular medication but did not report a PMOS diagnosis from a doctor). It is also likely that some of those assigned as a PMOS control have undiagnosed PMOS.

In this study we used a binary outcome variable – PMOS case versus control – and did not consider diagnoses for other fertility conditions. Our temperature-derived features may be capturing signatures of general anovulatory dysfunction rather than PMOS-specific signals. Future work should evaluate the extent that PMOS can be distinguished from other fertility conditions, for example using a multi-label classification approach (where each individual can be assigned more than one outcome category). We used only body temperature to predict PMOS but other physiological modalities measured (in addition to temperature) by other digital health devices, such as heart rate and physical activity, may help improve predictive performance.

We used a small sample of the overall OvuSense customer base, consisting of those who responded to the study questionnaire. This sample may not be representative of the overall set of customers such that our ability to predict PMOS in OvuSense users more widely may be different. Furthermore, OvuSense is used primarily by those who have been trying to conceive but have not been able to, and the extent that PMOS can be predicted from temperature data may be different in a more general population (e.g. among those using a more general purpose and widely used device such as a smartwatch). We derived features by first deriving a reference cycle from participants but could only use data from three participants who met our eligibility criteria for this; using more examples may improve these features and our ability to predict PCOS using them. The size of our sample also limited our ability to assess calibration, due to the number of participants in each fold. In order to assess this, we combined the predicted scores across all folds and assessed calibration performance of this as a single dataset, which assumes that the scores generated from the models generated in the different CV iterations are comparable. However, confidence intervals on the calibration curve were fairly wide and so larger samples are needed to assess calibration with greater statistical power.

## Conclusion

A prediction approach using body temperature is not intended to replace established clinical diagnostics, such as the Rotterdam criteria. Instead, its potential value is through being able to passively detect PMOS in the general population, among those using a device that measures body temperature who may not be seeking a diagnosis of PMOS. In this study, we take the first step towards establishing the feasibility of such an approach. We have demonstrated that using machine-learning models to predict PMOS from nightly body temperature measures does have potential. Much further work is needed, including assessing this approach in users of more general-purpose devices that are more widely used (targeting those in early adulthood for early intervention) and with PMOS measured prospectively. Qualitative work is also needed, engaging clinicians (gynaecologists, endocrinologists) and menstruating individuals who wear digital devices, in order to explore acceptability and potential challenges, implementation (e.g. how and when predictions may be presented to users), and implications for existing care pathways.

## Supporting information

Multimedia appendix 1

Multimedia appendix 2

Multimedia appendix 3

Multimedia appendix 4

Multimedia appendix 5

## Data Availability

The data used in study are not publicly available because they are owned by viO HealthTech.

## Funding

This research was carried out in the MRC Integrative Epidemiology Unit at the University of Bristol (MC_UU_00032/3, MC_UU_00011/4, MC_UU_00032/5) and supported by the Cancer Research UK Integrative Cancer Epidemiology Programme (C18281/A29019). This work was also supported by the National Institute for Health and Care Research (NIHR) Bristol Biomedical Research Centre (grant no: NIHR 203315). LACM received funding from a University of Bristol Vice-Chancellor’s Fellowship. The views expressed are those of the author(s) and not necessarily those of the MRC, Cancer Research UK, the NIHR or the Department of Health and Social Care. The funders had no role in study design, data collection and analysis, decision to publish, or preparation of the manuscript.

## Conflicts of interest

ViO HealthTech Ltd provided the data used in this study and was involved in participant recruitment and data collection, but was not involved in the design, analysis, or execution of the research, and had no role in the decision to publish or in the preparation of the manuscript. TRG receives funding from Biogen, GSK and Roche for unrelated research. DAL and TRG receive funding from Novartis for unrelated research.

## Acknowledgements

We are extremely grateful to all the OvuSense users who took part in this study.

## Abbreviations

AUC-ROC: Area under the receiver operating curve
BBT: Basal Body Temperature
CI: Confidence interval
CV: Cross-validation
DTW: Dynamic time warping
LR: Logistic regression
PCOS: Polycystic ovary syndrome
PMOS: Polyendocrine metabolic ovarian syndrome
RF: Random forest
SD: Standard deviation
SHAP: Shapley Additive exPlanations
SVM: Support Vector Machine

## Notes

### Author Declarations

Faculty of Health Sciences Research Ethics Committee of University of Bristol gave ethical approval for this work (Reference: 4527).

## References

1. Teede HJ, Khomami MB, Morman R, Laven JSE, Joham AE, Costello MF, Patil M, Rees DA, Berry L, Cree MG, Zhao H, Norman RJ, Dokras A, Piltonen T. Polyendocrine metabolic ovarian syndrome, the new name for polycystic ovary syndrome: a multistep global consensus process. The Lancet Elsevier; 2026 May 12; doi: 10.1016/S0140-6736(26)00717-8

2. Bulsara J, Patel P, Soni A, Acharya S. A review: Brief insight into Polycystic Ovarian syndrome. Endocrine and Metabolic Science Elsevier; 2021 Jun 30;3:100085. doi: 10.1016/J.ENDMTS.2021.100085

3. World Health Organization (WHO). Polycystic ovary syndrome. World Health Organisation Factsheet on Polycystic Ovarian Syndrome. 2023. Available from: https://www.who.int/news-room/fact-sheets/detail/polycystic-ovary-syndrome [accessed Nov 13, 2023]

4. Gibson-Helm M, Teede H, Dunaif A, Dokras A. Delayed Diagnosis and a Lack of Information Associated With Dissatisfaction in Women With Polycystic Ovary Syndrome. J Clin Endocrinol Metab J Clin Endocrinol Metab; 2017 Feb 1;102(2):604–612. PMID:27906550

5. Teede Helena, Tay Chau Thien, Laven Joop, Dokras Anuja, Moran Lisa, Piltonen Terhi, Costello Michael, Boivin Jacky, Redman Leanne, Boyle Jacqueline, Norman Robert, Mousa Aya, Joham Anju. International Evidence-based Guideline for the assessment and management of polycystic ovary syndrome. Melbourne, Australia; 2023 Sep. doi: 10.26180/24003834.v1

6. Wolf WM, Wattick RA, Kinkade ON, Olfert MD. Geographical Prevalence of Polycystic Ovary Syndrome as Determined by Region and Race/Ethnicity. International Journal of Environmental Research and Public Health 2018, Vol 15, Multidisciplinary Digital Publishing Institute; 2018 Nov 19;15(11). PMID:30463276

7. Deswal R, Narwal V, Dang A, Pundir CS. The Prevalence of Polycystic Ovary Syndrome: A Brief Systematic Review. J Hum Reprod Sci J Hum Reprod Sci; 2020 Oct 1;13(4):261–271. PMID:33627974

8. Rotterdam ESHRE/ASRM. Revised 2003 consensus on diagnostic criteria and long-term health risks related to polycystic ovary syndrome (PCOS). Human Reproduction 2004 Jan 1;19(1):41–47. doi: 10.1093/humrep/deh098

9. NICE. Diagnosis | Diagnosis | Polycystic ovary syndrome | CKS | NICE. 2024. Available from: https://cks.nice.org.uk/topics/polycystic-ovary-syndrome/diagnosis/diagnosis/ [accessed Feb 29, 2024]

10. Copp T, Hersch J, Muscat DM, McCaffery KJ, Doust J, Dokras A, Mol BW, Jansen J. The benefits and harms of receiving a polycystic ovary syndrome diagnosis: a qualitative study of women’s experiences. Hum Reprod Open 2019 Mar 1;2019(4). doi: 10.1093/hropen/hoz026

11. Zhu TY, Rothenbühler M, Hamvas G, Hofmann A, Welter JE, Kahr M, Kimmich N, Shilaih M, Leeners B. The accuracy of wrist skin temperature in detecting ovulation compared to basal body temperature: Prospective comparative diagnostic accuracy study. J Med Internet Res JMIR Publications Inc.; 2021 Jun 1;23(6):e20710. PMID:34100763

12. Reed BG, Carr BR. The Normal Menstrual Cycle and the Control of Ovulation. Endotext MDText.com, Inc.; 2018 Aug 5; PMID:25905282

13. Su H, Yi Y, Wei T, Chang T, Cheng C. Detection of ovulation, a review of currently available methods. Bioeng Transl Med Wiley; 2017 Sep 1;2(3):238–246. PMID:29313033

14. Baker FC, Siboza F, Fuller A. Temperature regulation in women: Effects of the menstrual cycle. Temperature: Multidisciplinary Biomedical Journal Taylor & Francis; 2020 Jul 2;7(3):226. PMID:33123618

15. World Health Organization (WHO). Family Planning—A Global Handbook for Providers. 4th ed. Geneva: WHO; 2022. ISBN:978-0-9-9920370-5

16. Sato D, Ikarashi K, Nakajima F, Fujimoto T. Novel Methodology for Identifying the Occurrence of Ovulation by Estimating Core Body Temperature During Sleeping: Validity and Effectiveness Study. JMIR Form Res 2024 Jul 5;8:e55834. doi: 10.2196/55834

17. Händel P, Wahlström J. Digital contraceptives based on basal body temperature measurements. Biomed Signal Process Control Elsevier; 2019 Jul 1;52:141–151. doi: 10.1016/J.BSPC.2019.04.019

18. Goodale BM, Shilaih M, Falco L, Dammeier F, Hamvas G, Leeners B. Wearable Sensors Reveal Menses-Driven Changes in Physiology and Enable Prediction of the Fertile Window: Observational Study. J Med Internet Res 2019 Apr 18;21(4):e13404. doi: 10.2196/13404

19. Wang Y, Park J, Zhang CY, Jukic AMZ, Baird DD, Coull BA, Hauser R, Mahalingaiah S, Zhang S, Curry CL. Performance of algorithms using wrist temperature for retrospective ovulation day estimate and next menses start day prediction: a prospective cohort study. Human Reproduction 2025 Mar 1;40(3):469–478. doi: 10.1093/humrep/deaf005

20. Shilaih M, Goodale BM, Falco L, Kübler F, De Clerck V, Leeners B. Modern fertility awareness methods: wrist wearables capture the changes in temperature associated with the menstrual cycle. Biosci Rep Portland Press Ltd; 2018 Nov 30;38(6):BSR20171279. PMID:29175999

21. Maijala A, Kinnunen H, Koskimäki H, Jämsä T, Kangas M. Nocturnal finger skin temperature in menstrual cycle tracking: ambulatory pilot study using a wearable Oura ring. BMC Womens Health BMC Womens Health; 2019 Nov 29;19(1). PMID:31783840

22. Papaioannou S, Aslam M, Al Wattar BH, Milnes RC, Knowles TG. User’s acceptability of OvuSense: a novel vaginal temperature sensor for prediction of the fertile period. J Obstet Gynaecol J Obstet Gynaecol; 2013 Oct;33(7):705–709. PMID:24127960

23. Regidor PA, Kaczmarczyk M, Schiweck E, Goeckenjan-Festag M, Alexander H. Identification and prediction of the fertile window with a new web-based medical device using a vaginal biosensor for measuring the circadian and circamensual core body temperature. Gynecol Endocrinol Gynecol Endocrinol; 2018 Mar 4;34(3):256–260. PMID:29082805

24. Hurst BS, Davies K, Milnes RC, Knowles TG, Pirrie A. Novel Technique for Confirmation of the Day of Ovulation and Prediction of Ovulation in Subsequent Cycles Using a Skin-Worn Sensor in a Population With Ovulatory Dysfunction: A Side-by-Side Comparison With Existing Basal Body Temperature Algorithm and Vaginal Core Body Temperature Algorithm. Front Bioeng Biotechnol Frontiers Media S.A.; 2022 Mar 4;10. doi: 10.3389/FBIOE.2022.807139/FULL

25. Uchida Y, Izumizaki M. The use of wearable devices for predicting biphasic basal body temperature to estimate the date of ovulation in women. J Therm Biol Pergamon; 2022 Aug 1;108:103290. PMID:36031211

26. Wark JD, Henningham L, Gorelik A, Jayasinghe Y, Hartley S, Garland SM. Basal Temperature Measurement Using a Multi-Sensor Armband in Australian Young Women: A Comparative Observational Study. JMIR Mhealth Uhealth JMIR Publications Inc.; 2015 Dec 1;3(4). PMID:26441468

27. Weiss G, Strohmayer K, Koele W, Reinschissler N, Schenk M. Confirmation of human ovulation in assisted reproduction using an adhesive axillary thermometer (femSense®). Front Digit Health Frontiers Media S.A.; 2022 Sep 19;4. doi: 10.3389/FDGTH.2022.930010/FULL

28. Chauhan P, Patil P, Rane N, Raundale P, Kanakia H. Comparative Analysis of Machine Learning Algorithms for Prediction of PCOS. 2021 International Conference on Communication information and Computing Technology (ICCICT) Mumbai: Institute of Electrical and Electronics Engineers (IEEE); 2021. p. 1–7. doi: 10.1109/ICCICT50803.2021.9510128

29. Nasim S, Almutairi MS, Munir K, Raza A, Younas F. A Novel Approach for Polycystic Ovary Syndrome Prediction Using Machine Learning in Bioinformatics. IEEE Access Institute of Electrical and Electronics Engineers Inc.; 2022;10:97610–97624. doi: 10.1109/ACCESS.2022.3205587

30. Mridul HA, Ahsan N, Alam SS, Afrose S, Sultana Z, Tanvir M, Kafi M. Polycystic Ovary Syndrome (PCOS) Disease Prediction Using Traditional Machine Learning and Deep Learning Algorithms. International Journal of Computer Information Systems and Industrial Management Applications 2024 Jul 10;16(3):25–25. Available from: https://cspub-ijcisim.org/index.php/ijcisim/article/view/712 [accessed Apr 9, 2026]

31. Rahman MM, Islam A, Islam F, Zaman M, Islam MR, Alam Sakib MS, Hasan Babu HM. Empowering early detection: A web-based machine learning approach for PCOS prediction. Inform Med Unlocked Elsevier; 2024 Jan 1;47:101500. doi: 10.1016/J.IMU.2024.101500

32. Kottarathil P. Polycystic ovary syndrome (PCOS). [Data set] Kaggle. 2020. Available from: https://www.kaggle.com/datasets/prasoonkottarathil/polycystic-ovary-syndrome-pcos [accessed Apr 9, 2026]

33. Goeckenjan M, Schiwek E, Wimberger P. Continuous Body Temperature Monitoring to Improve the Diagnosis of Female Infertility. Geburtshilfe Frauenheilkd Geburtshilfe Frauenheilkd; 2020 Jul 1;80(7):702–712. PMID:32675832

34. Kong Q, Siauw T, Bayen AM. Python Programming and Numerical Methods. A Guide for Engineers and Scientists. Python Programming and Numerical Methods. 2020. Available from: http://www.sciencedirect.com:5070/book/9780128195499/python-programming-and-numerical-methods [accessed Jul 31, 2023]ISBN:9780128195499

35. Morrisset D, Santamaria S, Hadden R, Emberley R. Implications of data smoothing on experimental mass loss rates. Fire Saf J Elsevier; 2022 Jul 1;131:103611. doi: 10.1016/J.FIRESAF.2022.103611

36. Staggs JEJ. Savitzky–Golay smoothing and numerical differentiation of cone calorimeter mass data. Fire Saf J Elsevier; 2005 Sep 1;40(6):493–505. doi: 10.1016/J.FIRESAF.2005.05.002

37. Zhang H, Dong Y. Advances in Brillouin dynamic grating in optical fibers and its applications. Prog Quantum Electron Pergamon; 2023 Jan 1;87:100440. doi: 10.1016/J.PQUANTELEC.2022.100440

38. Caldwell JA, Niro PJ, Farina EK, McClung JP, Caron GR, Lieberman HR. A Z-score based method for comparing the relative sensitivity of behavioral and physiological metrics including cognitive performance, mood, and hormone levels. PLoS One Public Library of Science; 2019 Aug 1;14(8):e0220749. PMID:31415596

39. Carré A, Klausner G, Edjlali M, Lerousseau M, Briend-Diop J, Sun R, Ammari S, Reuzé S, Alvarez Andres E, Estienne T, Niyoteka S, Battistella E, Vakalopoulou M, Dhermain F, Paragios N, Deutsch E, Oppenheim C, Pallud J, Robert C. Standardization of brain MR images across machines and protocols: bridging the gap for MRI-based radiomics. Scientific Reports 2020 10:1 Nature Publishing Group; 2020 Jul 23;10(1):1–15. PMID:32704007

40. Harris HR, Titus LJ, Cramer DW, Terry KL. Long and irregular menstrual cycles, polycystic ovary syndrome, and ovarian cancer risk in a population-based case-control study. Int J Cancer 2017 Jan 15;140(2):285–291. doi: 10.1002/ijc.30441

41. Aminikhanghahi S, Cook DJ. A survey of methods for time series change point detection. Knowl Inf Syst 2017 May 8;51(2):339–367. doi: 10.1007/s10115-016-0987-z

42. Truong C, Oudre L, Vayatis N. Selective review of offline change point detection methods. Signal Processing 2020 Feb;167:107299. doi: 10.1016/j.sigpro.2019.107299

43. Petitjean F, Ketterlin A, Gançarski P. A global averaging method for dynamic time warping, with applications to clustering. Pattern Recognit Pergamon; 2011 Mar 1;44(3):678–693. doi: 10.1016/J.PATCOG.2010.09.013

44. Hebbrecht K, Stuivenga M, Birkenhäger T, Morrens M, Fried EI, Sabbe B, Giltay EJ. Understanding personalized dynamics to inform precision medicine: a dynamic time warp analysis of 255 depressed inpatients. BMC Med BioMed Central Ltd; 2020 Dec 1;18(1):1–15. PMID:33353539

45. Samara E, Laperre B, Kieokaew R, Temmer M, Verbeke C, Rodriguez L, Magdalenić J, Poedts S. Dynamic Time Warping as a Means of Assessing Solar Wind Time Series. Astrophys J IOP Publishing; 2022 Mar 15;927(2):187. doi: 10.3847/1538-4357/AC4AF6

46. Kicińska AM, Stachowska A, Kajdy A, Wierzba TH, Maksym RB. Successful Implementation of Menstrual Cycle Biomarkers in the Treatment of Infertility in Polycystic Ovary Syndrome—Case Report. Healthcare Multidisciplinary Digital Publishing Institute (MDPI); 2023 Feb 1;11(4). PMID:36833150

47. Steward K, Raja A. Physiology, Ovulation And Basal Body Temperature. StatPearls StatPearls Publishing; 2023 Jul 17; PMID:31536292

