## Supplementary material for "A digital health approach for identifying polyendocrine metabolic ovarian syndrome using machine learning and body temperature": Multimedia appendix 1

**Awoniran et al.**

### MULTIMEDIA APPENDIX 1 – SUPPLEMENTARY TEXT

**Section A: The cycle validation phases**
The flowchart in Figure S1 in Multimedia Appendix 2 illustrates the stepwise attrition of the raw dataset to reach the final valid cycles. The following rationales were applied to ensure data integrity and physiological relevance:

Step 1: Daily Records Validation - To minimise noise from ambient temperature adjustment, the initial 25 minutes of each nightly recording session were discarded. A nightly record was only considered valid if it contained at least 10 readings within the physiological range (35.0°C to 40.0°C). Cycles with no valid nights were excluded to ensure a minimum baseline of longitudinal data.

Step 2: Chronological Consistency - Cycles with null start dates or start dates recorded after the first temperature reading were removed. These chronological errors would invalidate the calculation of cycle-level features.

Step 3: Cycle Termination (Subsequent Cycle Requirement) - To accurately determine the total duration and conclusion of a menstrual cycle, each included cycle was required to have a confirmed subsequent cycle start date.

Step 4: Physiological Plausibility - Cycles shorter than 10 days or longer than 366 days were excluded. Very short cycles may be due to a user indicating menstruation on multiple non-consecutive days for the same menstrual period, whereas very long cycles may be most likely due to a user not indicating the start of a menstrual period such that our defined cycles actually contain multiple menstrual cycles.

Step 5: Signal Stability - We excluded cycles that had fewer than 10 valid nightly records prior to a gap of 10 or more consecutive missing readings. This filter prevents the analysis of cycles with extreme initial sparsity where interpolation would lack sufficient physiological grounding.

Step 6: Minimum Data Density - Finally, we required 40% of cycle days to have a temperature value. This ensures that time-series features (such as Dynamic Time Warping and change-point detection) have sufficient signal density to accurately reflect the dynamics of the true basal body temperature (BBT) curve throughout the cycle.

The final validated dataset consists of 3,906 cycles from 499 individual users.

**Section B: PCOS Outcome Groupings of the Analytical Sample**

Using the defined classification criteria, 192 participants were classified as cases of PCOS and 195 as controls. The cases included individuals with self-reported physician diagnoses (n = 185) and those identified solely through taking PCOS-specific medications (n = 7). The control group (n = 195) included participants who had never sought medical advice for infertility (n = 78), as well as those who had been clinically evaluated for fertility issues but were diagnosed with unrelated conditions (n = 61). Among others, these diagnoses included other ovarian problems, endometriosis, partner/sperm-related issues, and thyroid disorders (see Table S1 in Multimedia Appendix 3 for the full list). Note that these categories were not mutually exclusive, as some participants reported multiple diagnoses. Additionally, individuals who underwent evaluation but received no specific diagnosis (n = 49) or did not indicate a diagnosis (n = 7) were included in the control group.

**Section C: Savitzky-Golay filtering applied to an example cycle to smooth cycle temperatures (with outliers)**
The Savitzky-Golay filter was used to smooth the raw temperature data, enabling precise identification of key points in the menstrual cycle, such as the nadir and peak temperatures. Correctly choosing the filter's parameters, particularly the window length and polynomial order, was essential to strike the right balance between removing noise and maintaining the true physiological trend.

1. **Window Length = 21, Polynomial Order = 2:** Using a large window length resulted in over-smoothing of the cycle data. As shown in the provided plot, this caused a shift in the nadir and peak days, which are critical for accurate analysis. This setting was deemed unsuitable because it averaged out important physiological information.
2. **Window Length = 5, Polynomial Order = 2:** Conversely, using a small window length resulted in trend overfitting. As shown in the provided plot, the smoothed curve was heavily influenced by individual temperature outliers, failing to accurately reflect the underlying physiological cycle.
3. **Window Length = 11, Polynomial Order = 4:** As shown in the provided plot, this combination of parameters also resulted in trend overfitting. Although the window length was moderate, the higher polynomial order caused the smoothed curve to follow the noise and outliers too closely, thereby failing to accurately represent the underlying physiological trend of the cycle.
4. **Window Length = 11, Polynomial Order = 2: The optimal parameters**, a window length of 11 and a polynomial order of 2, were chosen because they effectively removed noise from outliers while maintaining the true physiological trend. The smoothed curve accurately reflected the cycle’s nadir and peak, which are essential for subsequent feature extraction and analysis (red solid line in Figure S2). This selection of parameters successfully balanced noise reduction with the preservation of key features.

Figure S2 in Multimedia Appendix 2 shows the results of smoothing with various parameters.

**Section D: Deriving the reference cycle pattern**

The processing pipeline for deriving the reference cycle is discussed as follows-

<u>Data selection</u>

The reference cycle was derived using OvuSense temperature data through a systematic process. First, we identified participants who indicated that their main reason for using OvuSense was “I am just interested in monitoring my cycles.” Other responses to this question include “I have been trying to get pregnant and have not been able to” and “I have reduced fertility, and it might help”, both suggesting fertility issues. Another option, “I am trying to avoid becoming pregnant,” was considered but deemed infeasible because the few individuals who chose this option often stopped using the device after the post-ovulatory temperature rise, resulting in insufficient data for the full cycle. 25 participants selected “I am just interested in monitoring my cycles”. We then filtered the participants and cycles using the following steps.

- Step 1. Of the 25 participants, we excluded 16 who reported fertility issues (e.g., PCOS or amenorrhoea), leaving 9 participants.
- Step 2. Of the 9 participants, four were excluded due to missing temperature data, leaving five individuals. From this group, we identified 31 cycles containing temperature data and a subsequent cycle.
- Step 3. One cycle with more than seven days of missing temperature data at the start (i.e., from the beginning of the cycle to the first night with recorded temperature) was removed, leaving 30 cycles.
- Step 4. 17 cycles with missing data at the end were excluded. This reduced the number of cycles to 13 and the number of participants to three.
- Step 5. Finally, cycles with durations of 25-35 days were selected. This excluded one cycle.

This process yielded 12 high-quality cycles across the three individuals.

<u>Imputation for Missing Values</u>

Missing temperature data points were imputed using linear interpolation. When the first data point of a cycle was missing, we applied circular imputation, using the temperature from the last recorded point in that cycle as the proxy value for the missing first data point. This method assumes that, during healthy, regular cycles, the BBT at the end of menstruation returns to a value similar to that observed at the onset of the cycle (physiological periodicity). To ensure this assumption was valid, our data selection process specifically required that cycles have no more than 7 days of initial missingness (step 3 of data selection) and a complete data record at the cycle's conclusion (Step 4). Linear interpolation was then used to fill any remaining gaps.

<u>Data Smoothing</u>

To remove noise and subtle outliers from the raw temperature data, the cycles were smoothed using Savitzky-Golay filter, with parameters selected for optimal performance as discussed in Section C of this appendix. This step was crucial for isolating the underlying physiological trend of the BBT curve from random fluctuations.

<u>Data Standardization</u>

After smoothing, the data were Z-score standardised to a consistent scale. Each cycle's temperatures were centred and scaled by their mean and standard deviation, respectively. This step ensured that all cycles, regardless of their individual temperature range, were comparable, enabling a uniform analysis of their shape and temporal dynamics.

<u>User-Specific Averaging</u>

To mitigate the influence of individual users with many cycles, a user-specific average cycle was initially calculated for each selected participant. This was done using Dynamic Time Warping barycenter averaging (DBA). DBA is a technique for averaging time series that addresses temporal shifts while preserving the overall shape of the curve.

Final Reference Cycle

The final reference cycle was then obtained by averaging the user-specific averages. This second application of the DBA ensured a typical population-level trend, rather than being skewed by the number of cycles contributed by any single user. This comprehensive approach yielded a highly representative reference cycle, serving as a critical benchmark for the DTW-based features in our predictive models.

The reference cycle derived using this process is shown in Figure S3 in Multimedia Appendix 2.

**Section E: Identification of the Cycle Nadirs and Peaks using DTW**

DTW Application

Dynamic Time Warping (DTW) is employed as a flexible alignment algorithm to address the inherent non-uniformity of physiological processes across cycles (e.g., cycle day 15 for one participant may not align physiologically with that same day for another). The algorithm calculates an optimal non-linear warping path between a specific user cycle and the reference cycle. This path maps each data point in the user cycle to its most similar temporal and morphological counterpart in the reference cycle.

Given two sequences as $A = [a_1, a_2 \dots a_n]$ and $B = [b_1, b_2 \dots b_n]$, the cumulative distance $D(A_i, B_j)$ for the optimal alignment is computed using the following recurrence relation:

$$D(A_i, B_j) = \delta(a_i, b_j) + min \begin{cases} D(A_{i-1}, B_{j-1}) \\ D(A_i, B_{j-1}) \\ D(A_{i-1}, B_j) \end{cases} \quad \dots \quad Eqn\ i$$

The resulting warping and alignment cost between the reference cycle and an example cycle are illustrated in Figure S4 of Multimedia Appendix 2.

Nadir and Peak Determination

The nadir and peak of a user cycle are determined using established anchor points in the reference cycle. Because the reference cycle is derived from high-quality data, its nadir and peak days are stable and known. The DTW algorithm identifies which data points on the user cycle align optimally with these reference anchor points.

In the example cycle shown in Figure S5 of the Multimedia Appendix 2, the point aligning with the reference nadir is designated as the cycle nadir (yellow circle). Similarly, the point aligning with the reference peak is designated as the cycle peak (red circle). This shape-based approach ensures that features are robustly located based on the overall thermal-shift morphology, rather than on raw minimum or maximum values, which are often susceptible to noise or outliers. These identified days are subsequently used to derive key features, such as the magnitude of the thermal shift and the duration of the temperature rise.

Figure S5 in the Multimedia Appendix 2 illustrates the mapping process used to identify these critical physiological features.

**Section F: The user-level features**

To capture the complex physiological signatures and the internal consistency of a participant's menstrual cycles, we engineered a comprehensive set of 84 user-level features. These were derived using two distinct aggregation methods applied to the three randomly selected cycles for each participant.

1. Inter-Cycle Pairing Metrics (Stability Analysis)

The first set of features (Features 1–8) measures the internal consistency of a participant's cycles. For each participant, the three selected cycles were paired (i.e., Cycle 1 vs Cycle 2, Cycle 2 vs Cycle 3, and Cycle 1 vs Cycle 3). We used DTW to create alignments between these paired cycles and obtained their respective DTW distances and the lengths of the optimal warping paths. We then calculated the minimum, maximum, median, and range of these distances and lengths for each user. These metrics are critical for identifying the irregular, unpredictable patterns often associated with PCOS, as opposed to the more stable, repeatable patterns seen in healthy cycles.

2. Aggregated Cycle-Level Metrics (Physiological Dimensions)

The remaining features (Features 9–84) represent a user's overall profile. These were derived by aggregating the cycle-level metrics described in the main text across the three selected cycles by calculating their minimum, maximum, median, and range.

The complete list of the user-level features is shown in Table S2 in Multimedia Appendix 3.

**Section G: Hyperparameter tuning for the machine learning algorithms**

The hyperparameter grid for logistic regression includes solvers compatible with the specified regularisation penalties. Specifically, the *L1* penalty was exclusively paired with the *liblinear* and *saga* solvers, as the remaining gradient-based solvers, *newton-cg*, *lbfgs*, and *sag*, are incompatible with the non-differentiable *L1* penalty. Similarly, the elasticnet penalty was exclusively paired with the *saga* solver, which utilises proximal operators to handle the combined *L1* and *L2* penalties. The hyperparameter values tested for logistic regression are shown in Table S3 in Multimedia Appendix 3.

The support vector machine (SVM) hyperparameters were tuned across both linear and non-linear kernels. The *Radial Basis Function (RBF)* kernel, a non-linear kernel, was tested alongside the linear kernel to explore non-linear decision boundaries. The selection of *C* (regularisation) and *gamma* (kernel coefficient) was performed across a wide logarithmic range to test the model's flexibility and to penalise misclassification effectively. The hyperparameter values tested for support vector machines are shown in Table S4 in Multimedia Appendix 3.

The random forest classifier (RFC) hyperparameters were tuned to optimise the model's balance between variance reduction and computational cost. The number of trees (*n_estimators*) was tested up to 200 to ensure stable, low-variance predictions through ensemble aggregation. Maximum tree depth (*max_depth*) was varied, including the unconstrained option (None), to control the model complexity and mitigate overfitting. *min_samples_split* was also explored as a critical structural parameter. The *max_features* parameter was not explicitly tuned, and the model used the default setting, the square root of *n_features*, to select the number of features considered at each split, providing sufficient decorrelation between trees while maintaining computational efficiency. The hyperparameter values tested for random forest classifiers are shown in Table S5 in Multimedia Appendix 3.

**Section H: Methods and results for modelling with questionnaire features**

Selecting the questionnaire variables

We identified a broad range of categories from the NICE guidelines, specifically the ‘when to suspect’, ‘causes’, and ‘diagnosis’ PCOS information pages. These categories included (a) cycle regularity, (b) family history of PCOS, (c) diagnostic criteria for PCOS, and (d) evidence of ovulation disorders, such as amenorrhoea or pregnancy history. Since we aimed to develop a prediction tool useful for women before they attempt to conceive, we excluded pregnancy history. Only four women reported experiencing amenorrhoea, so we did not include this variable. The questionnaire did not collect any family history information, so we were unable to include variables in that category. We incorporated variables that are direct effects of PCOS, such as cycle regularity, but excluded more distant effects like diabetes, sleep disorders, and psychological conditions, including anxiety and depression, as our focus is on broadly applicable predictions, even for those who may not have yet experienced the consequences of PCOS.

We included variables characterising women’s menstrual cycle experiences. These include the age at first menstruation, as studies suggest that late onset of menarche may be associated with PCOS [1]. We also incorporated variables related to heavy or painful menstruation, as they are connected to gynaecological conditions [2,3]. Lastly, we incorporated body measurements, such as Body Mass Index (BMI), given the well-established association between obesity and PCOS [4]. Additionally, we considered smoking, as it is associated with poor reproductive outcomes, and women with PCOS are more likely to smoke than those without [5]. The final set of features included in our questionnaire-based model is:

- Cycle regularity: Regular periods, period in the last three months
- Cycle characteristics: Age menstruation started, heavy period, painful periods
- Other measures: BMI, smoking

Preprocessing the questionnaire-derived variables

BMI and age at which menstruation began were kept as numerical variables. The questions about cycle regularity, periods in the past three months, and smoking habits had responses of “Yes” and “No”; hence, we coded these responses as ones (yes) and zeros (no). The questions concerning cycle heaviness and pain during cycles had response options: “very”, “moderately”, “mildly”, and “not at all”; therefore, we coded these as ordinal variables ranging from 3 (very) to 0 (not at all). Unfortunately, the questionnaire did not ask for information about excessive hair growth, so we were unable to include it in our questionnaire-based prediction model.

Results

ROC curves are provided in Figures S11 and S12 in Multimedia Appendix 2, and calibration curves are provided in Figures S19-S24. Beeswarm plots are shown in Figures S31-S36. While discrimination performance was similar for the questionnaire-based and user-level temperature models, there was some evidence of improved performance when using a combined user-level and questionnaire-based feature set, with a higher AUC across all folds for LR and a higher or equal AUC across all folds for SVM and RF models. All models (combined across CV folds) for the questionnaire and combined user-level and questionnaire feature sets appeared reasonably well calibrated, though confidence intervals were wide (see Tables S8 and S9 in Multimedia Appendix 3 for Brier scores, and calibration slope and intercept estimates).

**Section I: The Feature distributions**

The cycle-level features

The distribution of the number of recorded cycles per user (Figure S7A in Multimedia Appendix 2) shows that most participants had fewer than 10 cycles, with frequency declining as cycle count increases. This suggests that many users utilise the device for short durations to achieve specific objectives, while a smaller cohort of long-term users forms the long tail of the distribution.

The *nadir day* and *peak day* follow approximately normal distributions (Figures S7B–C in Multimedia Appendix 2). As expected, the distribution of the *peak day* reflects that peak temperatures typically occur in the latter half of the cycle. *Time to peak* (Figure S7D in Multimedia Appendix 2) represents the typical duration for the thermal shift and is slightly right-skewed.

Temperature-specific metrics, including *Nadir temperature* and *Peak temperature* (derived by comparison to a reference cycle pattern), are centred around distinct physiological values (36.2°C and 36.8°C, respectively), demonstrating high consistency in the recorded data (Figures S7E–F in Multimedia Appendix 2). *Difference between the nadir and peak temperatures* (Figure S7G in Multimedia Appendix 2) is predominantly between 0.3°C and 0.6°C, aligning with the physiological expectation of a subtle but distinct thermal shift following ovulation [6,7].

The questionnaire features

The *BMI* distribution is right-skewed (Figure S8A in Multimedia Appendix 2). The age at which menstruation started is normally distributed, with most users reporting onset between ages 12 and 13, consistent with established physiological benchmarks (Figure S8B in Multimedia Appendix 2). Most participants identified as non-smokers and confirmed having had a menstrual period within the last three months (Figures S8C–D in Multimedia Appendix 2). The distribution of users reporting regular versus irregular cycles is similar (Figure S8E in Multimedia Appendix 2). Self-reported symptoms indicate that "Moderately" heavy periods are the most common experience (Figure S8F in Multimedia Appendix 2), while reported pain levels cluster around "Mildly" to "Moderately" painful, with fewer users reporting "Not at all" or "Very" painful periods (Figure S8G in Multimedia Appendix 2).

### References


1. Shukla A, Ashraf GM, Sudharsan V, Arora T, Rather KUI, Chowdhury S, Suri V, Joshi B, Bhattacharya PK, Agrawal S, Malhotra N, Sahay R, Jabbar PK, Nair A, Rozati R, Rashid H, Wani IA, Maan P, Gautam R. Trends of Age at Onset of Menarche Among Indian Women of Reproductive Age and Its Association with the Presence of PCOS and Related Features: A Multicentric Cross Sectional Study. Journal of Obstetrics and Gynecology of India Springer; 2025 Apr 1;75(Suppl 1):70–77. doi: 10.1007/S13224-024-01994-6/TABLES/5
2. James AH. Heavy menstrual bleeding: work-up and management. Hematology: the American Society of Hematology Education Program American Society of Hematology; 2016;2016(1):236. PMID:27913486
3. Petraglia F, Bernardi M, Lazzeri L, Perelli F, Reis FM. Dysmenorrhea and related disorders. F1000Res Faculty of 1000 Ltd; 2017;6:1645. PMID:28944048
4. Barber TM, Franks S. Obesity and polycystic ovary syndrome. Clin Endocrinol (Oxf) 2021 Oct 31;95(4):531–541. doi: 10.1111/cen.14421
5. Yang Y, Zhang H, Huang B-Y, Lu Y-H, Fukuzawa I, Yang S, Zhou L, Luo L, Wang C, Ding N, Li S, Shi L, Zhang H-L. Relationship between smoking, excessive androgen and negative emotions in women with polycystic ovary syndrome (PCOS). J Ovarian Res 2024 Oct 29;17(1):211. doi: 10.1186/s13048-024-01541-x
6. Su H, Yi Y, Wei T, Chang T, Cheng C. Detection of ovulation, a review of currently available methods. Bioeng Transl Med Wiley; 2017 Sep 1;2(3):238–246. PMID:29313033
7. Steward K, Raja A. Physiology, Ovulation And Basal Body Temperature. StatPearls StatPearls Publishing; 2023 Jul 17; PMID:31536292
