## Supplementary material for "A digital health approach for identifying polyendocrine metabolic ovarian syndrome using machine learning and body temperature": Multimedia appendix 2

**Awoniran et al.**

**MULTIMEDIA APPENDIX 2 - FIGURES**

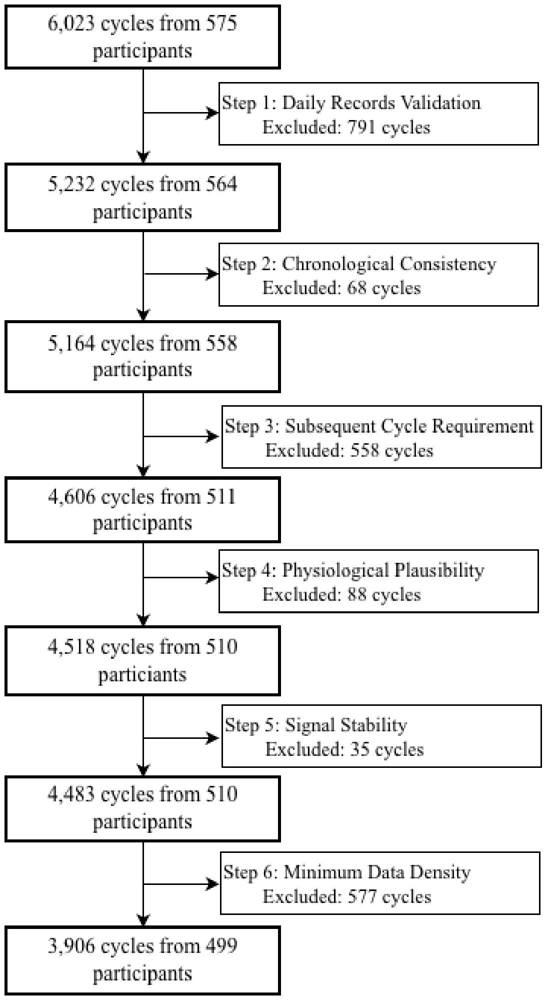


**Figure S1: Cycle Validation and Filtering Flowchart**. Flowchart of the cycle validation and filtering process. The diagram illustrates the six sequential steps taken to refine the raw OvuSense dataset into the final analytical sample, highlighting the number of cycles and users retained at each stage of quality control.

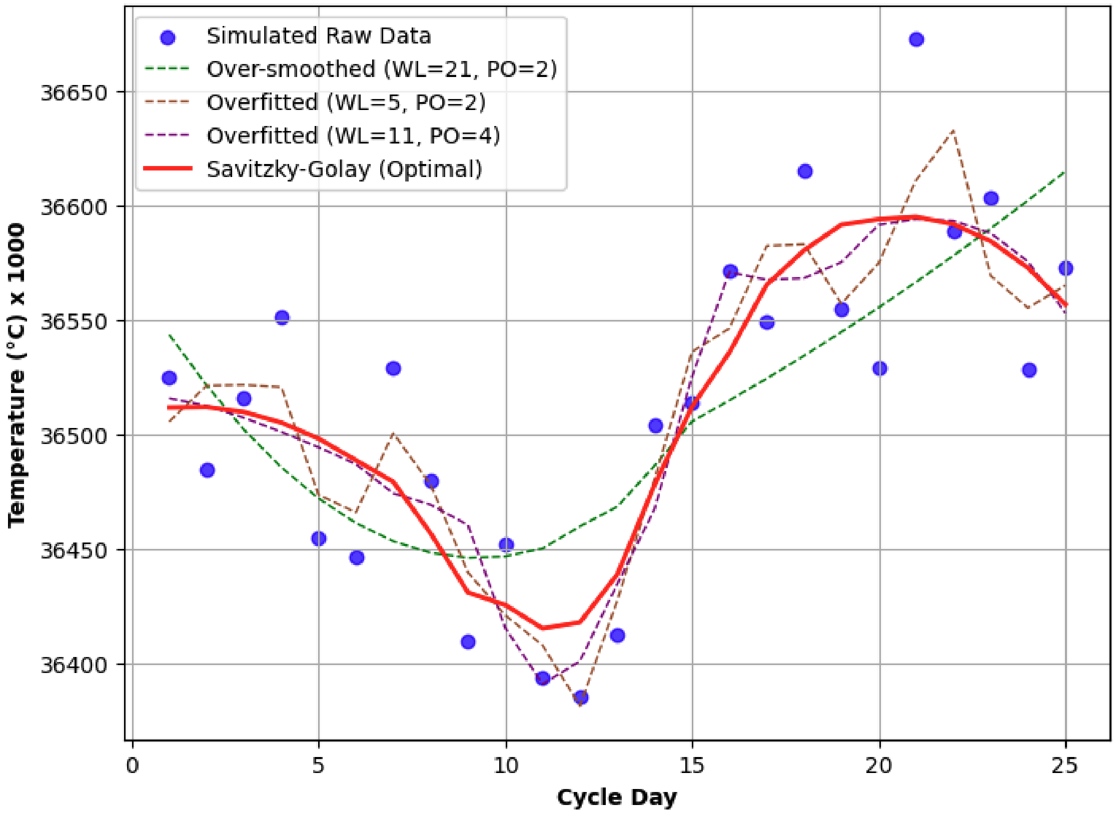


**Figure S2: Impact of Savitzky-Golay filter parameter selection on temperature smoothing.** A representative menstrual cycle is shown with simulated raw data (blue dots) and various smoothing configurations. Window Length (WL) and Polynomial Order (PO) are varied to demonstrate the trade-off between noise reduction and physiological accuracy. The optimal fit (solid red line; WL=11, PO=2) preserves the biphasic trend and identifies nadir and peak days. The over-smoothed fit (green dashed line; WL=21, PO=2) fails to capture the magnitude of thermal shifts, while overfitted fits (WL=5, PO=2; WL=11, PO=4) are negatively influenced by individual temperature outliers.

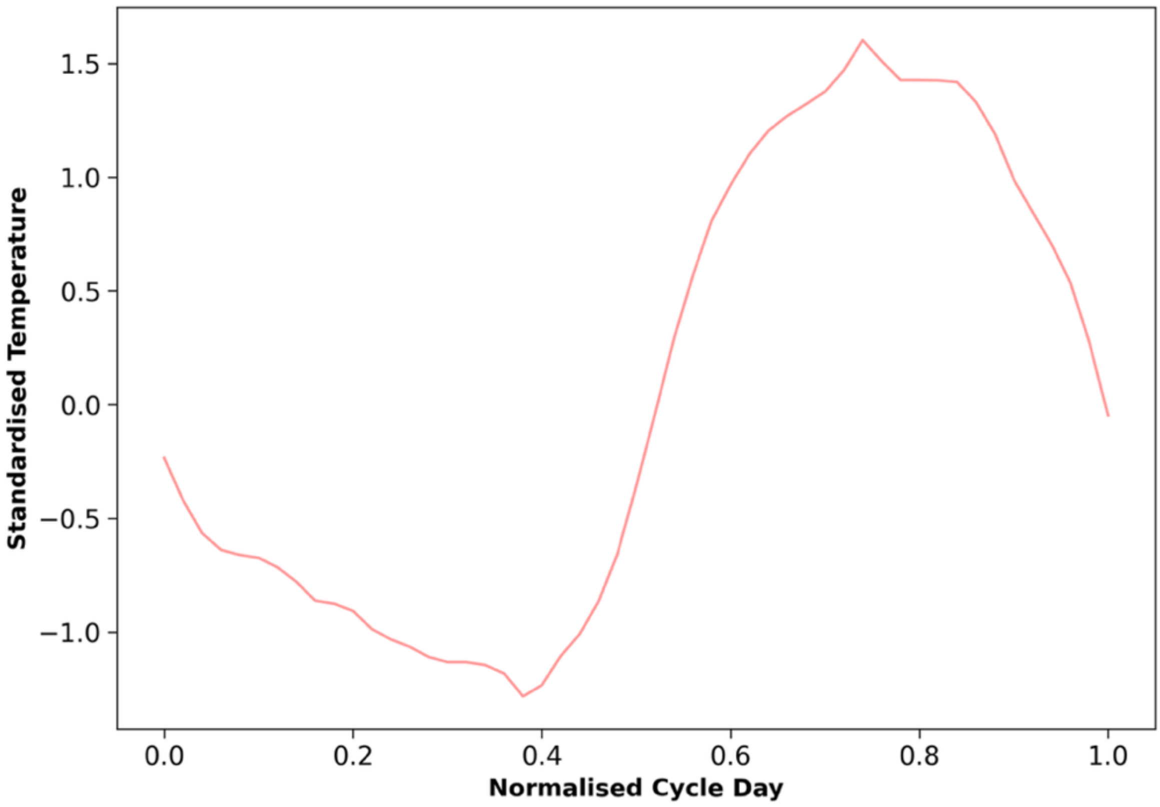


**Figure S3: The derived reference cycle pattern.** The plot shows the final reference cycle obtained through the systematic processing of high-quality cycles (n=12) from participants with no reported fertility issues. The x-axis represents the Normalised Cycle Day (scaled 0 to 1), and the y-axis shows the Standardised Temperature (Z-score). The biphasic trend was preserved using Dynamic Time Warping (DTW) barycenter averaging, which aligns temporal shifts across participants while maintaining the characteristic physiological shape of the basal body temperature (BBT) curve.

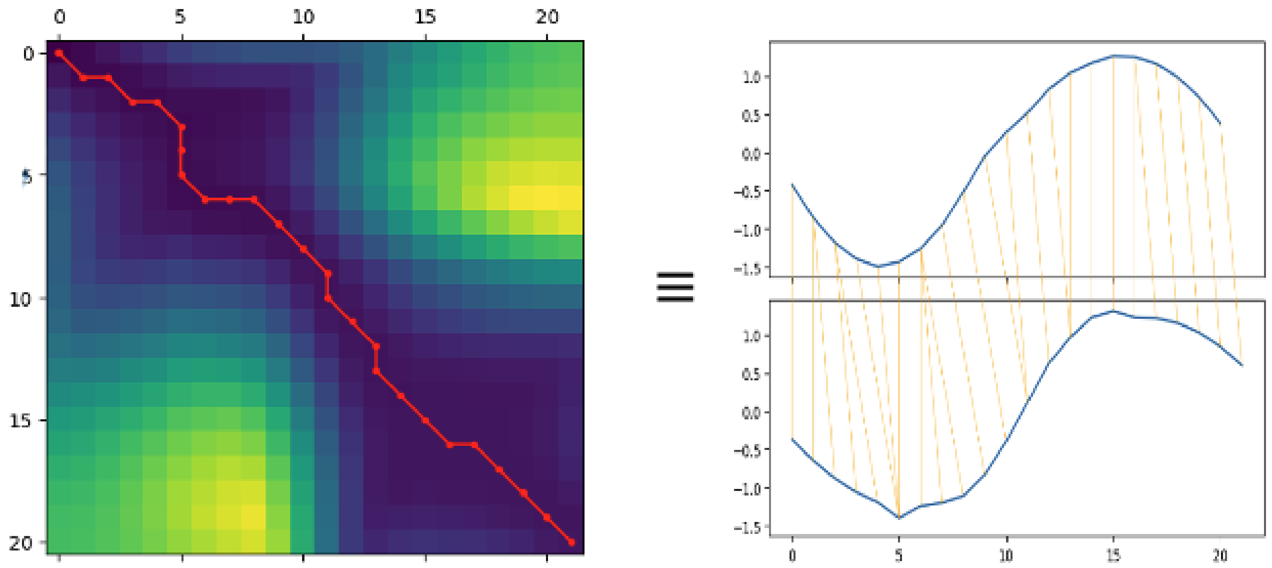

**Figure S4: Dynamic Time Warping (DTW) alignment logic**. Left - A cost matrix showing the optimal warping path (red line) between two sequences. The path minimises the cumulative distance between aligned points. Right - A visual representation of the resultant warping, where orange lines indicate how specific points in the temporal domain are mapped to one another to account for phase shifts and varying cycle lengths.

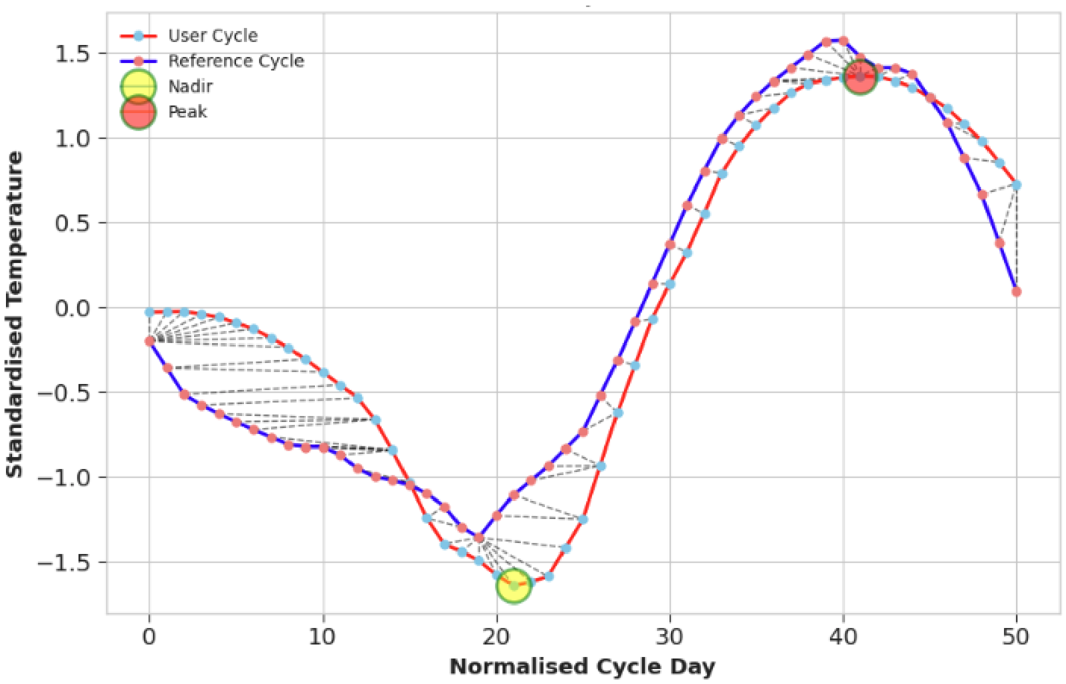


**Figure S5: Identification of cycle nadir and peak via reference mapping.** The figure demonstrates the alignment between a specific user cycle (red) and the reference cycle (blue). Dashed grey lines represent the DTW warping path. The user-specific nadir (yellow circle) and peak (red circle) are identified by their optimal alignment to the stable anchor points of the reference cycle, ensuring physiological features are captured despite individual variations in cycle timing.

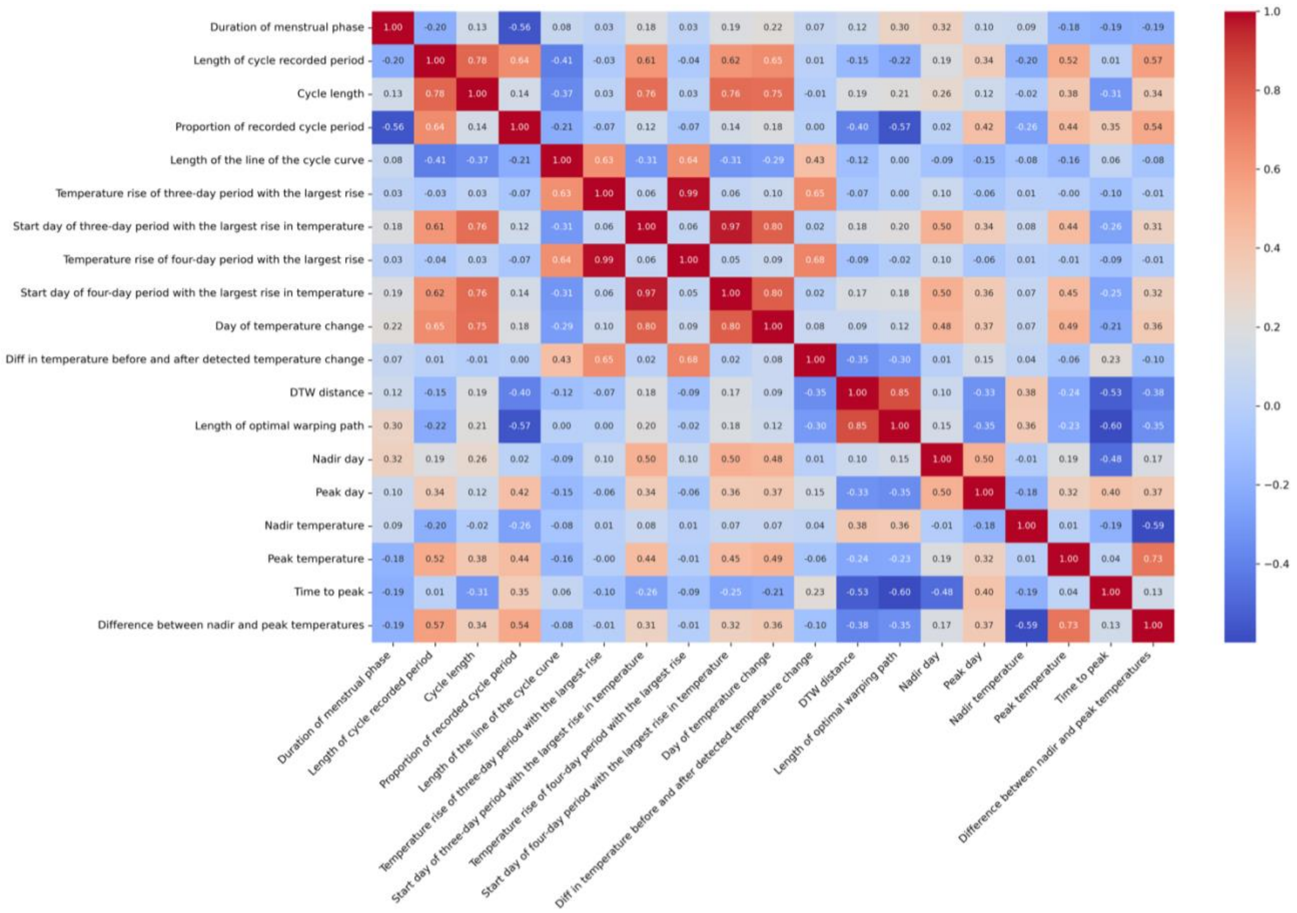


**Figure S6: Spearman correlation heatmap of cycle-level features.** The heatmap illustrates the monotonic relationships between the engineered features. The colour scale indicates the correlation coefficient ($\rho$), ranging from strong negative correlation (dark blue, -1.0) to strong positive correlation (dark red, +1.0). The correlation heatmap visually summarises the monotonic relationships between all features using Spearman's correlation ($\rho$). As expected, conceptually similar or related features are highly correlated. For example, *Length of cycle recorded period* is strongly positively correlated with both *Cycle length* and *Day of temperature change*. The negative correlation between *Proportion of recorded cycle period* and *Length of optimal warping path* indicates that more complete cycles generally require less warping to align with the reference. Many features, such as *Duration of the menstrual phase*, show weak or nearly zero correlations with other variables, indicating they provide unique, non-redundant information to the model.

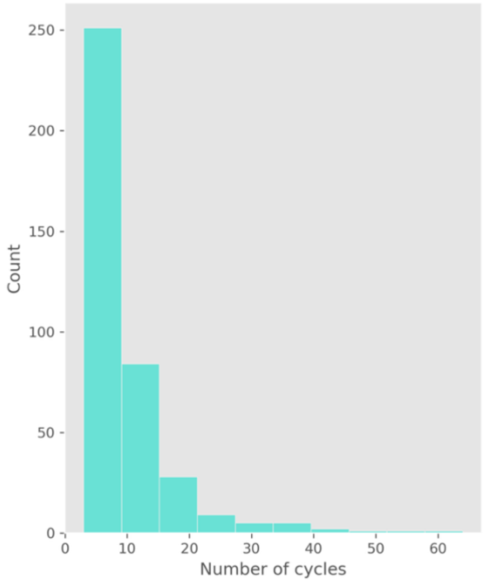
Count
0
50
100
150
200
250
0
10
20
30
40
50
60
Number of cycles

**(A)**

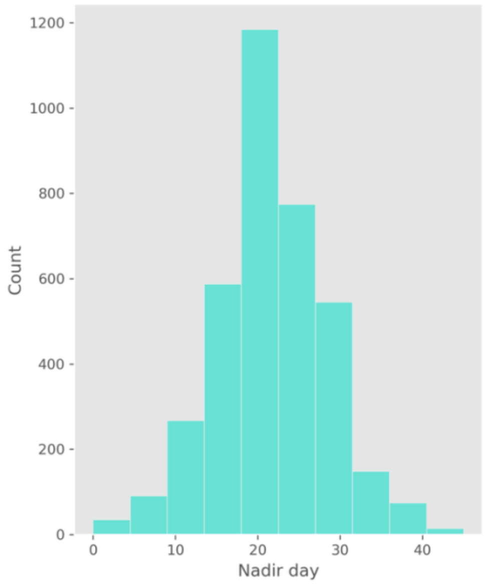
Count
0
200
400
600
800
1000
1200
0
10
20
30
40
Nadir day

**(B)**

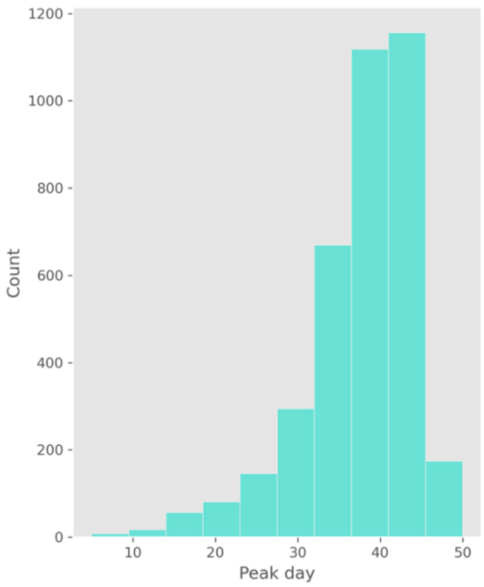
Count
0
200
400
600
800
1000
1200
10
20
30
40
50
Peak day

**(C)**

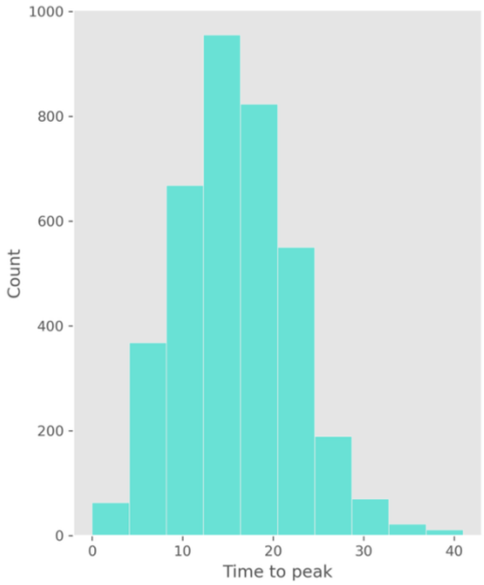
Count
0
200
400
600
800
1000
0
10
20
30
40
Time to peak

**(D)**

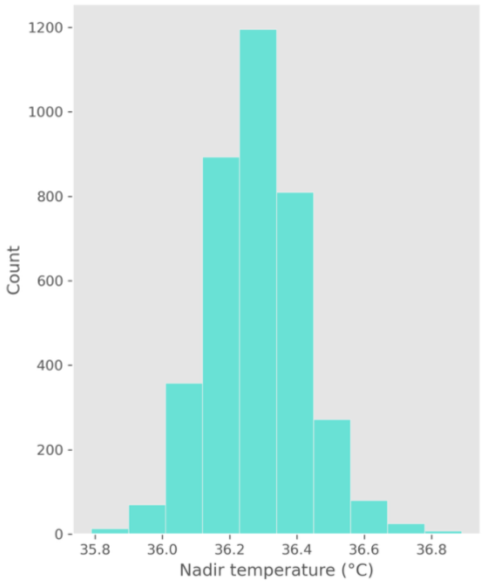


**(E)**

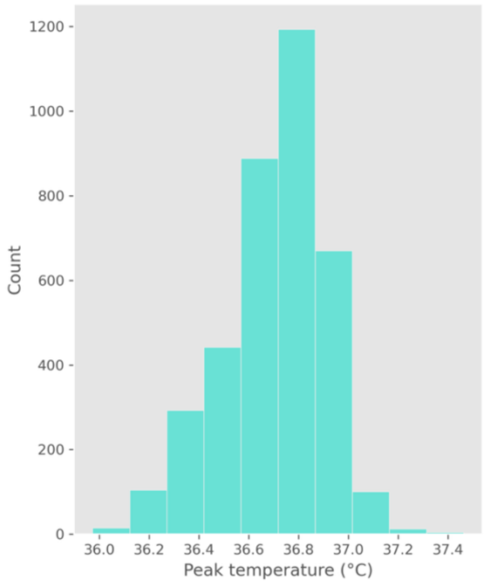


**(F)**

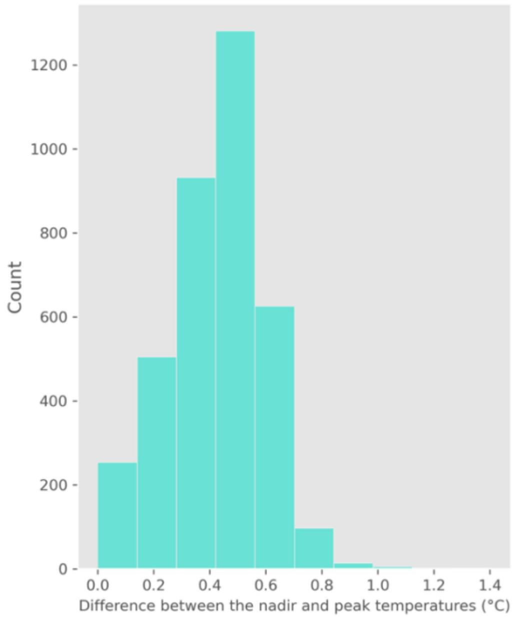


**(G)**

**Figure S7: Distribution of cycle-level physiological and engagement features.** (A) Number of cycles (user engagement): Histogram of the total number of cycles per user, showing the frequency of long-term versus short-term device usage. (B) Nadir day: Distribution of the timing of the lowest temperature point within the cycle. (C) Peak day: Distribution of the timing of the highest temperature point within the cycle. (D) Time to peak (thermal shift timing): Distribution of the difference between nadir and peak days. (E) Nadir temperature: Distribution of the lowest recorded temperature values. (F) Peak temperature: Distribution of the highest recorded temperature values. (G) Difference between the nadir and peak temperatures (post-ovulatory thermal shift): Histogram of the temperature difference between nadir and peak values, illustrating the characteristic magnitude of the post-ovulatory increase. Section I in Multimedia Appendix 1 discusses these results.

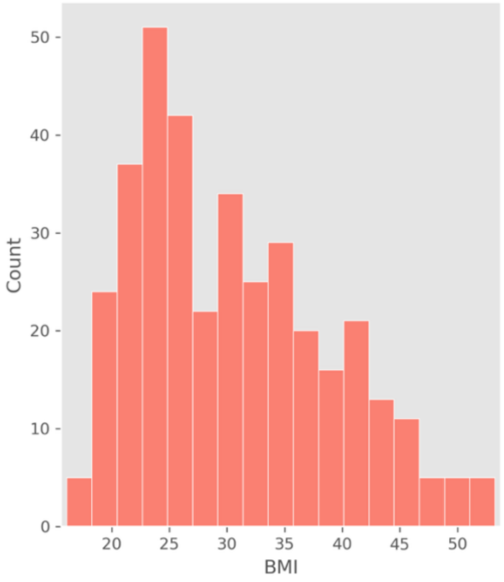
Count
0
10
20
30
40
50
20
25
30
35
40
45
50
BMI

**(A)***

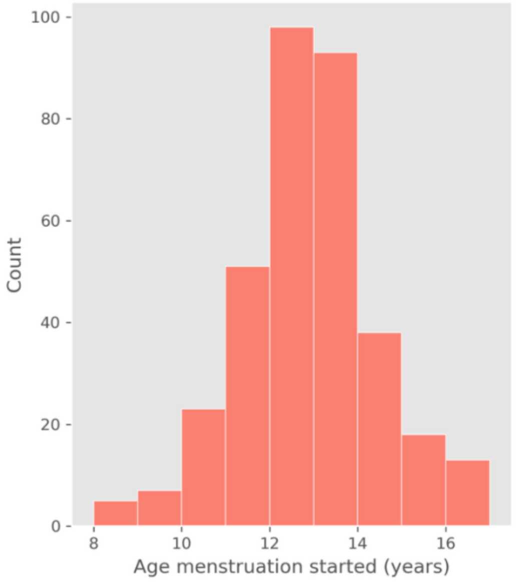
Count
0
20
40
60
80
100
8
10
12
14
16
Age menstruation started (years)

**(B)***

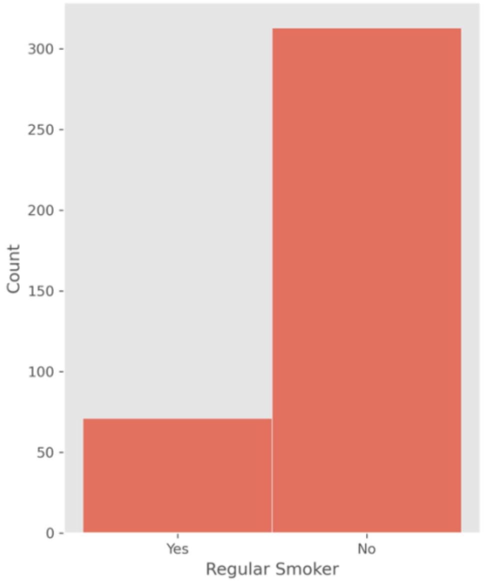
Count
0
50
100
150
200
250
300
Yes
No
Regular Smoker

**(C)**

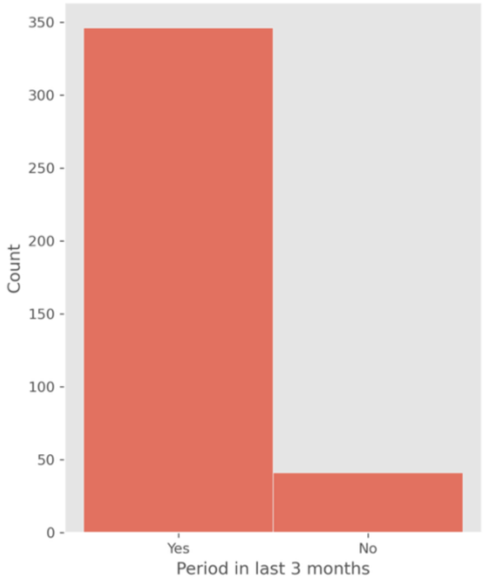
Count
0
50
100
150
200
250
300
350
Yes
No
Period in last 3 months

**(D)**

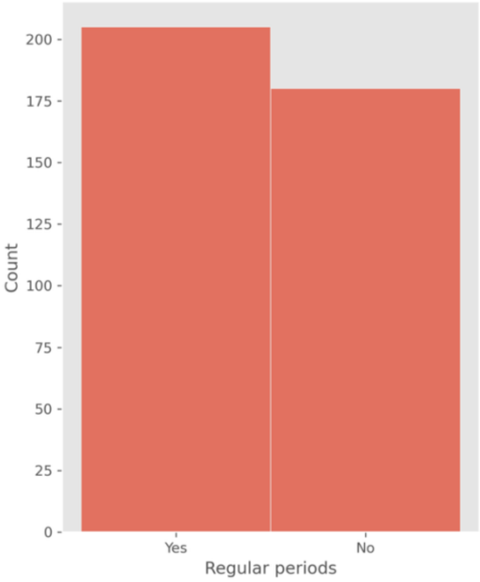


**(E)**

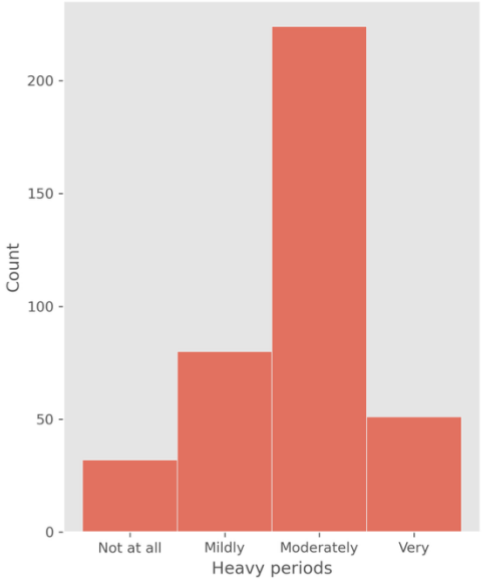


**(F)**

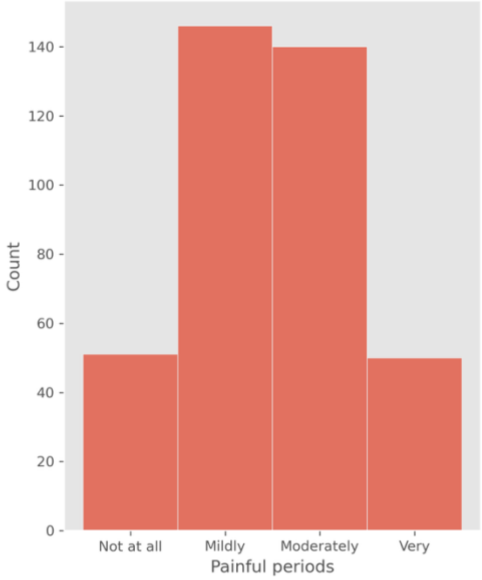


**(G)**

**Figure S8: Distribution of demographic and self-reported questionnaire features.** (A) BMI: Distribution of participants' Body Mass Index. *3 bins containing a total of 5 participants are not shown. (B) Menstrual onset: Distribution of the age at which menstruation started (years). *4 bins containing a total of 5 participants are not shown. (C) Smoking status: Reported frequency of smoking habits. (D) Recent menstrual history: Reported frequency of menstrual periods occurring within the last three months. (E) Cycle regularity: Self-reported regularity of menstrual cycles (regular vs. irregular). (F) Heavy period frequency: Self-reported frequency of heavy menstrual bleeding. (G) Menstrual pain intensity: Self-reported intensity of pain during menstrual periods. Section I in Multimedia Appendix 1 discusses these results. Asterisks (*) indicate categories where counts fewer than five have been suppressed.

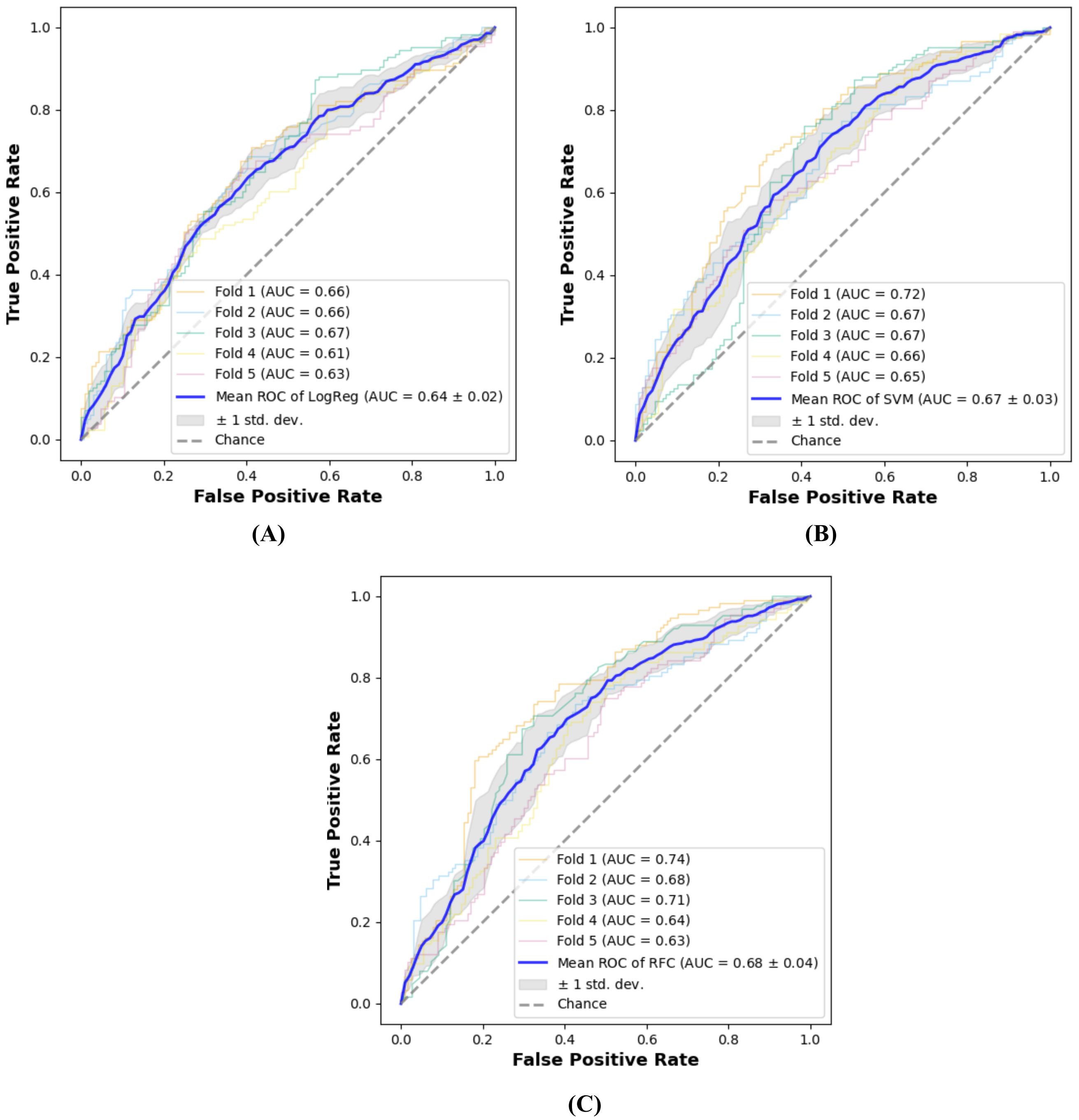


**Figure S9: Cross-validation performance of cycle-level PCOS classification models with variability across folds.** (A) Logistic Regression (LR). (B) Support Vector Machine (SVM). (C) Random Forest Classifier (RF). In each panel, the shaded area represents the standard deviation across the five cross-validation splits, with the bold line representing the mean AUC-ROC score.

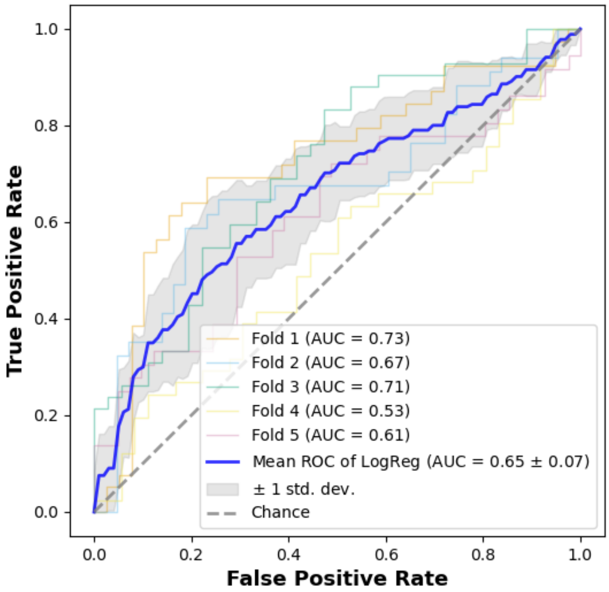


**(A)**

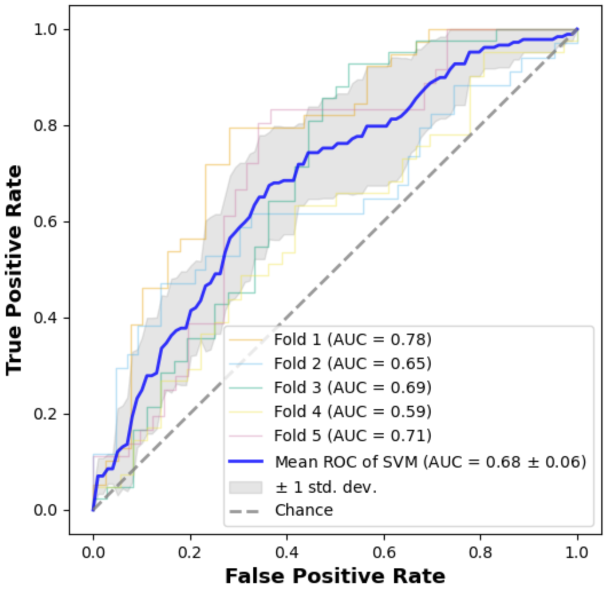


**(B)**

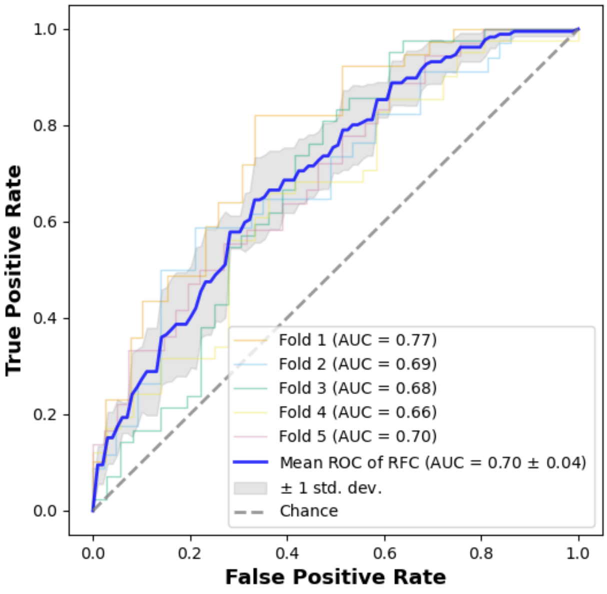


**(C)**

**Figure S10: Cross-validation performance of user-level PCOS classification models with variability across folds.** (A) LR (B) SVM (C) RF. In each panel, the shaded area represents the standard deviation across the five cross-validation splits, with the bold line representing the mean AUC-ROC score.

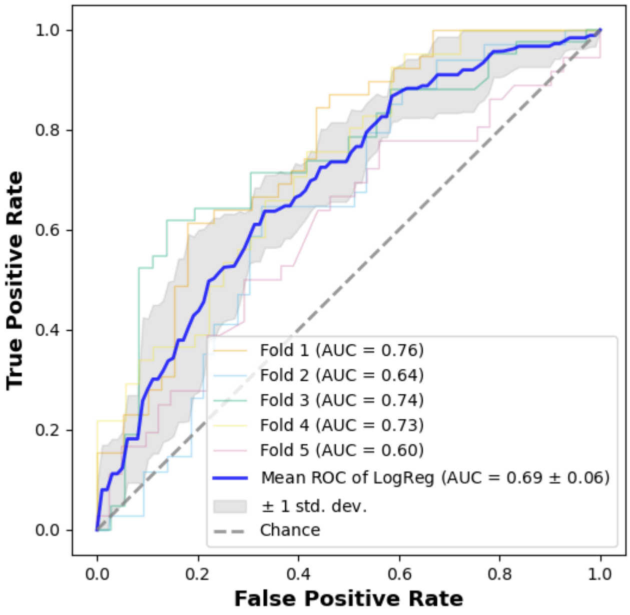


**(A)**

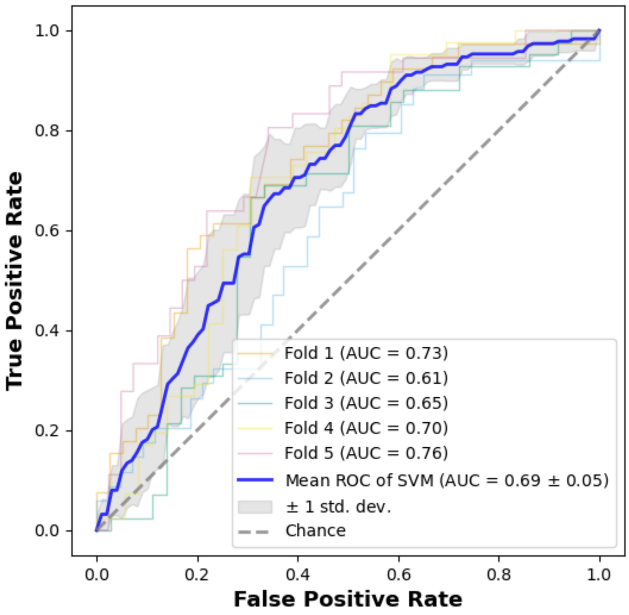


**(B)**

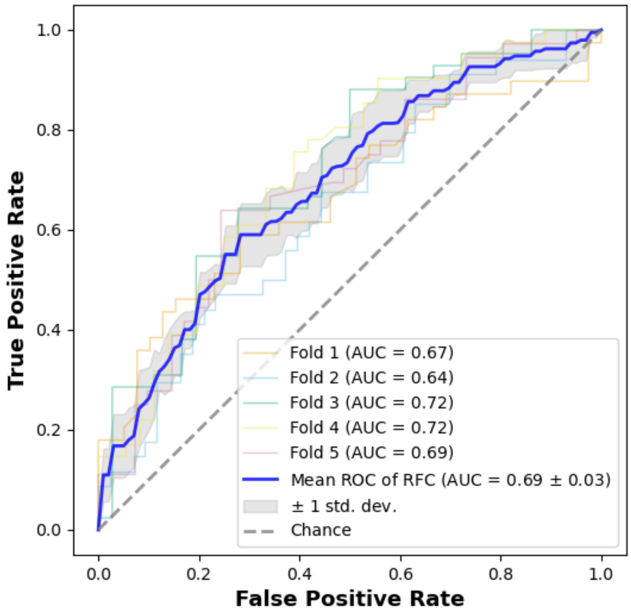


**(C)**

**Figure S11: Cross-validation performance of questionnaire-level PCOS classification models with variability across folds.** (A) LR (B) SVM (C) RF. In each panel, the shaded area represents the standard deviation across the five cross-validation splits, with the bold line representing the mean AUC-ROC score.

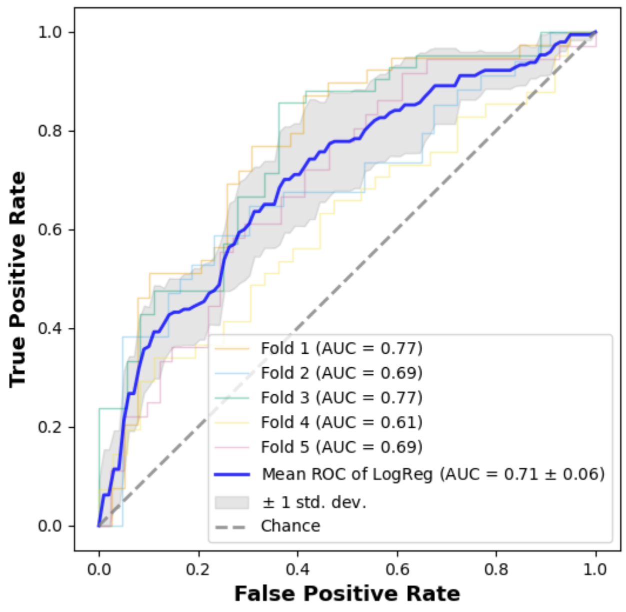


**(A)**

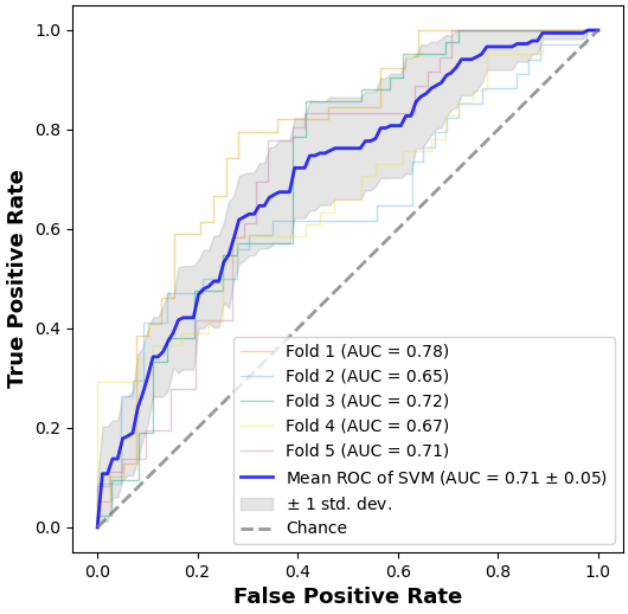


**(B)**

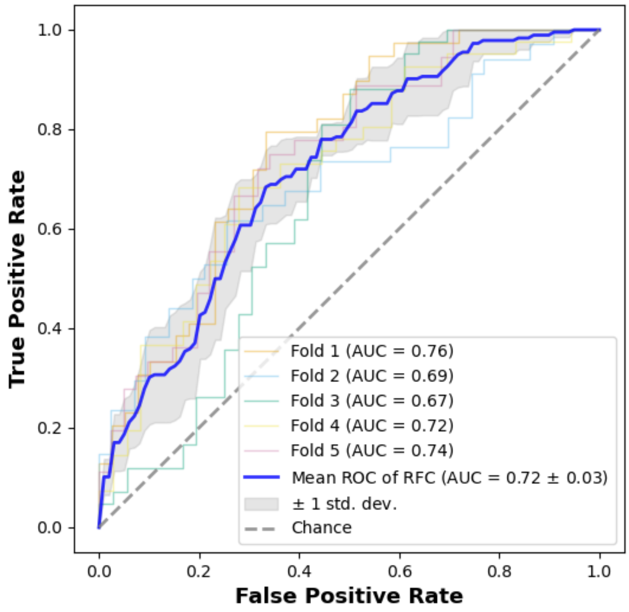


**(C)**

**Figure S12: Cross-validation performance of the combined user-level and questionnaire PCOS classification models with variability across folds.** (A) LR (B) SVM (C) RF. In each panel, the shaded area represents the standard deviation across the five cross-validation splits, with the bold line representing the mean AUC-ROC score**.**

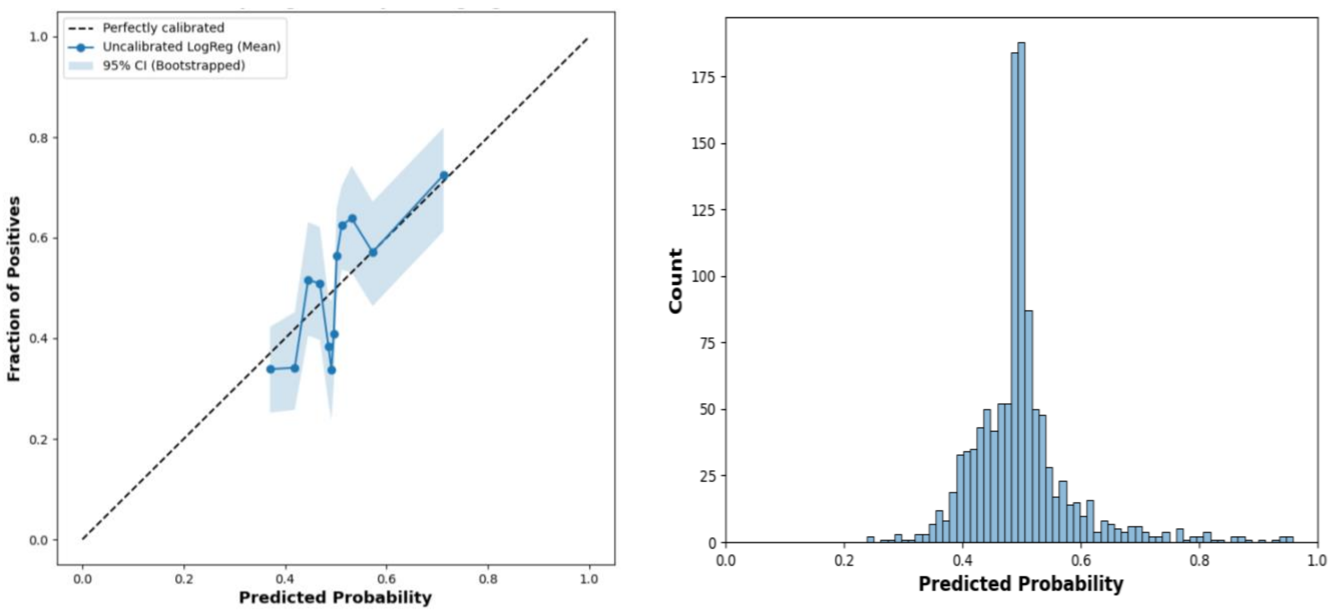


**(A)** **(B)**

**Figure S13: Calibration and reliability of the cycle-level LR model.** (A) Reliability curve indicating calibration performance across the probability range. (B) Probability distribution histogram.

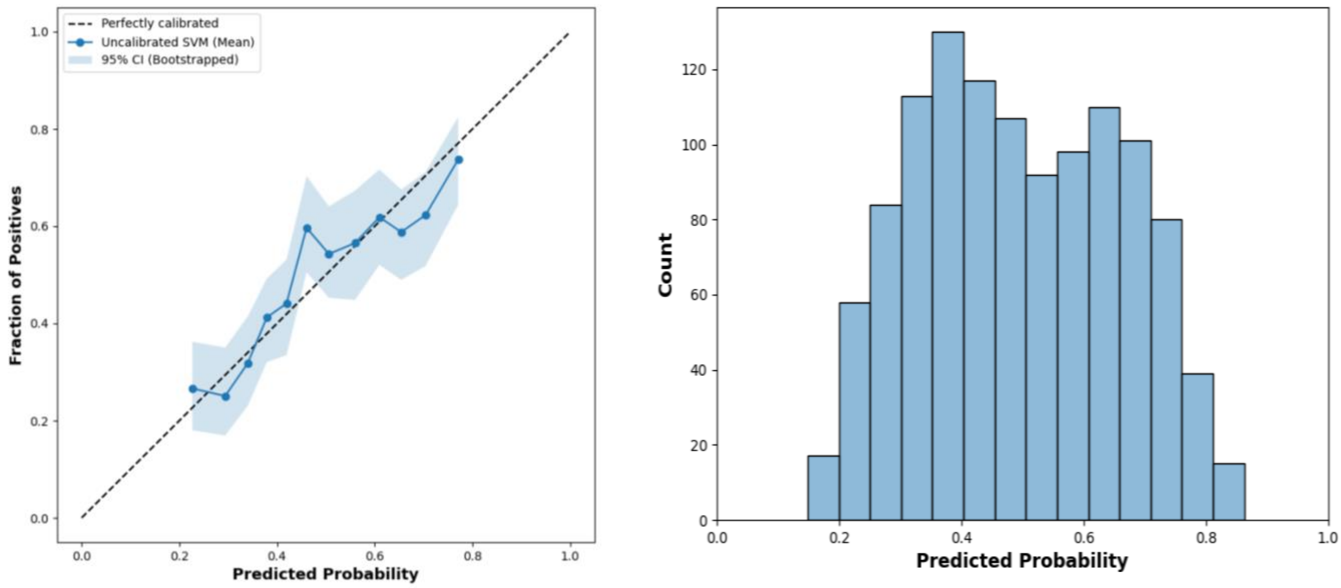


**(A)** **(B)**

**Figure S14: Calibration and reliability of the cycle-level SVM model.** (A) Reliability curve indicating calibration performance across the probability range. (B) Probability distribution histogram.

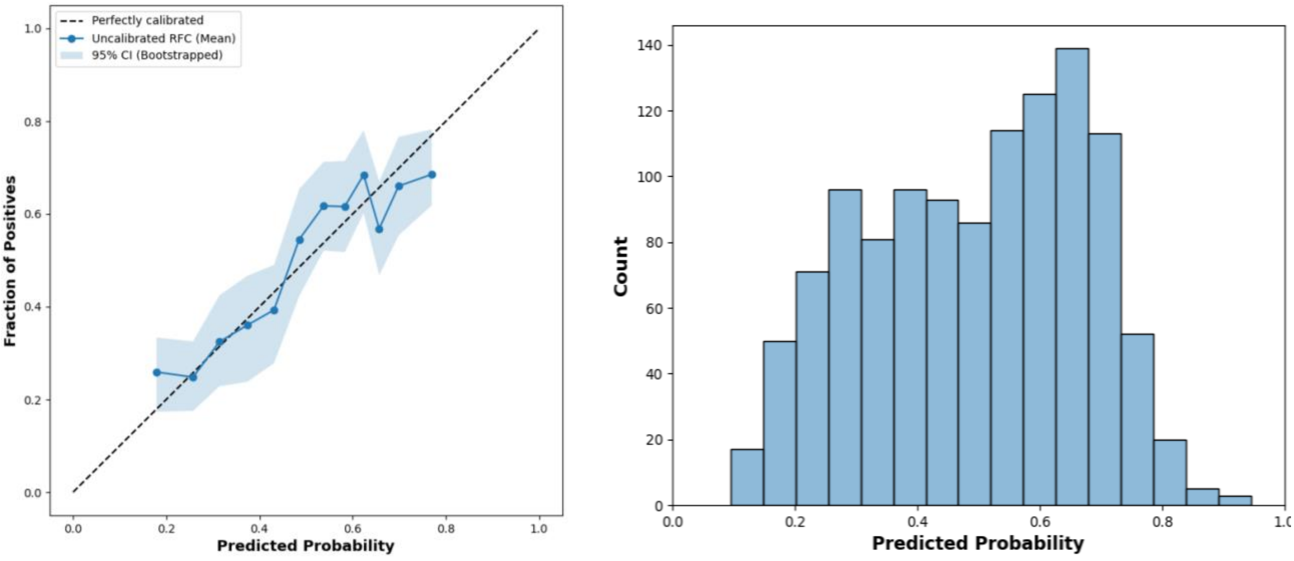


**(A)** **(B)**

**Figure S15: Calibration and reliability of the cycle-level RF model.** (A) Reliability curve indicating calibration performance across the probability range. (B) Probability distribution histogram.

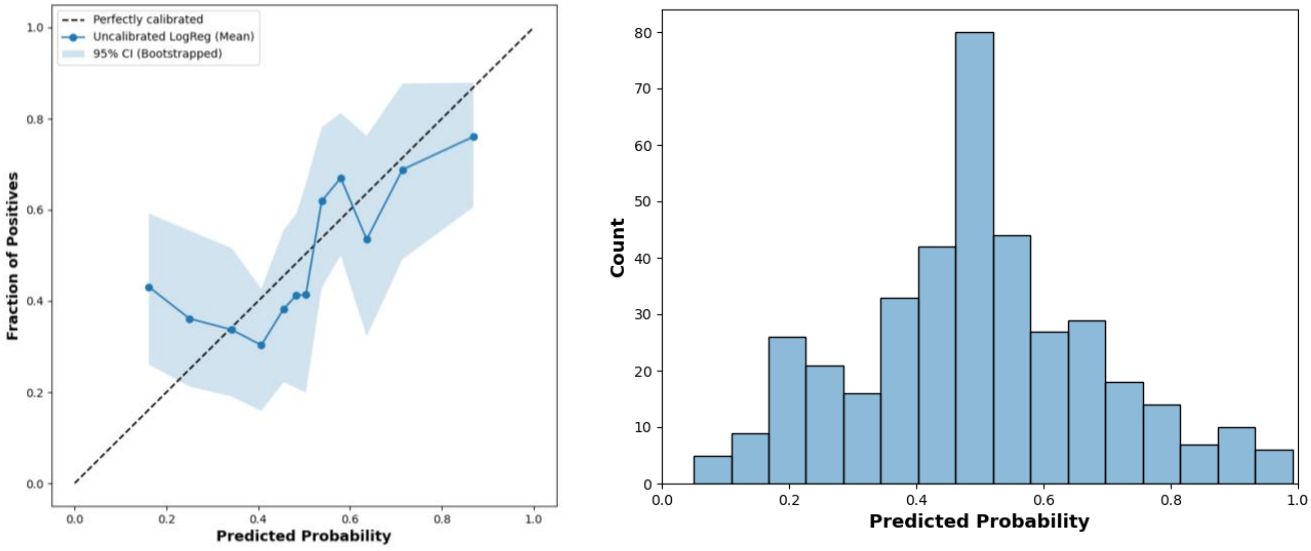


**(A)** **(B)**

**Figure S16: Calibration and reliability of the user-level LR model.** (A) Reliability curve indicating calibration performance across the probability range. (B) Probability distribution histogram.

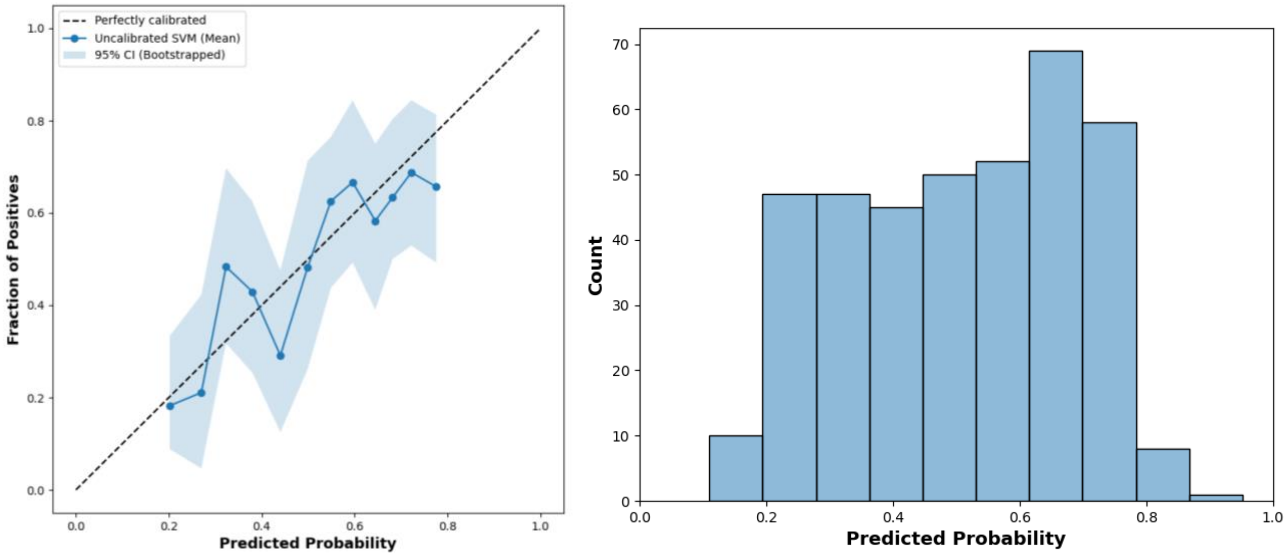


(A) (B)

**Figure S17: Calibration and reliability of the user-level SVM model.** (A) Reliability curve indicating calibration performance across the probability range. (B) Probability distribution histogram.

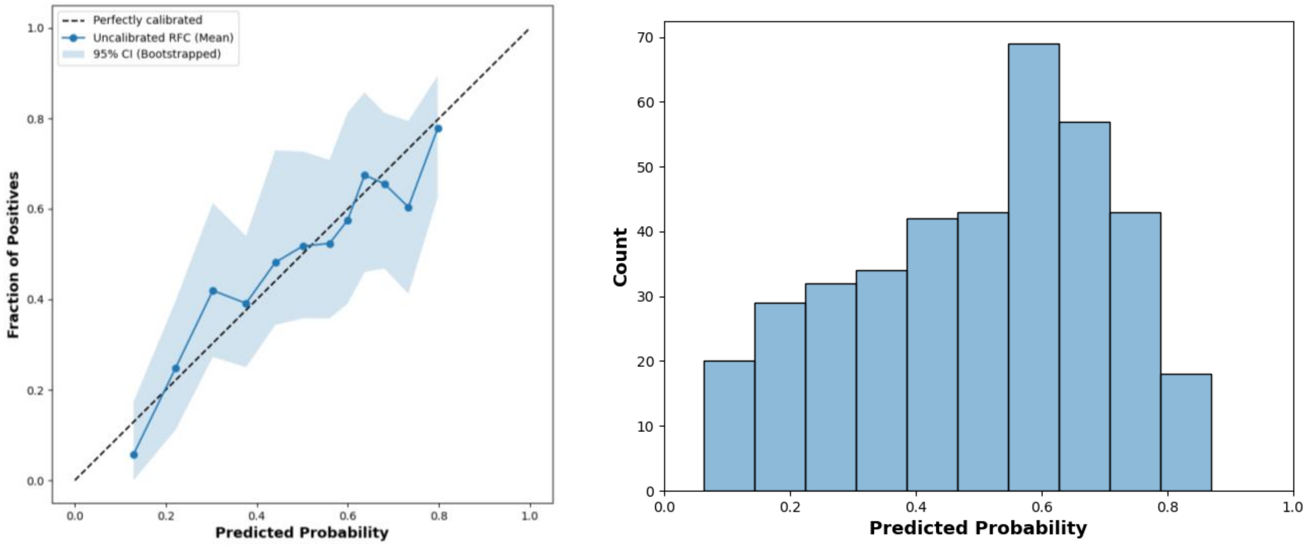


(A) (B)

**Figure S18: Calibration and reliability of the user-level RF model.** (A) Reliability curve indicating calibration performance across the probability range. (B) Probability distribution histogram.

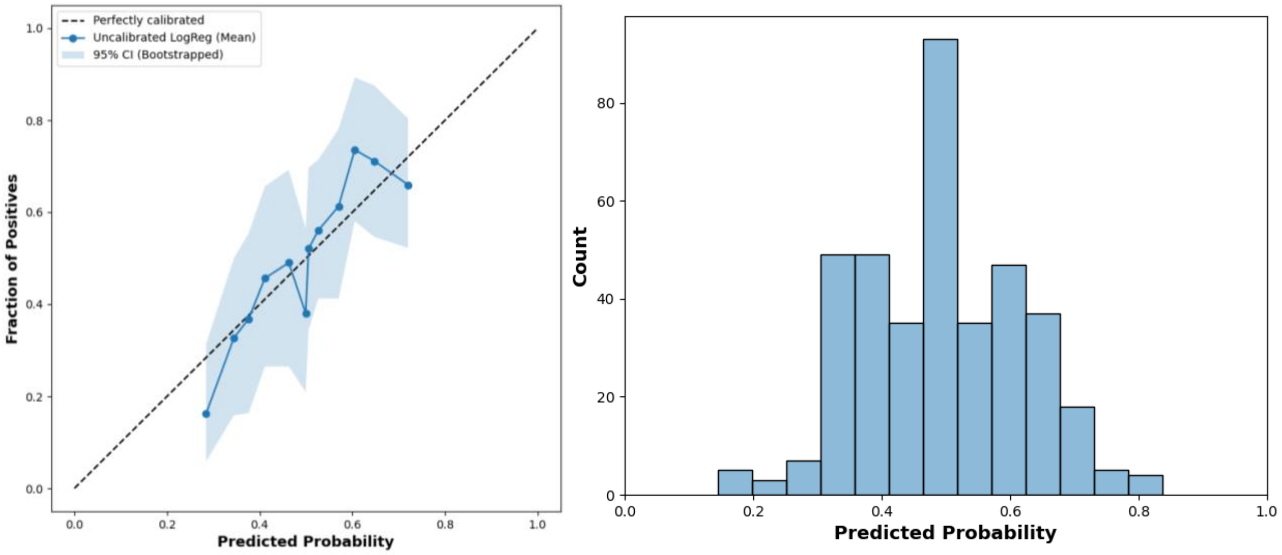


(A) (B)

**Figure S19: Calibration and reliability of the questionnaire-based LR model.** (A) Reliability curve indicating calibration performance across the probability range. (B) Probability distribution histogram.

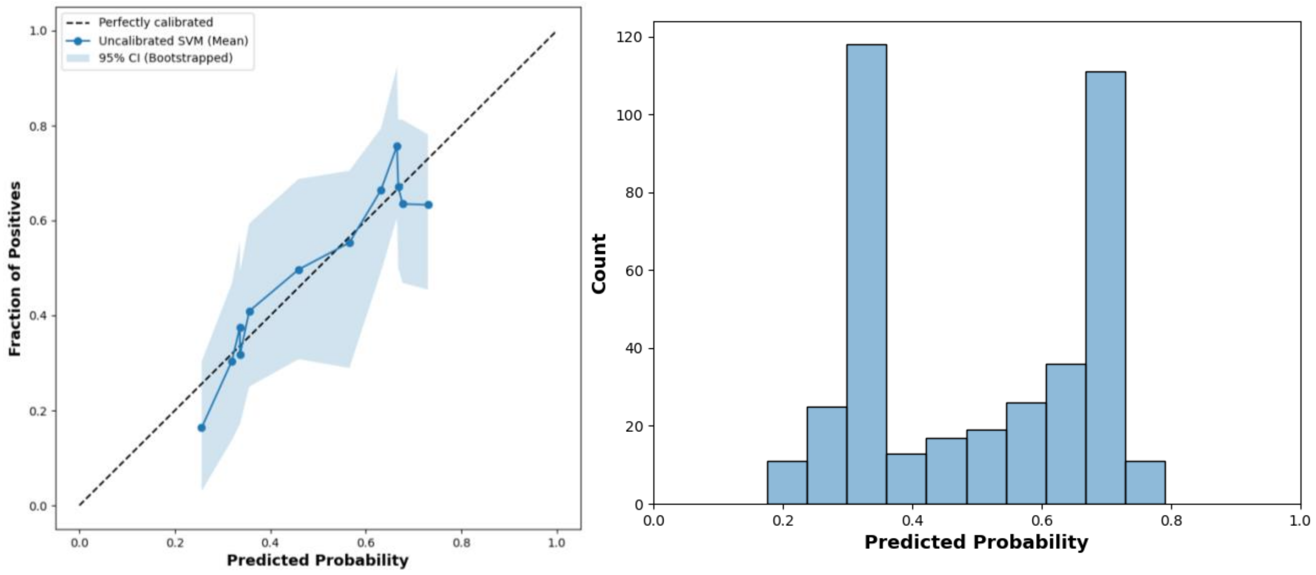


(A) (B)

**Figure S20: Calibration and reliability of the questionnaire-based SVM model.** (A) Reliability curve indicating calibration performance across the probability range. (B) Probability distribution histogram.

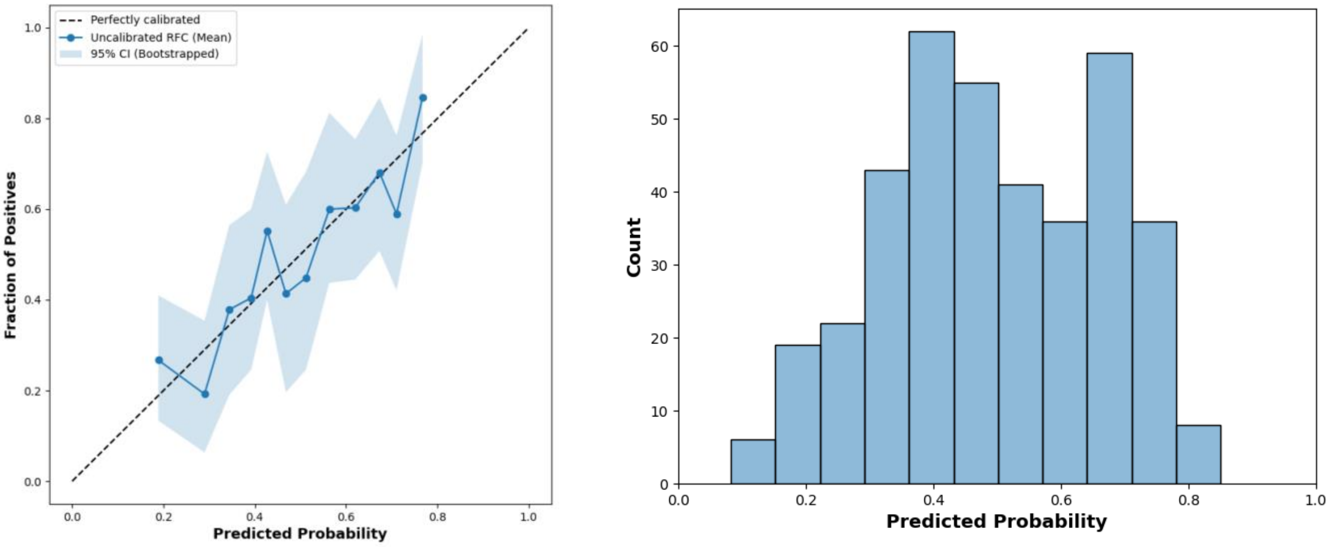


**(A)** **(B)**

**Figure S21: Calibration and reliability of the questionnaire-based RF model.** (A) Reliability curve indicating calibration performance across the probability range. (B) Probability distribution histogram.

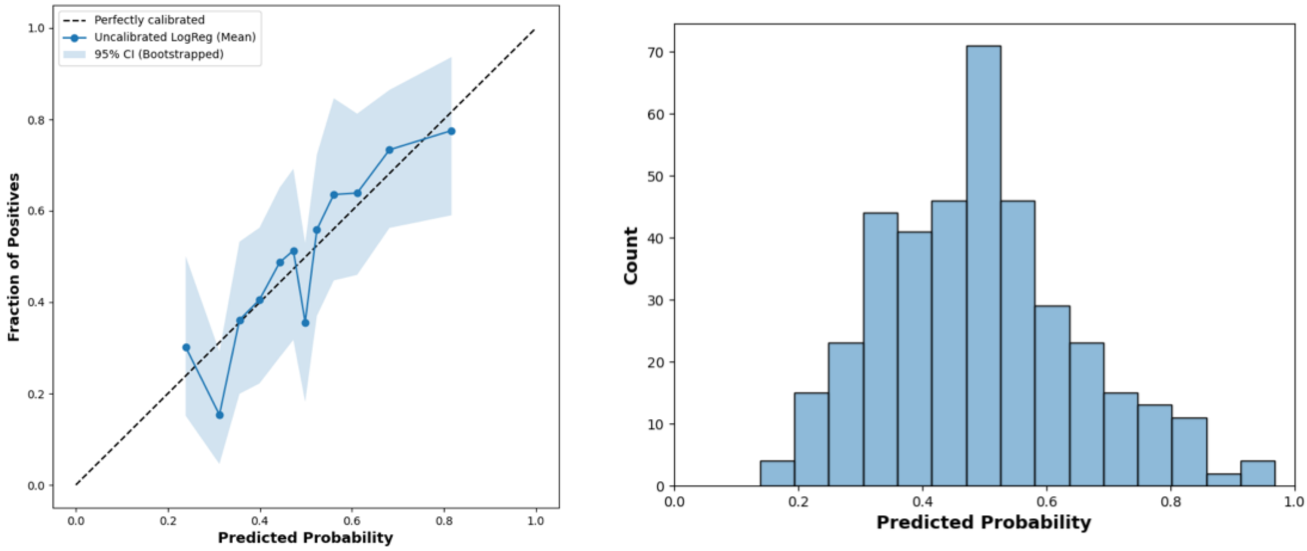


**(A)** **(B)**

**Figure S22: Calibration and reliability of the combined user-level and questionnaire-based LR model.** (A) Reliability curve indicating calibration performance across the probability range. (B) Probability distribution histogram.

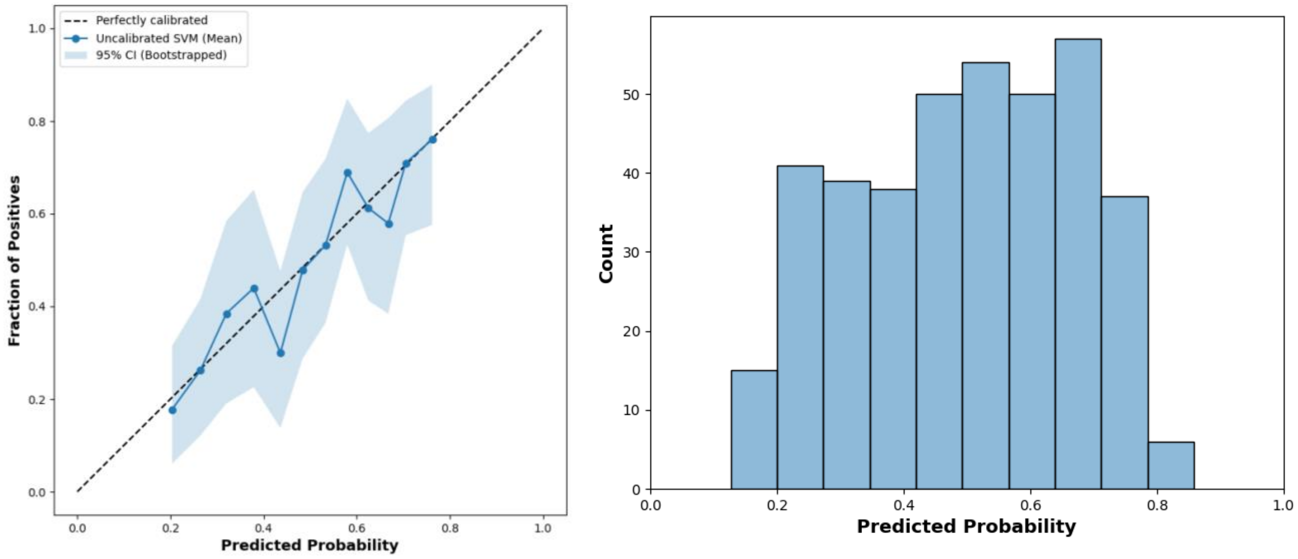


(A) (B)

**Figure S23: Calibration and reliability of the combined user-level and questionnaire-based SVM model.** (A) Reliability curve indicating calibration performance across the probability range. (B) Probability distribution histogram.

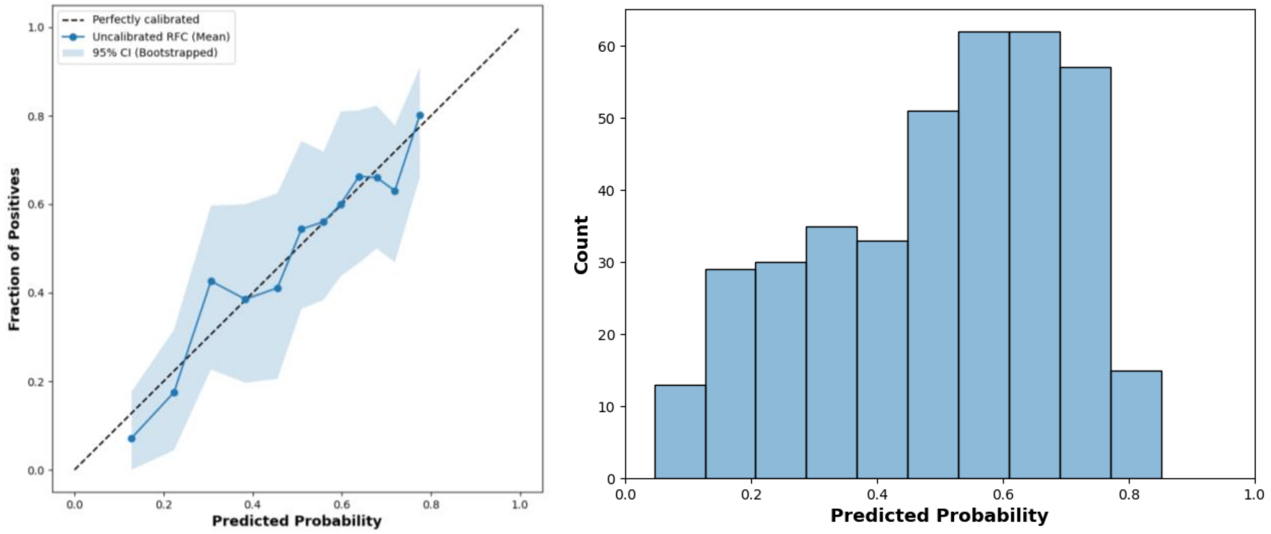


(A) (B)

**Figure S24: Calibration and reliability of the combined user-level and questionnaire-based RF model.** (A) Reliability curve indicating calibration performance across the probability range. (B) Probability distribution histogram.

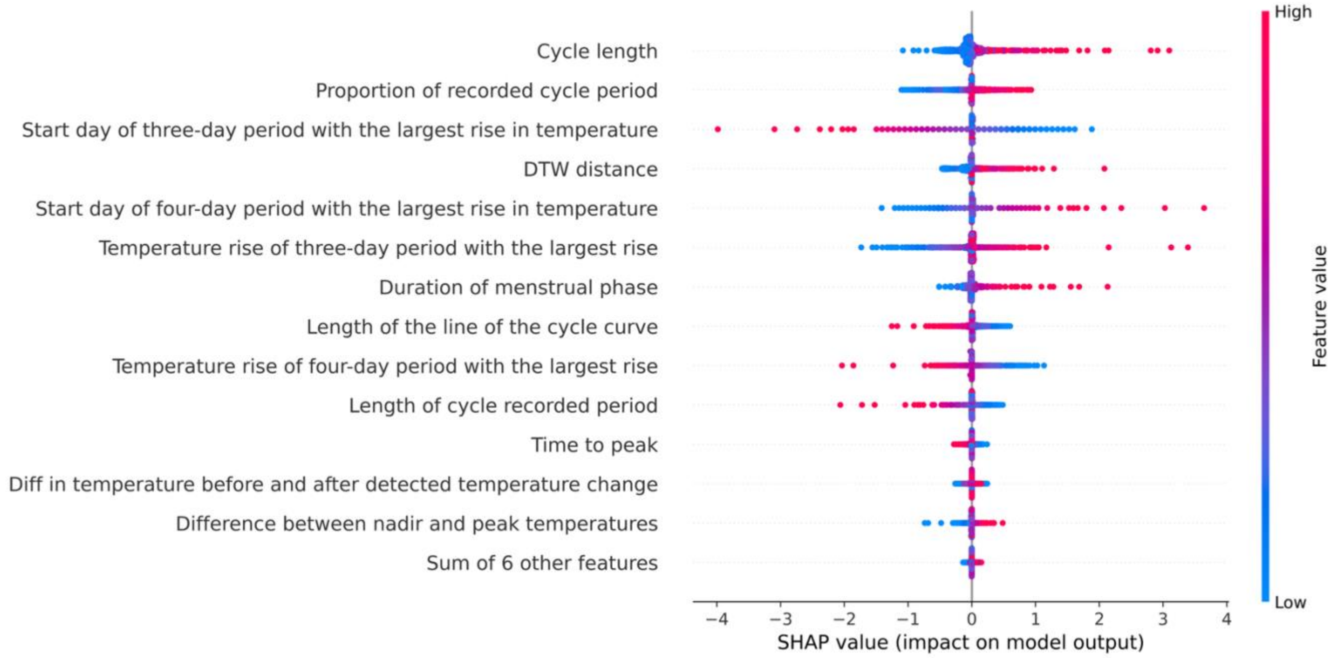


**Figure S25: Cycle-level SHAP summary plot for the LR model.** The plot illustrates the distribution of SHAP values across the features. Features are ranked by their global importance, with red and blue dots indicating high and low feature values, respectively.

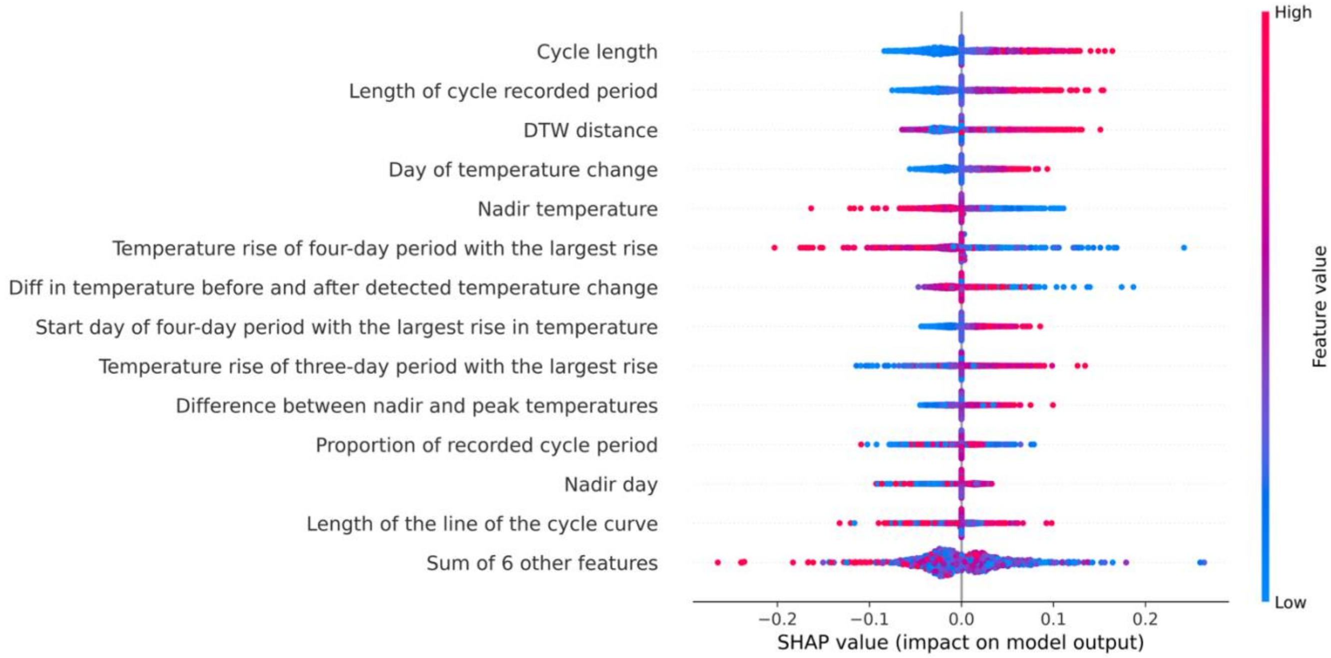


**Figure S26: Cycle-level SHAP summary plot for the SVM model.** The plot displays the impact of individual cycle-level features on the SVM model's output. *Cycle length* and *Length of cycle recorded period* are highlighted as contributing the most to the model's predictions.

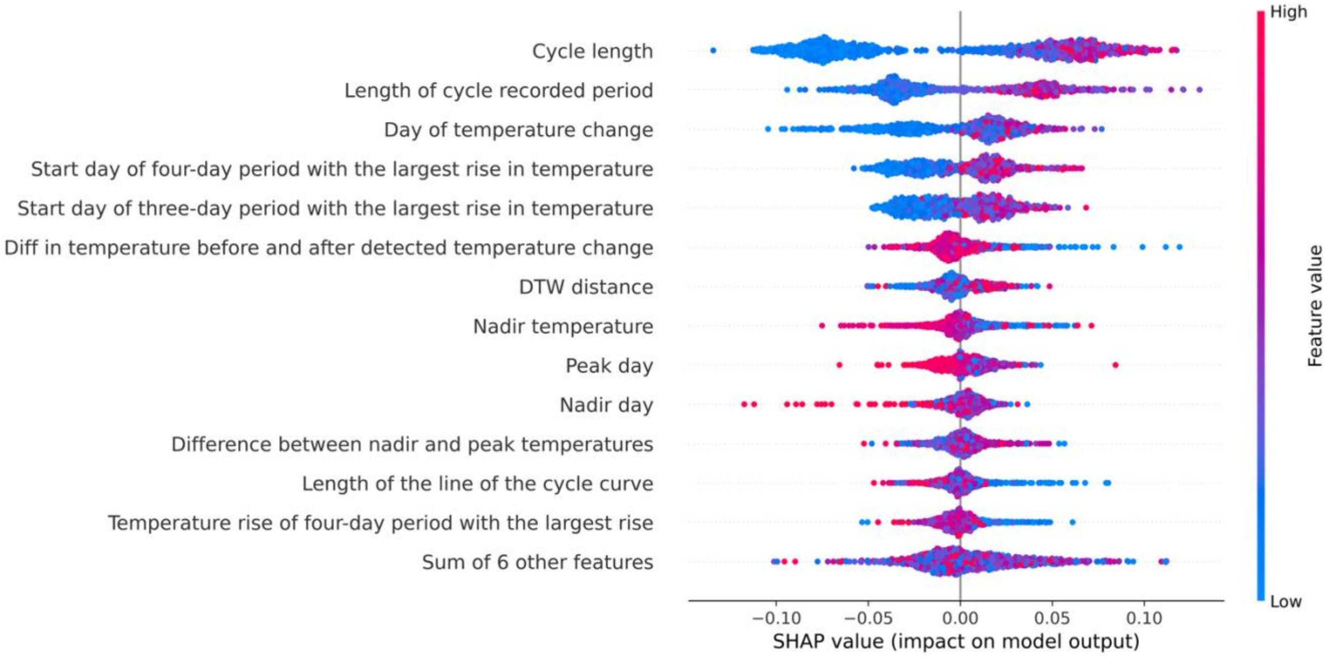


**Figure S27: Cycle-level SHAP summary plot for the RF model.** The plot depicts the feature influence on the ensemble model. *Cycle length* and *Length of the recorded period* are shown as playing a primary role in the decision-making process.

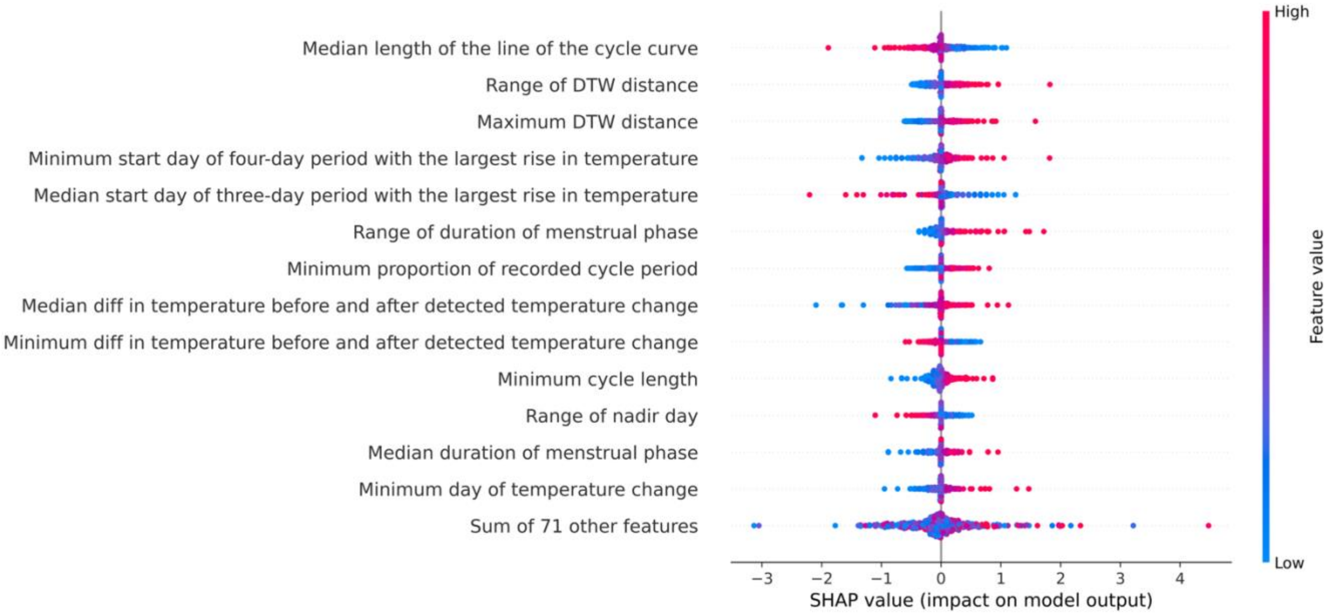


**Figure S28: User-level SHAP summary plot for the Logistic Regression model.** Features derived from *DTW distance* demonstrated high importance

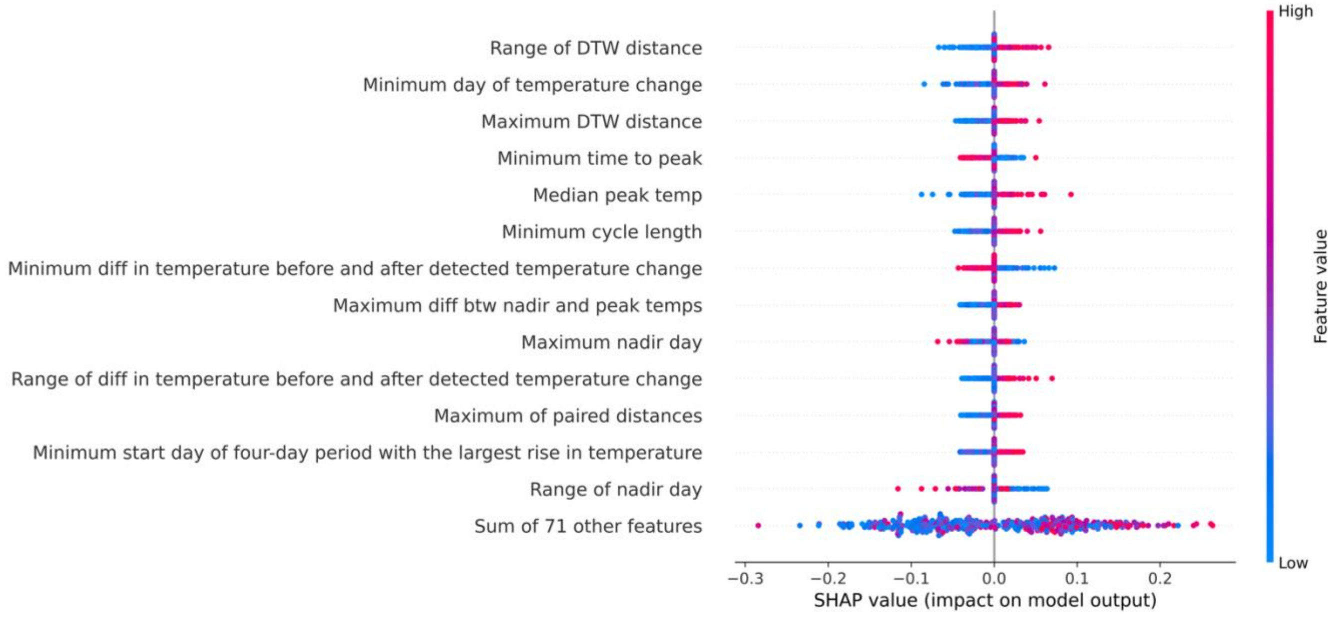


**Figure S29: User-level SHAP summary plot for the Support Vector Machine model.** Features derived from *DTW distance* demonstrated high importance

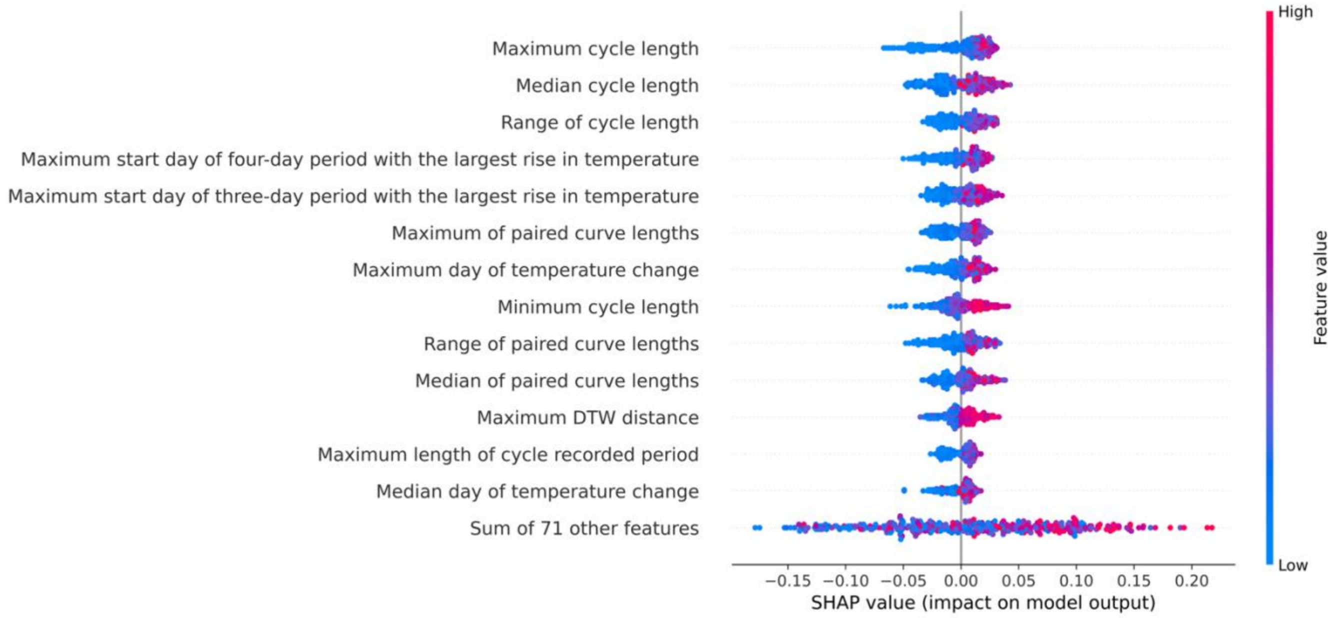


**Figure S30: User-level SHAP summary plot for the Random Forest model.** Features based on cycle length were the primary contributors

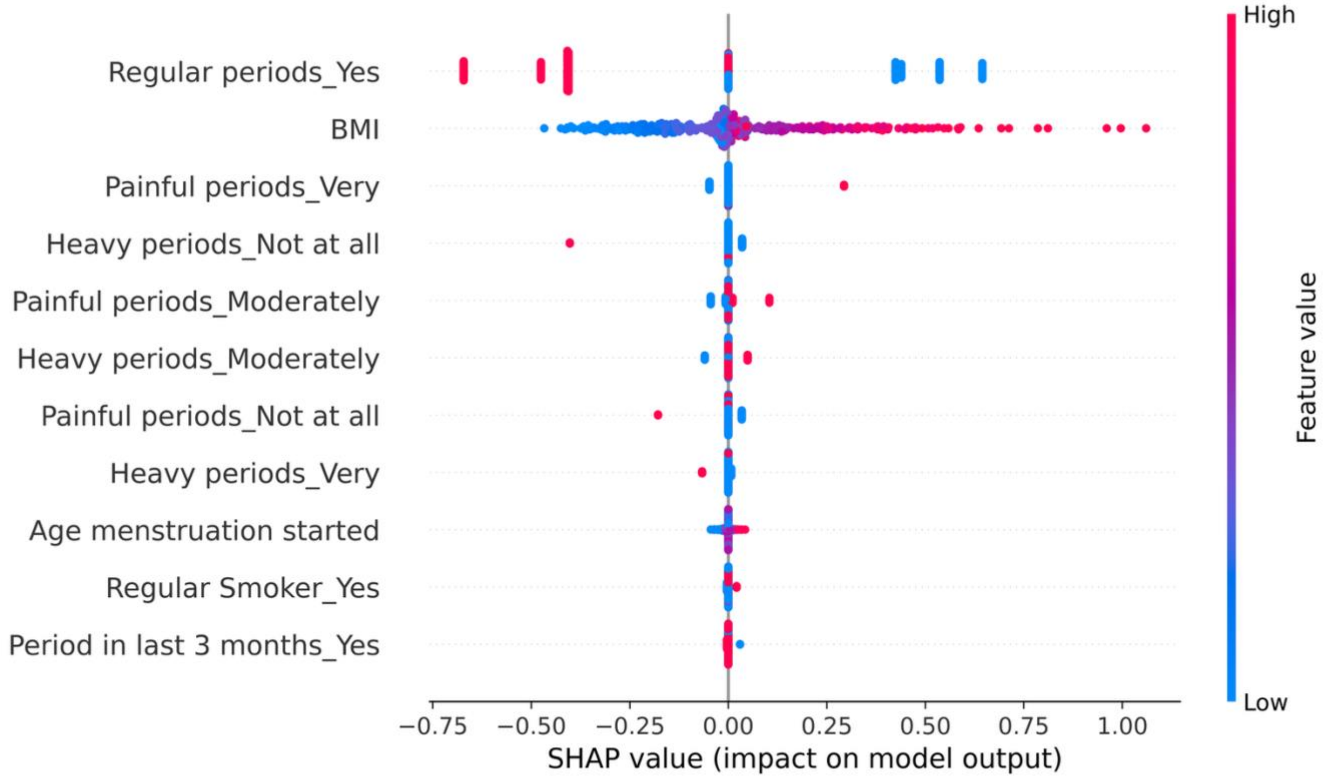


**Figure S31: Questionnaire-based SHAP summary plot for the Logistic Regression model.** Menstrual period regularity and BMI were highly influential features

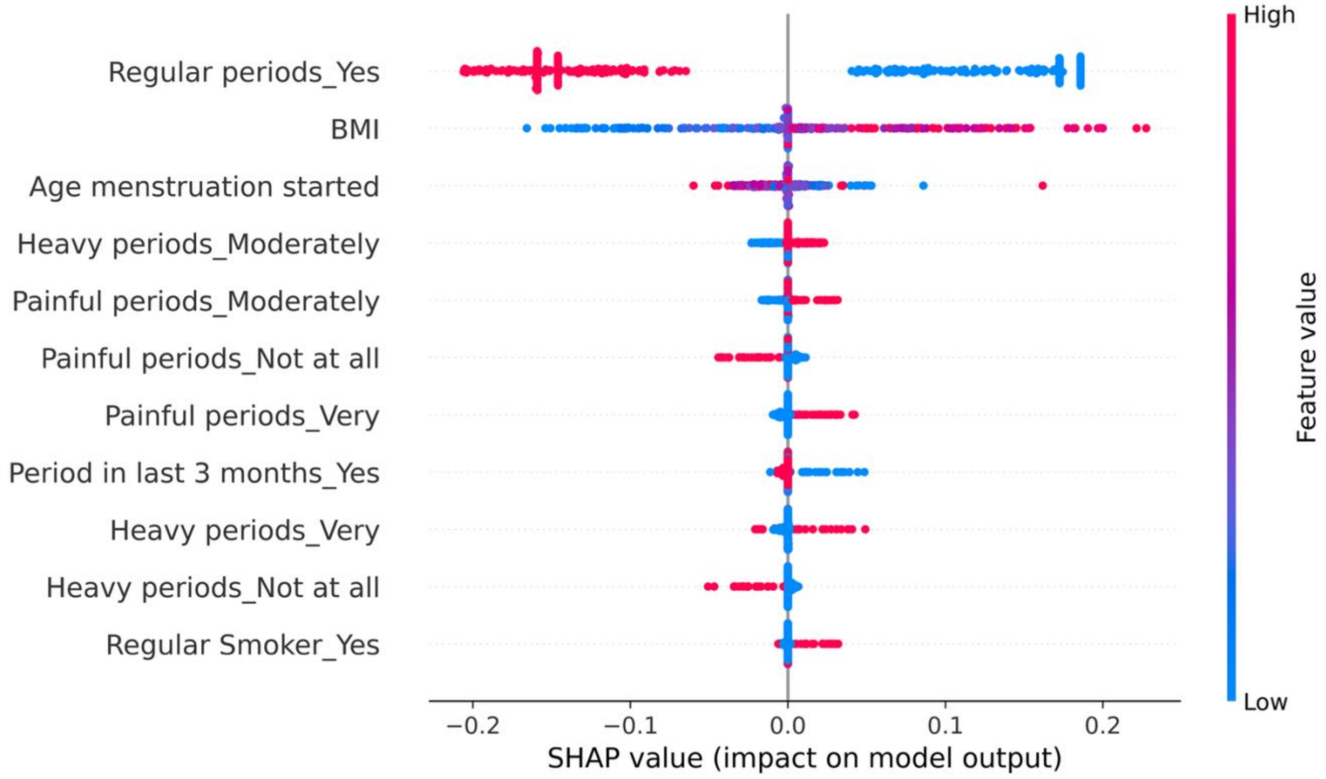


**Figure S32: Questionnaire-based SHAP summary plot for the Support Vector Machine model.** Menstrual period regularity and BMI were highly influential features

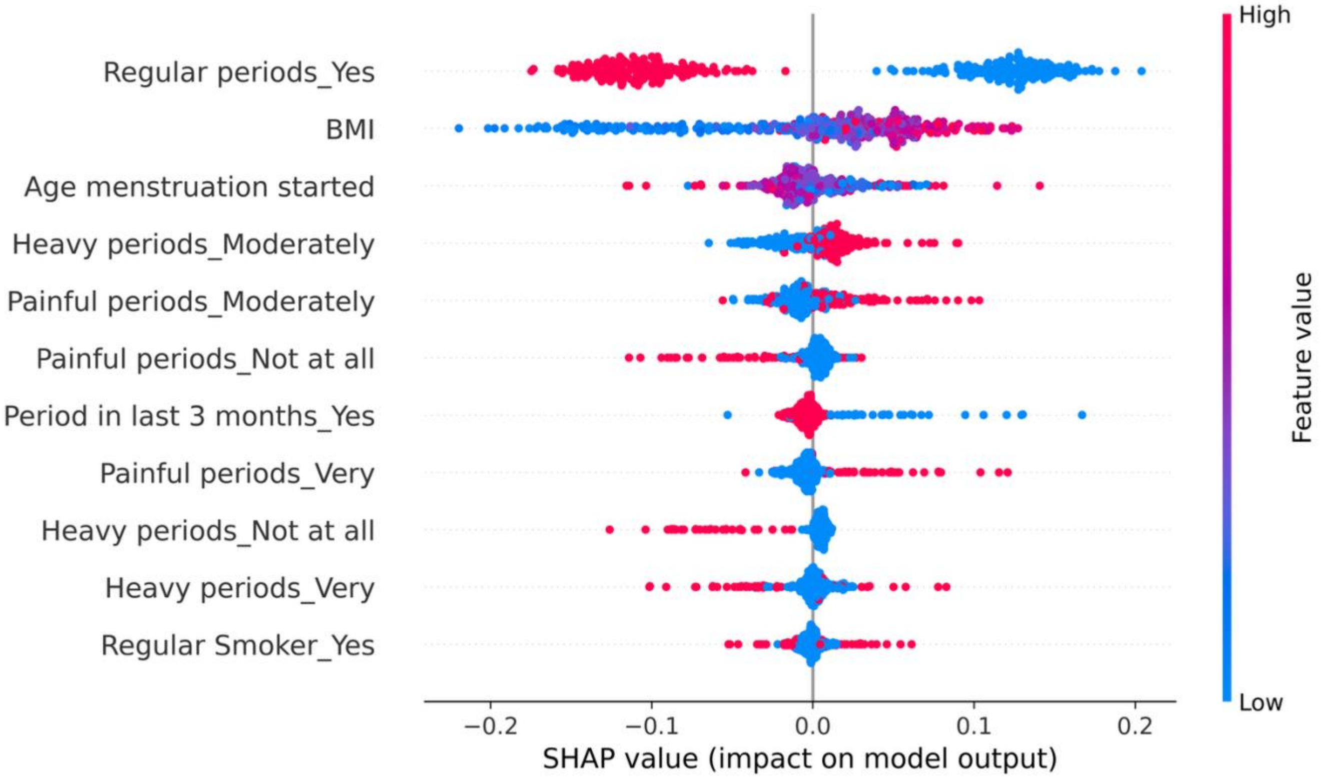


**Figure S33: Questionnaire-based SHAP summary plot for the Random Forest model.** Menstrual period regularity and BMI were highly influential features

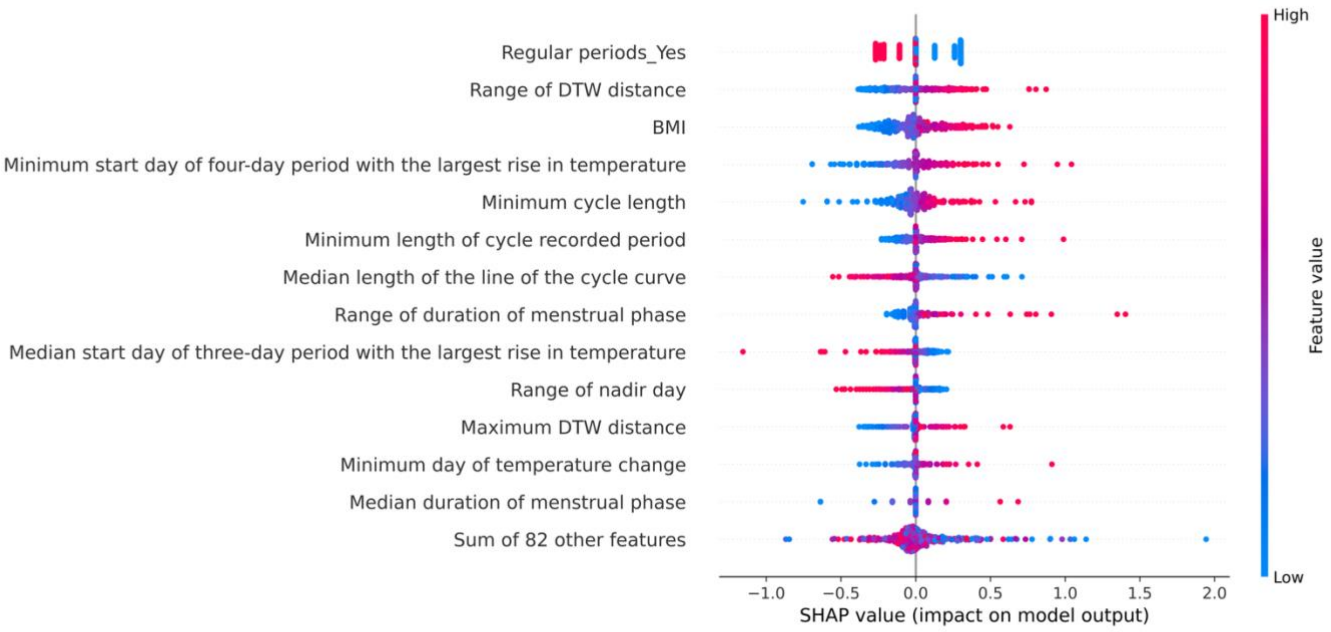


**Figure S34: Combined user-level and questionnaire SHAP summary plot for the Logistic Regression model.** The combined models synthesise information from both lifestyle and physiological data sources to inform classification.

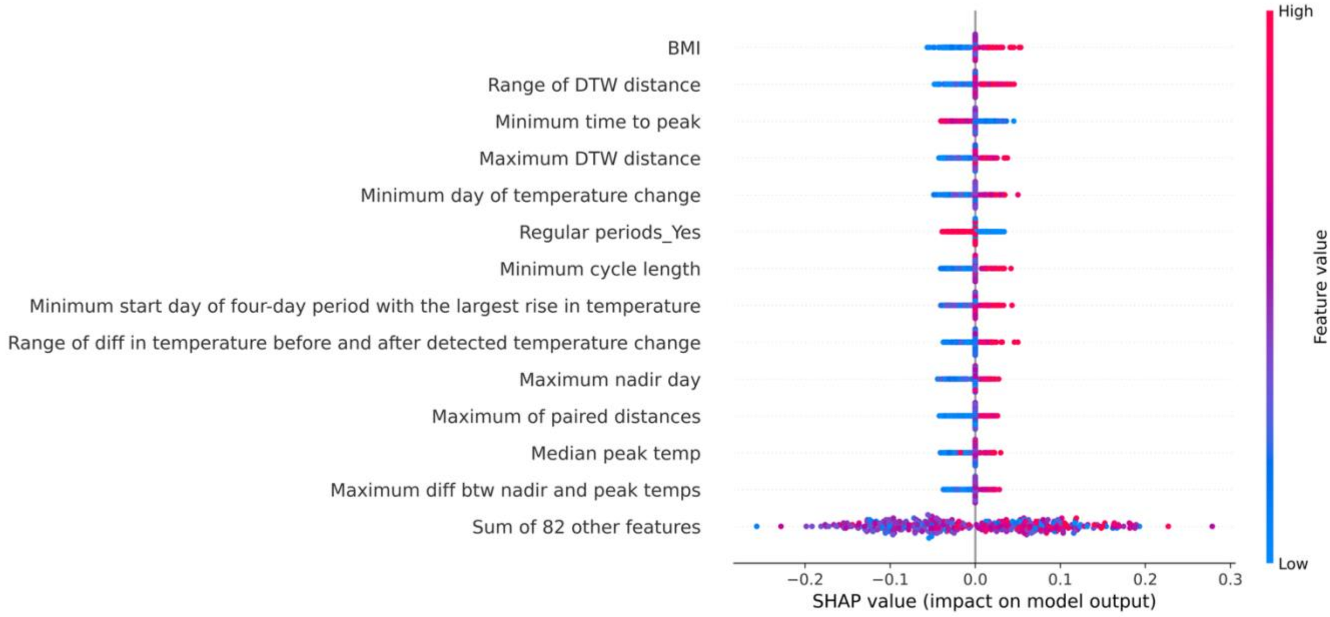


**Figure S35: Combined user-level and questionnaire SHAP summary plot for the Support Vector Machine model.** The combined models synthesise information from both lifestyle and physiological data sources to inform classification.

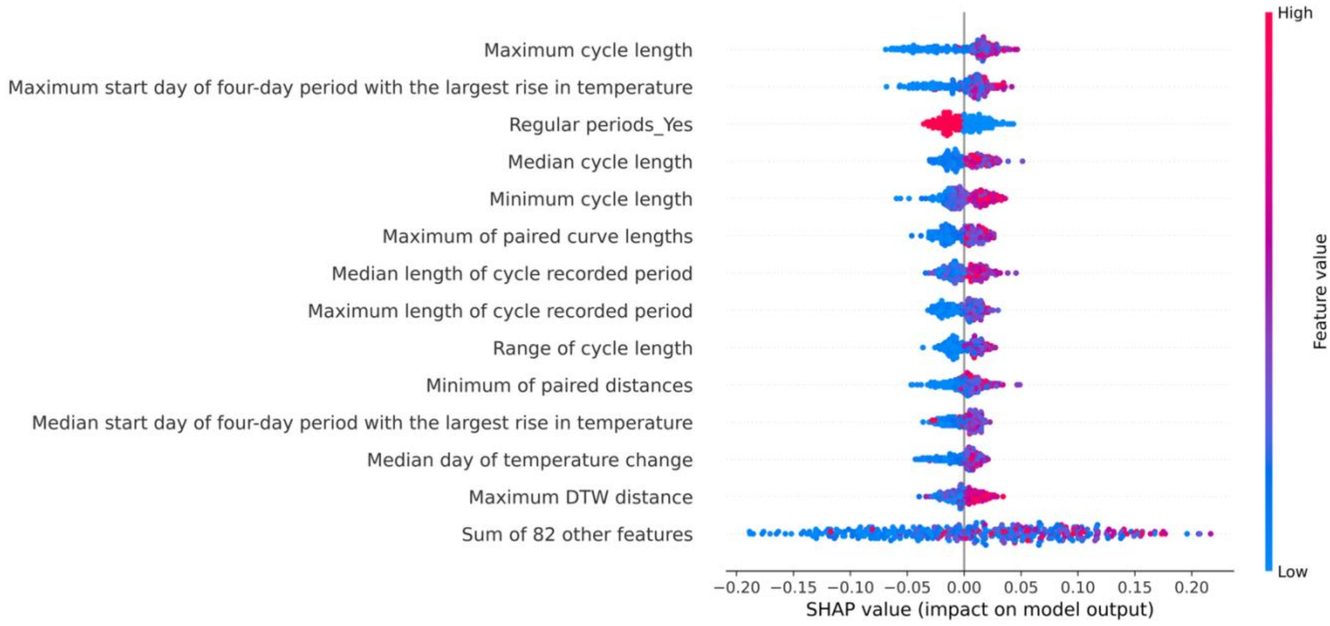


**Figure S36: Combined user-level and questionnaire SHAP summary plot for the Random Forest model.** The combined models synthesise information from both lifestyle and physiological data sources to inform classification.
