## Supplementary material for "A digital health approach for identifying polyendocrine metabolic ovarian syndrome using machine learning and body temperature": Multimedia appendix 3

Awoniran et al.

### MULTIMEDIA APPENDIX 3 – TABLES

**Table S1: PCOS Outcome Groupings of the Analytical Sample**

| Classification | Subgroup Description | n | Total (N) |
|---|---|---|---|
| **PCOS Cases** | Self-Reported Physician Diagnoses | 185 | **192** |
| | Medication-Defined (only)[a] | 7 | |
| **Controls** | Never visited doctor for infertility | 78 | **195** |
| | Visited doctor for fertility and diagnosis given but not for PCOS [b] | 61 | |
| | Visited doctor for fertility and no diagnosis given | 49 | |
| | Visited doctor for fertility but did not indicate diagnosis | 7 | |
| **Total Population** | | | **387** |

[a] Participants who did not report a doctor's visit but confirmed taking medication specifically for PCOS
[b] Includes *Other ovarian problem* (n=6), *Endometriosis* (n=10), *Problem with your cervix* (1), *Partner/Sperm issues* (n=12), *Thyroid disorders* (n=14), *Pelvic inflammatory disease* (n=1) and *others* (n=40). The list is not mutually exclusive, and some individuals have more than one response.

n - Subsample or the size of a specific.

N - The total size of the analytical sample.

**Table S2: The complete list of the user-level features.** This table details the 84 features derived for each participant. Features are categorized into two primary methodologies: Inter-Cycle Pairing Metrics (Features 1–8), which evaluate internal cycle stability via dynamic time warping (DTW) distance and warping path lengths; and aggregated cycle-level metrics (Features 9–84), which represent the statistical distribution (minimum, maximum, median, and range) of physiological and morphological parameters across the three randomly selected cycles per participant.

| | **The Features** |
|---|---|
| 1 | The minimum of paired distances |
| 2 | The maximum of paired distances |
| 3 | The median of paired distances |
| 4 | The range of paired distances |
| 5 | The minimum of paired curve lengths |
| 6 | The maximum of paired curve lengths |
| 7 | The median of paired curve lengths |
| 8 | The range of paired curve lengths |
| 9 | The minimum duration of menstrual phase |
| 10 | The minimum length of cycle recorded period |
| 11 | The minimum cycle length |
| 12 | The minimum proportion of recorded cycle period |
| 13 | The minimum length of the line of the cycle curve |
| 14 | The minimum start day of three-day period with the largest rise in temperature |
| 15 | The minimum temperature rise of three-day period with the largest rise |
| 16 | The minimum start day of four-day period with the largest rise in temperature |
| 17 | The minimum temperature rise of four-day period with the largest rise |
| 18 | The minimum day of temperature change |
| 19 | The minimum difference in temperature before and after detected temperature change |
| 20 | The minimum DTW distance |
| 21 | The minimum length of optimal warping path |
| 22 | The minimum nadir temperature |
| 23 | The minimum peak temperature |
| 24 | The minimum difference between nadir and peak temperatures |
| 25 | The minimum nadir day |
| 26 | The minimum peak day |
| 27 | The minimum time to peak |
| 28 | The maximum duration of menstrual phase |
| 29 | The maximum length of cycle recorded period |
| 30 | The maximum cycle length |
| 31 | The maximum proportion of recorded cycle period |
| 32 | The maximum length of the line of the cycle curve |
| 33 | The maximum start day of three-day period with the largest rise in temperature |
| 34 | The maximum temperature rise of three-day period with the largest rise |
| 35 | The maximum start day of four-day period with the largest rise in temperature |
| 36 | The maximum temperature rise of four-day period with the largest rise |
| 37 | The maximum day of temperature change |
| 38 | The maximum difference in temperature before and after detected temperature change |
| 39 | The maximum DTW distance |
| 40 | The maximum length of optimal warping path |
| 41 | The maximum nadir temperature |
| 42 | The maximum peak temperature |
| 43 | The maximum difference between nadir and peak temperatures |

| | **The Features** |
|---|---|
| 44 | The maximum nadir day |
| 45 | The maximum peak day |
| 46 | The maximum time to peak |
| 47 | The median duration of menstrual phase |
| 48 | The median length of cycle recorded period |
| 49 | The median cycle length |
| 50 | The median proportion of recorded cycle period |
| 51 | The median length of the line of the cycle curve |
| 52 | The median start day of three-day period with the largest rise in temperature |
| 53 | The median temperature rise of three-day period with the largest rise |
| 54 | The median start day of four-day period with the largest rise in temperature |
| 55 | The median temperature rise of four-day period with the largest rise |
| 56 | The median day of temperature change |
| 57 | The median difference in temperature before and after detected temperature change |
| 58 | The median DTW distance |
| 59 | The median length of optimal warping path |
| 60 | The median nadir temperature |
| 61 | The median peak temperature |
| 62 | The median difference between nadir and peak temperatures |
| 63 | The median nadir day |
| 64 | The median peak day |
| 65 | The median time to peak |
| 66 | The range of duration of menstrual phase |
| 67 | The range of the length of cycle recorded period |
| 68 | The range of the cycle length |
| 69 | The range proportion of recorded cycle period |
| 70 | The range of the length of the line of the cycle curve |
| 71 | The range of the start day of three-day period with the largest rise in temperature |
| 72 | The range of temperature rise of three-day period with the largest rise |
| 73 | The range of start day of four-day period with the largest rise in temperature |
| 74 | The range of temperature rise of four-day period with the largest rise |
| 75 | The range of days of temperature change |
| 76 | The range of difference in temperature before and after detected temperature change |
| 77 | The range of DTW distance |
| 78 | The range of length of optimal warping path |
| 79 | The range of nadir temperature |
| 80 | The range of peak temperature |
| 81 | The range of the difference between nadir and peak temperatures |
| 82 | The range of nadir day |
| 83 | The range of peak day |
| 84 | The range of time to peak |

**Table S3: The hyperparameters for logistic regression.** This table details the search space for the logistic regression classifier. The solvers were selected based on their compatibility with specific regularisation penalties (L1, L2, and Elastic Net) to ensure convergence.

| | **Hyperparameter** | **Range/Values Tested** | **Explanation** |
|---|---|---|---|
| 1 | penalty | *L1*, *L2*, *elasticnet* | Regularization (Lasso for sparsity, Ridge for shrinkage, and a hybrid) |
| 2 | *C* (Inverse Regularization) | 100, 10, 1.0, 0.1, 0.01 | Optimizing the bias-variance trade-off |
| 3 | *L1_ratio* | 0.0, 0.25, 0.5, 0.75, 1.0 | Spectrum of the L1 mixing parameter for the elasticnet penalty |
| 4 | solver | *newton-cg*, *lbfgs*, *liblinear, sag*, *saga* | Solvers selected for compatibility with the respective penalties and scalability |

**Table S4: The hyperparameters for support vector machines.** The table lists the parameters tuned for the SVM classifier. Both linear and non-linear (RBF) kernels were evaluated, with the regularisation parameter (C) and the kernel coefficient (gamma) varied across a logarithmic scale to optimise the balance between margin maximisation and misclassification error.

| | **Hyperparameter** | **Range/Values Tested** | **Explanation** |
|---|---|---|---|
| 1 | *C* (Regularization) | 0.1, 1, 10 | Controlling the penalty for misclassification errors. |
| 2 | *gamma* (Kernel Coefficient) | scale, auto | Scaling using default and auto to define the influence of a single training example. |
| 3 | kernel | linear, rbf | Linear and non-linear boundaries |

**Table S5: The hyperparameters for the random forest classifier.** This table details the structural parameters for the RFC ensemble. The parameter tuning focused on the number of estimators and tree depth to prevent overfitting while ensuring model stability. Note that *max_features* was maintained at the default.

| **Hyperparameter** | **Range/Values Tested** | **Explanations** |
|---|---|---|
| *n_estimators* (Tree Count) | 10, 50, 100, 200 | Increasing numbers of trees to ensure stable and robust ensemble predictions. |
| *max_depth* (Tree Depth) | None, 5, 10 | Fixed depths to prevent overfitting, including "None" (full depth) as a baseline. |
| *min_samples_split* | 2, 5, 10 | The minimum number of samples required to split an internal node. |

**Table S5: Comparative performance of PCOS classification models across the various feature sets.** Values represent the mean AUC-ROC score, with the corresponding standard deviation shown in parentheses, obtained from five-fold cross-validation. Feature sets are grouped by main and comparison categories to evaluate the predictive performance across cycle-level, user-level, questionnaire-level, and combined data streams.

| | **Main Feature Sets** | | **Comparison Feature Sets** | |
|---|---|---|---|---|
| | Cycle Level | User Level | Questionnaire Level | Combined User and Questionnaire Level |
| Logistic Regression | 0.64 (0.02) | 0.65 (0.07) | 0.69 (0.06) | 0.71 (0.06) |
| Support Vector Machine | 0.67 (0.03) | 0.68 (0.06) | 0.69 (0.05) | 0.71 (0.05) |
| Random Forest Classifier | 0.68 (0.04) | 0.70 (0.04) | 0.69 (0.03) | 0.72 (0.03) |

**Table S6: Calibration results for the cycle-level models**. Values represent mean Brier loss, calibration slope, and intercept with 95% confidence intervals. Brier loss measures the accuracy of probabilistic predictions

| Model | Mean Brier Loss | Slope | Intercept |
|---|---|---|---|
| Logistic Regression | 0.241 ± 0.008 | 1.05 (0.69, 1.40) | -0.02 (0.14, 0.10) |
| Support Vector Machine | 0.229 ± 0.006 | 0.87 (0.70, 1.05) | 0.01 (-0.11, 0.13) |
| Random Forest Classifier | 0.226 ± 0.012 | 0.83 (0.68, 1.00) | 0.02 (-0.11, 0.14) |

**Table S7: Calibration results for the user-level models.** Values represent mean Brier loss, calibration slope, and intercept with 95% confidence intervals. Brier loss measures the accuracy of probabilistic predictions

| **Model** | **Mean Brier Loss** | **Slope** | **Intercept** |
|---|---|---|---|
| Logistic Regression | 0.242 ± 0.024 | 0.47 (0.22, 0.79) | -0.01 (-0.22, 0.15) |
| Support Vector Machine | 0.228 ± 0.023 | 0.81 0.64, 1.09) | -0.04 (-0.25, 0.12) |
| Random Forest Classifier | 0.219 ± 0.012 | 0.88 (0.69, 1.15) | -0.01 (-0.22, 0.16) |

**Table S8: Calibration results for the questionnaire-level models.** Values represent mean Brier loss, calibration slope, and intercept with 95% confidence intervals. Brier loss measures the accuracy of probabilistic predictions

| **Model** | **Mean Brier Loss** | **Slope** | **Intercept** |
|---|---|---|---|
| Logistic Regression | 0.228 ± 0.015 | 1.24 (0.84, 1.70) | 0.01 (-0.20, 0.17) |
| Support Vector Machine | 0.221 ± 0.012 | 1.01 (0.74, 1.32) | -0.01 (-0.21, 0.16) |
| Random Forest Classifier | 0.223 ± 0.009 | 0.95 (0.71, 1.22) | 0.00 (-0.19, 0.6) |

**Table S9: Calibration results for combined user-level and questionnaire feature models.** Values represent mean Brier loss, calibration slope, and intercept with 95% confidence intervals. Brier loss measures the accuracy of probabilistic predictions.

| **Model** | **Mean Brier Loss** | **Slope** | **Intercept** |
|---|---|---|---|
| Logistic Regression | 0.224 ± 0.018 | 1.01 (0.65, 1.44) | 0.01 (-0.18, 0.17) |
| Support Vector Machine | 0.221 ± 0.019 | 0.98 (0.75, 1.30) | 0.00 ( -0.23, 0.16) |
| Random Forest Classifier | 0.212 ± 0.010 | 1.02 (0.85, 1.34) | -0.01(-0.22, 0.18) |
