## Supplementary material for "A digital health approach for identifying polyendocrine metabolic ovarian syndrome using machine learning and body temperature": Multimedia appendix 4

### Fertility questionnaire

**Please give your name and the email address:**
…………………………………………

**Please give your date of birth:**
dd/mm/yyyy
(US version mm/dd/yyyy)

**Please tick all the boxes to indicate agreement with the statements below:**

☐ I confirm that I have read and understand the information sheet for the above study. I have had the opportunity to consider the information, ask questions and have had any questions answered satisfactorily.

☐ I understand that my participation is voluntary and that I am free to withdraw at any time without giving any reason, and without consequence or liability.

☐ I understand that my anonymised data, collected during the study, will be used by researchers from the University of Bristol, UK and Carolinas Medical Center, Charlotte, USA.

**Please choose from one answer below to indicate your consent to take part in the survey and study, or to leave the survey:**

○ I agree to take part in the survey and study (*If chosen go to A1)*

○ I don't wish to continue with the survey or study (*If chosen show disqualification message)*

© The University of Bristol, 2018

### Section A: About you

**A1. How would you describe the race or ethnic group of yourself and your natural parents? (tick all that apply)**

| | **Yourself** | **Your mother** | **Your father** |
|---|---|---|---|
| White | | | |
| Black/Caribbean | | | |
| Black/African | | | |
| Black/other | | | |
| Indian | | | |
| Pakistani | | | |
| Bangladeshi | | | |
| Chinese | | | |
| Hispanic | | | |
| Any other ethnic group | ……………….. | ……………….. | ……………….. |

**A2.** **a) Are you currently employed or self-employed?**

Yes
No *If no go to A4d*
Prefer not to answer *If prefer not to answer go to A4d*

*If yes:*

**b) What is your occupation (or job title)?**
………………………

**c) Does your work involve night shifts?** (*If you have more than one 'current job' then answer this question for your MAIN job only. Night shifts are a work schedule that involves working through the normal sleeping hours, for instance working through the hours from 12am [midnight] to 6am)*

Never/rarely
Sometimes
Usually
Always
Do not know
Prefer not to answer

**d) Do you have a university degree?**

Yes
No

**A3.** **a) Do you have a partner?**

Yes
No *if no go to A6*
Prefer not to answer *If prefer not to say go to A6*

© The University of Bristol, 2018

*If yes*

**b) Is your partner currently employed or self-employed?**
Yes
No *If no go to A5d*
Prefer not to say *If prefer not to say go to A5d*

*If yes:*

**c) What is their occupation (or job title)?**
………………………

**d). Does your partner have a university degree?**
Yes
No

**A4. a) What units do you use to measure your weight**

- ◯ Stones
- ◯ Kilos
- ◯ Pounds (US)
- ◯ Prefer not to give my weight *(if ticked go to A7)*

**b) What is your current weight?**
…………………………….

**A5. a) What units do you use to measure your hip size?**

- ◯ cm
- ◯ inches
- ◯ Prefer not to give my hip size *(if ticked go to A7ii)*

© The University of Bristol, 2018

**What size are your hips?**

…………….

**b) What units do you use to measure your waist size?**

- ◯ cm
- ◯ inches
- ◯ Prefer not to give my waist size *(if ticked go to A7iii)*

**What size is your waist?**

…………….

**c) What units do you use to measure your bust circumference?**

- ◯ cm
- ◯ inches
- ◯ Prefer not to give my bust size *(if ticked go to A8)*

**What size is your bust circumference?**

…………….

© The University of Bristol, 2018

**A6. What units do you use to measure your height?**

- ◯ Feet and inches
- ◯ Centimeters

◯ Prefer not to give my height *(if ticked go to B1)*

**How tall are you?**

…………….

#### Section B: Lifestyle

**B1. a) Have you ever been a regular smoker?**
Yes
No *if no, go to B2*
Prefer not to answer *if prefer not to answer, go to B2*

*if yes*

**b) At what age did you start smoking regularly?**
……….. years

**c) Which of the following have you ever smoked regularly? (tick all that apply)**
Cigarettes
Pipe
Cigar
e-cigarette
other

**d) Have you now stopped smoking?**
Yes
No *If no go to B1g*
Prefer not to answer *If prefer not to answer go to B2*

*If yes*

**e) Please enter the estimated date on which you stopped smoking. If you do not know the month, please just give the year**
…… Month (as number mm) ……. Year (as number yyyy)

**f) If you smoked cigarettes, how many did you smoke per day?**
On weekdays: ……
On weekend days ……

*If no*

**g) If you smoke cigarettes, how many do you currently smoke per day?**

© The University of Bristol, 2018

On weekdays: ……
On weekend days ……

**B2. Which of the following statements about alcohol best applies to yourself** (1 glass = 1 standard bar measure of spirits or 1 small glass of wine or a 1/2 pint of beer or cider)**:**

Never drink alcohol
Very occasionally (less than once a week)
Occasionally (at least once a week)
Drinks 1-2 glasses* nearly every day
Drinks 3-9 glasses* every day
Drinks at least 10 glasses a day
Don't know

**B3. a) Compared with other women of your age, would you consider yourself to be:**

Much more active
Somewhat more active
About the same
Somewhat less active
Much less active

**b) Currently at least once a week do you engage in any regular activity like brisk walking, gardening, housework, jogging, cycling, etc. long enough to work up a sweat?**

Yes
No *if no go to section B4*
Prefer not to answer *if prefer not to answer go to section B4*

*If yes*

**c) How many hours a week:**
…………. hours

**d) How much do you do the following at present?**

| | **More than 7 hours per week** | **2-6 hours per week** | **Less than one hour per week** | **Never** |
|---|---|---|---|---|
| Jogging | | | | |
| Aerobics | | | | |
| Keep fit exercises | | | | |
| Yoga | | | | |
| Squash | | | | |
| Tennis/badminton | | | | |
| Swimming | | | | |
| Brisk walking | | | | |
| Weight training | | | | |
| Cycling | | | | |
| Other exercise | | | | |

……………………….…………………………..……………………….

© The University of Bristol, 2018

© The University of Bristol, 2018

………………………..……………………...………………………..

B4. a) About how many hours sleep do you get in every 24 hours? (please include naps)
*sliding scale from 0 to 14*

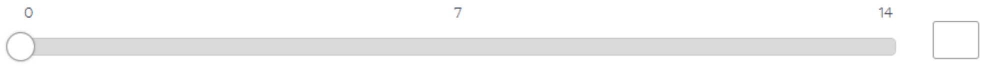


b) Do you have trouble falling asleep at night or do you wake up in the middle of the night?
Never/rarely
Sometimes
Usually
Prefer not to answer

c) How likely are you to doze off or fall asleep during the daytime when you don't mean to?
Never/rarely
Sometimes
Often
Do not know
Prefer not to answer
All of the time

d) Do you consider yourself to be?
Definitely a 'morning' person
More a 'morning' than 'evening' person
More an 'evening' than a 'morning' person
Definitely an 'evening' person
Do not know
Prefer not to answer

#### Section C: General health

**C1. a) Have you ever had diabetes?**
Yes
No *if no, go to C2*
Prefer not to answer *if prefer not to answer, go to C2*

*If yes*

**b) Have you only had it when you were pregnant?**
Not applicable
Yes
No

© The University of Bristol, 2018

**c) How old were you (in years) when you first developed it?**
…………….

**d) How was/ is your diabetes treated? (please tick all that apply)**
Diet and exercise advice
Drugs that you took/ take by mouth
Insulin injections

**C2.** **a) Have you ever had hypertension (high blood pressure)?**
Yes
No *if no, go to C3*
Prefer not to answer *if prefer not to answer, go to C3*

*if yes*

**b) Have you had hypertension only when pregnant?**
Not applicable
Yes
No

**c) How old were you (in years) when you first developed it?**
………..

**d) Are you currently taking drugs prescribed by a doctor for high blood pressure?**
Yes
No
Prefer not to answer *if prefer not to answer, go to C3*

*if Yes*

**e) Please write the name of the drug you are taking for high blood pressure (e.g. Ramipril)**

………………………….

**f) What is the dose of the drug? (e.g. 5mg)**

………………………….

**g) Please enter the frequency (e.g. twice per day)**

………………………….

**C3.** **a) Are there any problems for which you have regular treatment or medicine?**

© The University of Bristol, 2018

Yes
No *If no, go to section D*
Prefer not to answer *If prefer not to answer, go to section D*

*If yes*

**b) Please describe the problem and <u>regular</u> treatment or medicine (include the name of any drug/medicine and the dose and frequency that you take):**

Problem 1: ……………………………………………………………………
Treatment 1: ……………………………………………………………………
Problem 2: ……………………………………………………………………
Treatment 2: ……………………………………………………………………
Problem 3: ……………………………………………………………………
Treatment 3: ……………………………………………………………………
Problem 4: ……………………………………………………………………
Treatment 4: ……………………………………………………………………
Problem 5: ……………………………………………………………………
Treatment 5: ……………………………………………………………………
Problem 6: ……………………………………………………………………
Treatment 6: ……………………………………………………………………
Please add any other problems and treatments not covered above:
……………………………………………………………………………………

#### Section D: Your feelings in the past week

**Your feelings in the past week.**

**D1. I have been able to laugh and see the funny side of things:**
As much as I always could
Not quite so much now
Definitely not so much now
Not at all

**D2. I have looked forward with enjoyment to things:**
As much as I ever did
Rather less than I used to
Definitely less than I used to
Hardly at all

**D3. I have blamed myself unnecessarily when things went wrong:**
Yes, most of the time
Yes, some of the time
Not very often

© The University of Bristol, 2018

Never

**D4. I have been anxious or worried for no good reason:**
No, not at all
Hardly ever
Yes, sometimes
Yes, often

**D5. I have felt scared or panicky for no very good reason:**
Yes, quite a lot
Yes, sometimes
No, not much
No, not at all

**D6. Things have been getting on top of me:**
Yes, most of the time I haven't been able to cope
Yes, sometimes I haven't been coping as well as usual
No, most of the time I have coped quite well
No, I have been coping as well as ever

**D7. I have been so unhappy that I have had difficulty sleeping:**
Yes, most of the time
Yes, sometimes
Not very often
No, not at all.

**D8. I have felt sad or miserable:**
Yes, most of the time
Yes, quite often
Not very often
No, not at all

**D9. I have been so unhappy that I have been crying:**
Yes, most of the time
Yes, quite often
Only occasionally
Never

**D10. The thought of harming myself has occurred to me:**
Yes, quite often
Sometimes
Hardly ever
Never

**D11. I have felt stressed:**
Yes, most of the time
Yes, quite often
Not very often
No, not at all

© The University of Bristol, 2018

### Section E: Previous (or current) pregnancies

**E1. a) Are you currently pregnant?**

Yes
No *if no, go to E2*
Prefer not to answer *if prefer not to answer, go to E2*

**b) How many weeks pregnant are you?**

……………………

**c) How long were you trying before you got pregnant?**

Less than 6 months
6-11 months
12 months or more
Pregnancy wasn't planned
Don't remember

**d) Have you had excessive vomiting which meant you have had to stay in hospital for some days during this pregnancy?**

Yes*
No

*** If yes, please say how many days you were you in hospital for**

………………

**e) Have you been told by a doctor or midwife that you have had gestational diabetes during this pregnancy?**

Yes
No *if no, go to E1f*
Prefer not to answer *if prefer not to answer, go to E1f*

**(If yes) How is your gestational diabetes treated? (please tick all that apply)**

Diet and exercise advice
Drugs that you took by mouth
Insulin

**f) Have you been told by a doctor or midwife that you have had pre-eclampsia during this pregnancy?**

Yes
No *if no, go to E1g*
Prefer not to answer *if prefer not to answer, go to E1g*

**(If yes) Have you been admitted to hospital for pre-eclampsia during this pregnancy?**

Yes

© The University of Bristol, 2018

No

**g) Have you been told by a doctor or midwife that you have had gestational hypertension (also called pregnancy related or pregnancy induced hypertension) during this pregnancy?**

Yes
No
Prefer not to answer

**E2. a) Have you been pregnant in the past?**

Yes
No *if no, go to Section F*
Prefer not to answer *if prefer not to answer, go to F*

**b) How many times have you been pregnant in the past?**

- ○ 1
- ○ 2
- ○ 3
- ○ 4
- ○ 5
- ○ 6
- ○ 7
- ○ 8
- ○ 9
- ○ 10

*For each pregnancy please give the following details (if you have had more than 10 pregnancies then please give details for the 10 most recent):*

**c) How long were you trying before you got pregnant?**

Less than 6 months
6-11 months
At least 12 months
Pregnancy wasn't planned
Don't remember

**d) Did this result in a live birth?**

Yes *if yes go to E2e*
No
Prefer not to answer

*If no*
Please give the date of the end of this pregnancy.
*Go to next pregnancy*

© The University of Bristol, 2018

**e) Please give the date of birth.**

**f) Please give the units of weight for your baby**
pounds ounces ◉ kilograms ○ ○

**g) Please give the birth weight of your baby?**
……………………

**h) Did you have excessive vomiting which meant you had to stay in hospital for some days during this pregnancy?**
Yes
No

*** If yes, please say how many days you were you in hospital for**
………………

**i) Were you <u>told by a doctor or midwife that you had gestational diabetes during this pregnancy?</u>**
Yes
No *if prefer not to answer, go to E2j*
Prefer not to answer *if prefer not to answer, go to E2j*

*If yes*, how was your gestational diabetes treated? (please tick all that apply)
Diet and exercise advice
Drugs that you took by mouth
Insulin

**j) Were you <u>told by a doctor or midwife that you had pre-eclampsia during this pregnancy?</u>**
Yes
No *if prefer not to answer, go to E2k*
Prefer not to answer *if prefer not to answer, go to E2k*

*If yes*,
Did you need to be admitted to hospital for the pre-eclampsia during this pregnancy?
Yes
No

Did your baby need to be delivered early during this pregnancy because of the pre-eclampsia?
Yes
No

**k) Were you <u>told by a doctor or midwife that you had gestational hypertension (also called pregnancy related or pregnancy induced hypertension) during this pregnancy?</u>**
Yes
No
Prefer not to answer

© The University of Bristol, 2018

### Section F: Current fertility

**F1.** **a) How old were you when your periods (menstrual bleeds) first started?**

I have not had periods *go to F3*
Do not remember
Prefer not to answer
My age when my periods first started was:
……...

**b) Have you had a period in the last 3 months?**
Yes *if yes, go to F2*
No
Prefer not to answer *if prefer not to answer, go to F2*

*If no*

**c) What is the reason you haven't had a period in the past 3 months?**
Irregular periods
Contraceptive injection
Contraceptive implant
Intrauterine contraceptive device (IUD, also known as coil)
Contraceptive tablets
Don't know
Other ………………

**F2.** **a) What was the date of the first day of your last period? (if you cannot remember the exact day please provide the month and year)**
Day (as number dd) ……
Month (as number mm) ..….
Year (as number yyyy) ……

**b) Are your periods regular?**
Yes
No
Prefer not to answer *if prefer not to answer go to F2.c*

*If yes*

**How regular are your periods?**
Every 24 to 27 days
Every 28 to 30 days
Every 30 or more days
Prefer not to answer

© The University of Bristol, 2018

*If no*

**How many periods do you have each year?**
1-2
3-6
7-12
More than 12
Prefer not to answer

**c) How would you describe your most recent (last six) periods:**

| | Very | Moderately | Mildly | Not at all |
|---|---|---|---|---|
| How heavy were your periods? | | | | |
| How painful were your periods? | | | | |

**F3.** **a) Have you ever used the contraceptive pill?**
Yes
No *If no go to F4*
Prefer not to answer *If prefer not to answer, go to F4*

*If yes,*

**b) How old were you (in years) when you first took the contraceptive pill?**
……..

**c) How many years altogether did you take a contraceptive pill?**
For less than 1 year
1-2 years
3-4 years
5 years or more

**d) Are you currently using the contraceptive pill?**
Yes
No

**F4.** **a) Have you ever gone to a doctor because you thought you were infertile?**
Yes
No *If no go to F5*
Prefer not to answer *If prefer not to answer, go to F5*

*If yes,*

**b) What diagnoses were made? (tick all that apply)**
Polycystic ovary syndrome (PCOS)
Other ovarian problem
Endometriosis
Problem with your cervix

© The University of Bristol, 2018

Partner has a low sperm count or other problems with sperm
Overactive thyroid
Underactive thyroid
Pelvic inflammatory disease
Diagnosis not made
Other (please specify)…………….

**c) Which treatments were given? (tick all that apply)**
Hormonal ovarian stimulation (please specify in the box below)
Other fertility drugs (please specify in the box below)
Intrauterine insemination (IUI)
IVF
Surgery
Other (please describe in detail)

**d) Please specify the drug treatments you were given in as much detail as possible (drug names, dosages, when you took them)?**
……………………………………………………………………………………………
……………………………………………………………………………………………
……………………………………………………………………………………………

**e) If you had IVF (with or without ICSI), was it performed with:**
Own eggs and partner's sperm
Own eggs and donor sperm
Donor eggs and partner's sperm
Donor eggs and donor sperm

**F5. a) Have you ever had a D and C (scrape)?**
Yes
No *if no go to F6*
Don't know *if don't know go to F6*
Prefer not to answer *if prefer not to answer go to F6*

*If yes*

**b) Why did you have a D and C scrape? (tick all that apply)**

| | Yes | No |
|---|---|---|
| i) Heavy periods | | |
| ii) Painful periods | | |
| iii) Fibroids | | |
| iv) Termination | | |
| v) Infertility | | |
| vi) Miscarriage | | |
| vii) Don't know | | |
| viii) Other (please tick and describe) ………………………………… | | |

**F6. Have you ever had any of the following sexually transmitted infections? (tick all that apply)**
Chlamydia

© The University of Bristol, 2018

Gonorrhea
Genital herpes
Syphilis
HIV/AIDS
Human papillomavirus (HPV)
Trichomoniasis
Other (please specify) ………………..

**F7.** **a) What is your main reason for using OvuSense?**
I have been trying to get pregnant and have not been able to
I have reduced fertility and it might help
I am trying to avoid becoming pregnant
I am just interested in monitoring my cycles
Other ……………..

*If you are trying to get pregnant, then*

**b) Are you using any treatments to help you conceive at the moment?**
Yes
No
Prefer not to answer

*If yes*

**c) Which treatments have you been given? (tick all that apply)**
Hormonal ovarian stimulation (please specify in the box below)
Other fertility drugs (please specify in the box below)
Intrauterine insemination (IUI)
IVF (with or without ICSI)
Surgery
Supplements of homeopathic remedies (e.g. Inositol)
None
Prefer not to answer
Other (please describe in detail)

**d) Please specify the drug treatments you have been given in as much detail as possible (drug names, dosages, when you took them)?**
……………………………………………………………………………………………
……………………………………………………………………………………………
……………………………………………………………………………………………

**e) If you had IVF (with or without ICSI), was it performed with:**
Own eggs and partner's sperm
Own eggs and donor sperm
Donor eggs and partner's sperm
Donor eggs and donor sperm

© The University of Bristol, 2018

**VERY MANY THANKS FOR ALL YOUR HELP**

© The University of Bristol, 2018
