## Supplementary material for "A digital health approach for identifying polyendocrine metabolic ovarian syndrome using machine learning and body temperature": Multimedia appendix 5

Bristol Medical School
Dr Louise A C Millard


University of BRISTOL

**Study Information Sheet**

### Evaluating continuous overnight monitoring of menstrual cycles: a study of women using the OvuSense device.

Thank you for expressing interest in this study. This information sheet will provide an outline of the aims of the research and describe what you will be asked to do as a participant in the study, and what will happen to the data that we collect from you. If you have any questions on any aspect of the study, please contact the researcher (contact details above).

#### What is the study about?

You have been asked to participate in this study as a user of the OvuSense device.

We are particularly interested in understanding how menstrual cycles differ between women, and the reasons for these differences. The OvuSense device captures temperature inside the vagina every five minutes during the night, and this is currently used to predict day of ovulation up to one day in advance. This works by detecting a change in temperature that commonly occurs during ovulation. It may be the case that different changes in temperature during a menstrual cycle relate to other aspects of a woman's health.

In this study, we will use temperature change data to identify different menstrual cycle patterns. We will then examine whether we can use the temperature data to predict fertility problems, such as polycystic ovary syndrome. We will also explore whether ovulation can be predicted earlier during each cycle (e.g. 5 days before the date of ovulation), and the extent to which a device such as OvuSense can be used for pregnancy prevention, by identifying 'safe' days when a woman would be unlikely to conceive.

#### Who is doing this study?

This study will be done by researchers at the MRC Integrative Epidemiology Unit at the University of Bristol, a leading centre for research into women's reproductive health and for analysing complicated data with lots of repeat measurements (such as the repeat OvuSense temperature data). Given the large number of US OvuSense users, assistance with the study will also be provided by Carolinas Medical Center, Charlotte, USA. Dr Louise Millard at the University of Bristol is the lead researcher for this study.

#### What will we ask you to do?

We will ask you to complete a questionnaire and we will ask you for consent to use this data together with your temperature data recorded by the OvuSense device. This questionnaire should take 20-60 minutes to complete, depending upon the extent of your TTC (trying to conceive) experiences. The questionnaire will ask about your age, lifestyle (e.g. whether you smoke, how much exercise you do), whether you have any health problems, and about whether you have been pregnant before, had problems trying to get pregnant and your use of contraception. We will also ask your reasons for using OvuSense, and for you to tell us how much you weigh and how tall you are. With your consent we will link your OvuSense temperature data to these questionnaire data so that we can do our research.

#### How will your data be used and stored?

Researchers at the MRC Integrative Epidemiology Unit at the University of Bristol run and use data from a large number of studies. We are very experienced at ensuring that your data are protected and comply with all data regulatory requirements. Also, we will not actually hold your name or other personal identifiers as all data supplied to us by Fertility Focus (the Company who owns OvuSense) will be anonymised.

We will use data from both the OvuSense device and the questionnaire to determine what factors, such as a woman's age, lifestyle, health issues or fertility, relate to different patterns of temperature change and menstrual cycles. As above, both the device and questionnaire data will be anonymised by removing personal information (eg your name and address) before any data are sent to researchers at the University of Bristol. Data will be stored securely, and the anonymised data will only be used by the researchers working on this project.

#### What are the benefits of participating in this study?

This is a research study. We currently do not know if the OvuSense temperature data can be used to identify women with problems, such as polycystic ovarian disease, or to accurately identify safe periods when conception will not happen. Therefore, we will not be able to provide you with any information that could directly benefit you. Depending on the findings of our research we might be able to improve the OvuSense device, for example, by improving the ability to predict timing of ovulation. Our results might also improve understanding of women's fertility. You would be contributing to this new knowledge and might indirectly benefit from it in the future.

#### What if I do not want to take part, or want to stop participating?

It is your choice whether you take part or not. There are three levels of participation you can choose:

a) Full participation in this study: You decide to complete the questionnaire, and we will use these answers in our study, linked to the temperature data from your OvuSense device.
b) Partial participation in this study: As part of the terms of conditions of using the OvuSense device you provided consent that your temperature data can be used for research purposes (https://www.ovusense.com/uk/terms-and-conditions/; point 5 in 'General'

section). If you are happy for us to use your temperature data but do not want to complete the questionnaire, then you do not need to do anything further.

c) No participation: If you no longer want your data to be used in this study at all, then please to let us know, and we will remove you from the study.

If you decide to take part and then want to stop being in the study you can let us know and, depending on your preference, we will stop any further data collection and/or also remove all your existing data that has been collected.

#### Who has reviewed this study?

This study has been reviewed by the ethics committee of the Faculty of Health Sciences, University of Bristol.

#### Who can I contact about this study?

For further information about the study, please contact Dr Louise Millard using the contact details at the top of this sheet.

Should you wish to make a formal complaint about any aspect of this study or its implementation, please write to:
